# AlphaGenome Atlas: *in silico* mutagenesis of the entire human genome improves prioritization and interpretation of non-coding variants

**DOI:** 10.64898/2026.09.16.26363192

**Authors:** Lead authors:, Jun Cheng, Kyle R. Taylor, Lauren Nicolaisen, Joshua Pan, Clare Bycroft, Matteo Perino, Tom Ward, Gareth Hawkes, Laura E. Covill, Melanie Weilert, Raina W. Thomas, Natasha Latysheva, Contributing authors:, Maile J. Hirschmann, Xi Dawn Chen, Robin N Beaumont, V Kartik Chundru, Michael N Weedon, Simon Bourdareau, Hoyin Chu, Dhavanthi Hariharan, Thais Kagohara, Lucas Tenório, Yosuke Ushigome, Courtney A. Shearer, Barbara Ikica, Ada Fang, Mouad Naciri, Victoria Johnston, Richard Green, Lai Hong Wong, Vincent Dutordoir, Anne Mottram, Adam Gayoso, Eirini Arvaniti, Guido Novati, Supervising authors:, Heidi L. Rehm, Fei Chen, Caleb A. Lareau, Caroline F Wright, Anne O’Donnell-Luria, Julia Zeitlinger, Pushmeet Kohli, Žiga Avsec

**Author notes:** These authors contributed equally. Correspondence to (J.C.); (P.K.); (Ž.A.).

## Abstract

A major challenge in genomics is deciphering the functional consequences of non-coding genetic variation. Here we present AlphaGenome Atlas, a comprehensive resource that enables the joint interpretation and prioritization of variant effects across the entire human genome. Using AlphaGenome, we predicted the regulatory effects across thousands of molecular phenotypes for every possible human single nucleotide variant and many observed indels. These predictions were then used to derive a unified and interpretable AlphaGenome Variant Impact (AVI) score and to map cis-regulatory motifs across the genome. AVI achieved state-of-the-art performance across diverse benchmarks with improved prioritization of deleterious non-coding variants. Application of the combined Atlas resource helped solve an epileptic encephalopathy rare disease case, increased the statistical power to detect rare non-coding variants driving population-level phenotypes, and enhanced the mechanistic interpretation of these variants. Thus, AlphaGenome Atlas improves the prioritization and molecular interpretation of non-coding variants with genetic and clinical significance.

## Introduction

From trait associations across populations to rare disease diagnosis in individuals, human genetics is bottlenecked by our ability to understand the function of the variants that we discover (*1*). Progress hinges on overcoming two challenges: prioritizing genetic variants by their likelihood of functional impact and accurately interpreting their molecular effects. Non-coding variants, which approximately account for 98% of human genetic variation and influence when and where genes are expressed, are especially challenging because of the complexity of the cis-regulatory code. Plateauing rates of variant identification in population biobanks and labor-intensive experimental characterizations are unlikely to close this gap in the near term. Since demographic models suggest that every fitness-compatible variant currently exists in the global population (*2*), genome-scale variant prioritization and interpretation is a pressing need.

While computational approaches operate at genome scale, current methods trade off between the precision required for ranking variant impacts and the breadth of cis-regulatory information necessary to interpret their molecular effects. Genomic annotation frameworks like Ensembl Variant Effect Predictor (VEP) (*3*) and ENCODE candidate *cis*-Regulatory Elements (cCREs) (*4*) define broad regulatory regions but lack the basepair resolution to map the 4–20 basepair (bp) motifs of DNA and RNA binding proteins that regulate gene expression, and thus cannot easily prioritize individual variants. Conversely, evolutionary models such as GPN-Star (*5*) prioritize variants by leveraging cross-species conservation, but lack molecular interpretability and can overlook non-conserved yet functionally critical variants. Although supervised frameworks like Combined Annotation-Dependent Depletion (CADD) (*6*, *7*) aggregate disparate annotations into unified impact scores by using allele frequencies as a proxy for negative selection, their efficacy for non-coding variants depends on the quality of the input features.

Sequence-to-function models take DNA sequences as input and predict the readout of various genomics assays profiling specific steps of gene regulation, thereby learning how these processes are encoded by features like motif combinations in individual DNA regions (*8*). By comparing predictions between reference and alternative sequences, such models can score the effects of variants unseen during model training. We recently described AlphaGenome (*9*), a state-of-the-art sequence-to-function model unifying long 1 megabase (Mb) DNA context, basepair resolution outputs, and joint prediction of multiple genomic assay modalities including TF binding, chromatin accessibility, histone modifications, gene transcript abundance, and splicing. However, because AlphaGenome requires on-the-fly inference for each variant, large-scale variant predictions and an interpretation of AlphaGenome’s learned cis-regulatory motif maps have yet to be performed. We reasoned that precomputing an allelic-resolution database of regulatory variant effects genome-wide would allow us to identify and annotate the cis-regulatory motifs learned by AlphaGenome, while also augmenting non-coding performance of a CADD-style variant prioritization model by leveraging AlphaGenome predictions as input features.

Here, we present AlphaGenome Atlas, a comprehensive resource for the joint prioritization and interpretation of genetic variants. At its core, we performed *in silico* saturation mutagenesis (ISM) across the human genome, generating precomputed predictions for all possible human single nucleotide variants (SNVs) as well as indels observed in different population datasets. To enable variant prioritization, we trained the AlphaGenome Variant Impact (AVI) score, integrating these ISM predictions with AlphaMissense, conservation metrics, and protein-coding features in a supervised framework. We further engineered Atlas for interpretability: we computed SHAP (SHapley Additive exPlanations) feature attributions (*10*) for every AVI score to decompose variant impact into specific coding and non-coding molecular effects, and derived *de novo* motifs directly from the AlphaGenome ISM to map high-impact non-coding variants to their underlying regulatory motifs.

To our knowledge, this represents the most comprehensive repository of precomputed variant effect predictions to date. To navigate this resource, we provide a technical overview (Fig. 1) and comprehensively evaluate AVI against variant impact benchmarks (Fig. 2). We then demonstrate the specific utility of the Atlas to analyse rare non-coding variants. First, we deployed AVI to help resolve a rare disease case within the Genomics Research to Elucidate the Genetics of Rare diseases (GREGoR) cohort (Fig. 3). Second, we show how Atlas features can increase statistical power for identifying rare non-coding variants driving circulating protein levels across the population (Fig. 4). Finally, we demonstrate how AVI and AlphaGenome are interpretable by connecting variant effects to molecular effects through feature attribution (Fig. 5) and genome-wide motif mapping (Fig. 6). To enable broad access, we made precomputed scores, AVI and AVI feature attributions, curated motifs and genome-wide instances available through a programmatic API alongside a fully interactive graphical user interface for rapid exploration (https://alphagenome.google/atlas).

**Fig. 1.**
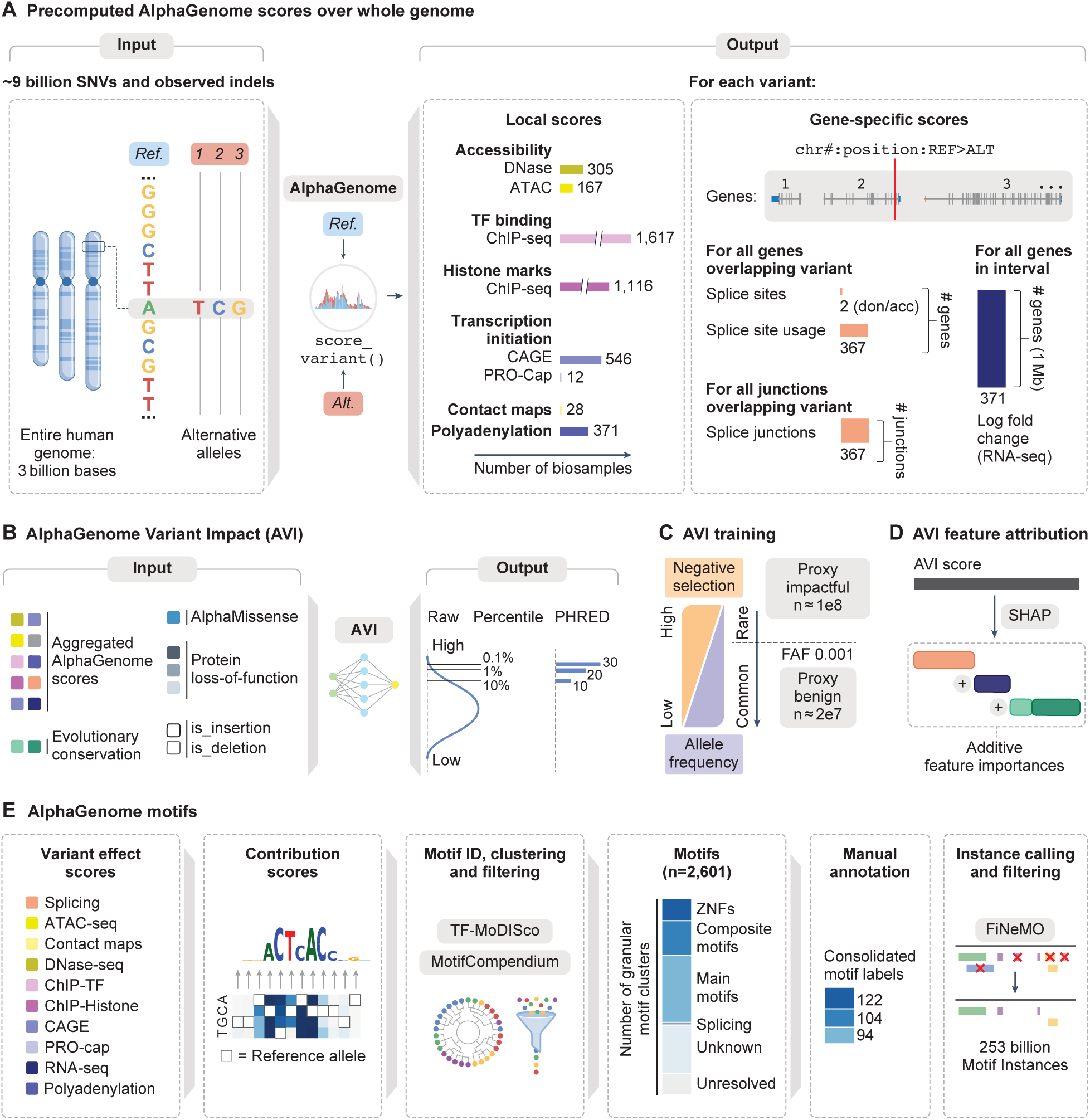
AlphaGenome Atlas: *in silico* mutagenesis of the entire human genome for non-coding variant interpretation. **(A)** AlphaGenome Atlas predictions. Each possible single nucleotide variant and observed indels from different population datasets were scored across 12 AlphaGenome variant scores for all supported biosamples. See also figs. S1 and S2 for more detail. don/acc=splicing donor and splicing acceptor. **(B)** AlphaGenome Variant Impact (AVI) model architecture. AVI integrates AlphaGenome and AlphaMissense predictions, 3 protein loss-of-function annotations from ENSEMBL Variant Effect Predictor (protein termination, stop lost, start lost), two conservation scores (PhastCons 470-way alignments and Zoonomia’s Cactus 241-way), and indel type indicator variables. The predicted raw scores are transformed to a PHRED score based on the score ranking of all SNVs. The PHRED scores of Indels are computed by mapping the indels scores to the quantile curve of the SNV scores. See also fig. S2 for more detail on the AVI model architecture. **(C)** AVI training data. The filtering allele frequencies of the population group max filtering allele frequency from gnomAD v4.1 are used to derive proxy labels to train AVI scores. Variants with allele frequency larger than 0.001 are labeled as proxy benign (label 0) whereas variants with allele frequency smaller than 0.001 are labeled as proxy impactful (label 1). These are proxy labels during training, meaning that the model is trained to assign higher scores for variants with lower allele frequency on average. **(D)** AVI feature attribution. Approximate SHAP feature contributions using expected gradients were computed for each AVI prediction. The reference prediction is an AVI prediction with all input features assigned as 0. These AVI SHAP values are additive, and sum to the raw AVI score (i.e., before PHRED scaling), with positive and negative values indicating each feature’s directional effect. **(E)** AlphaGenome motifs. The ISM contribution scores of AlphaGenome were used as input to TF-MoDiSco for *de novo* motif discovery. TF-MoDiSco-discovered motifs were filtered, clustered, and annotated to obtain a consolidated set of relevant motifs using MotifCompendium. Motifs were manually assigned TF family names, which fell under broad annotation categories: Main (known motifs), Composite (multiple binding sites), ZNF (zinc finger), Unknown (high quality motifs, unknown binders) and Unresolved (lower confidence motifs). The location of motif instances across the genome for all the different modalities and cell types was called with FiNeMo, followed by an additional filtering step to drop low importance motif instances.

**Fig. 2.**
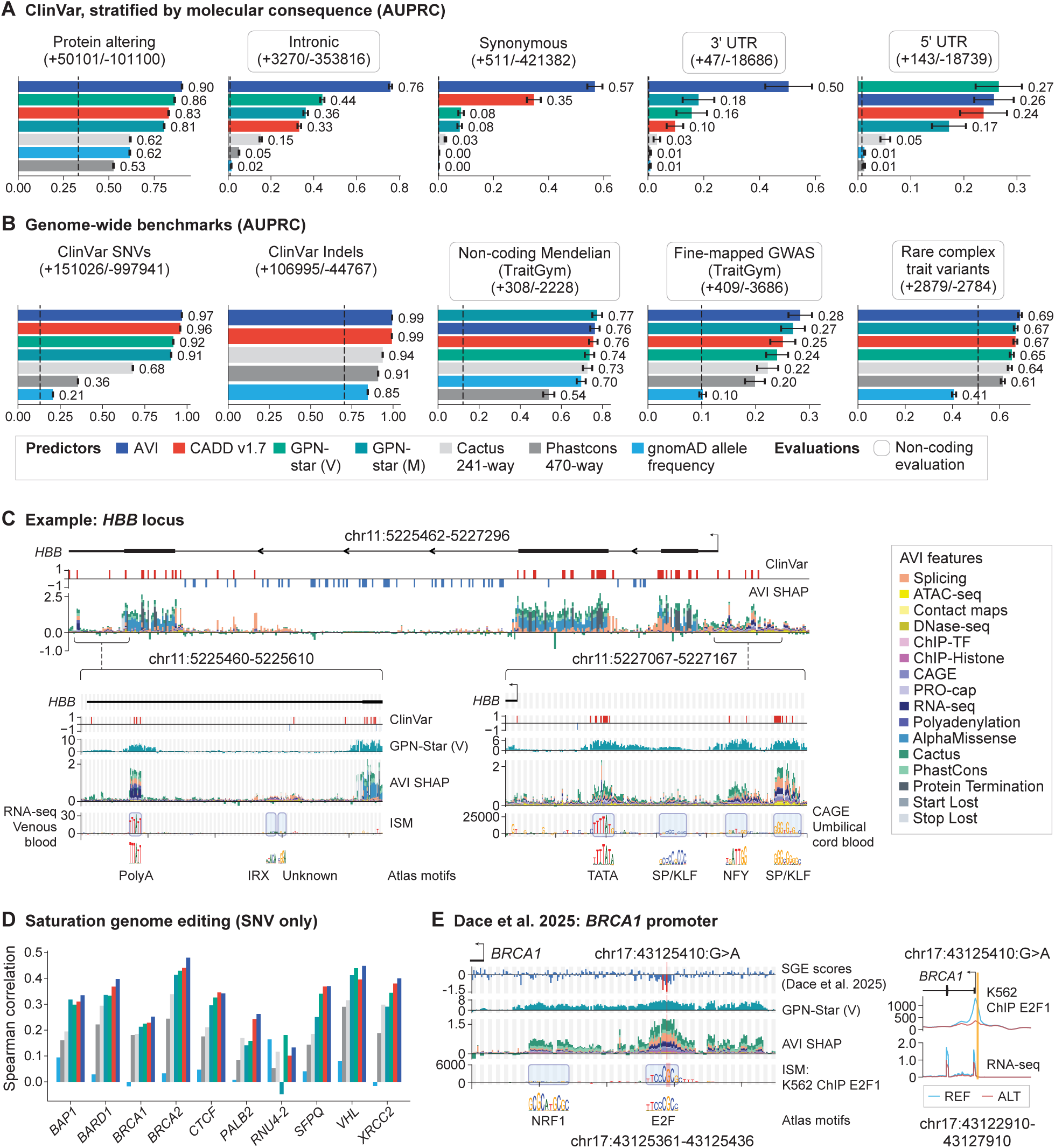
AlphaGenome Variant Impact (AVI): a combined score for genome-wide coding and non-coding variant prioritization. **(A)** Comparison of AVI to other methods for classifying ClinVar pathogenic single nucleotide variants (SNVs) from benign SNVs, stratified by ClinVar variant consequences (AUPRC). Across the figure, error bars indicate 95% confidence intervals from 100 bootstraps, and black dashed lines indicate the value expected for a random classifier. **(B)** Comparison of AVI to other methods across additional genome wide benchmarks including complex traits and rare disease. This includes: ClinVar stratified by SNV versus Indel, TraitGym Mendelian and Complex traits, as well as a benchmark for rare variants associated with complex traits. For more information, see Methods. **(C)** Visualization of AVI feature attributions across the *HBB* locus. Top: The transcript annotation for the *HBB* gene, alongside ClinVar known pathogenic (red) and benign (blue) variants shown above the AVI feature attributions. Both ClinVar and AVI feature attributions are represented at 4 bp resolution by the maximum datapoint within each 4 bp bin. Insets: Expanded locus views of chosen regions of *HBB*, including the 3’ UTR (left) and the promoter (right). GPN-Star (V) is shown for comparison, as well as Atlas motifs and ISM scores for chosen tracks. Grey and white stripes in the insets mark base-pair resolution. For AVI the 3 data points within each stripe represent the 3 alternative bases at allelic resolution. Alternative variants are ordered alphabetically. See fig. S7C for the conversion of AVI SHAP raw scores to AVI PHRED scores for interpretation. **(D)** Spearman correlation between model predictions and experimental saturation genome editing (SGE) assay measurements across 10 genes. Each color indicates one computational method, with model colors the same as shown in **(A)**. Only SNVs from these screens are used in this evaluation. **(E)** Left: An example variant (chr17:43125410:G>A) in the promoter region of *BRCA1* that was measured by SGE (*31*). Negative SGE scores indicate loss of cell fitness as a proxy for *BRCA1* expression; red variants exceed author-defined thresholds for loss-of-function, while blue are under threshold. SGE scores are shown together with AVI SHAP, motif instances and AlphaGenome K562 ChIP-TF Active ISM contributions scores in this region. K562 was chosen as the closest cell line proxy available for the HAP1 cell line used in the original experiment. An Atlas E2F motif is identified in the ChIP-TF E2F1 track overlapping the variant. Grey and white stripes mark base-pair resolution. For SGE and AVI scores the 3 data points within each stripe represent the 3 alternative bases at allelic resolution. Alternative variants are ordered alphabetically. Right: AlphaGenome E2F1 ChIP-TF and RNA-seq (K562) predictions for the reference and alternative sequence.

**Fig. 3.**
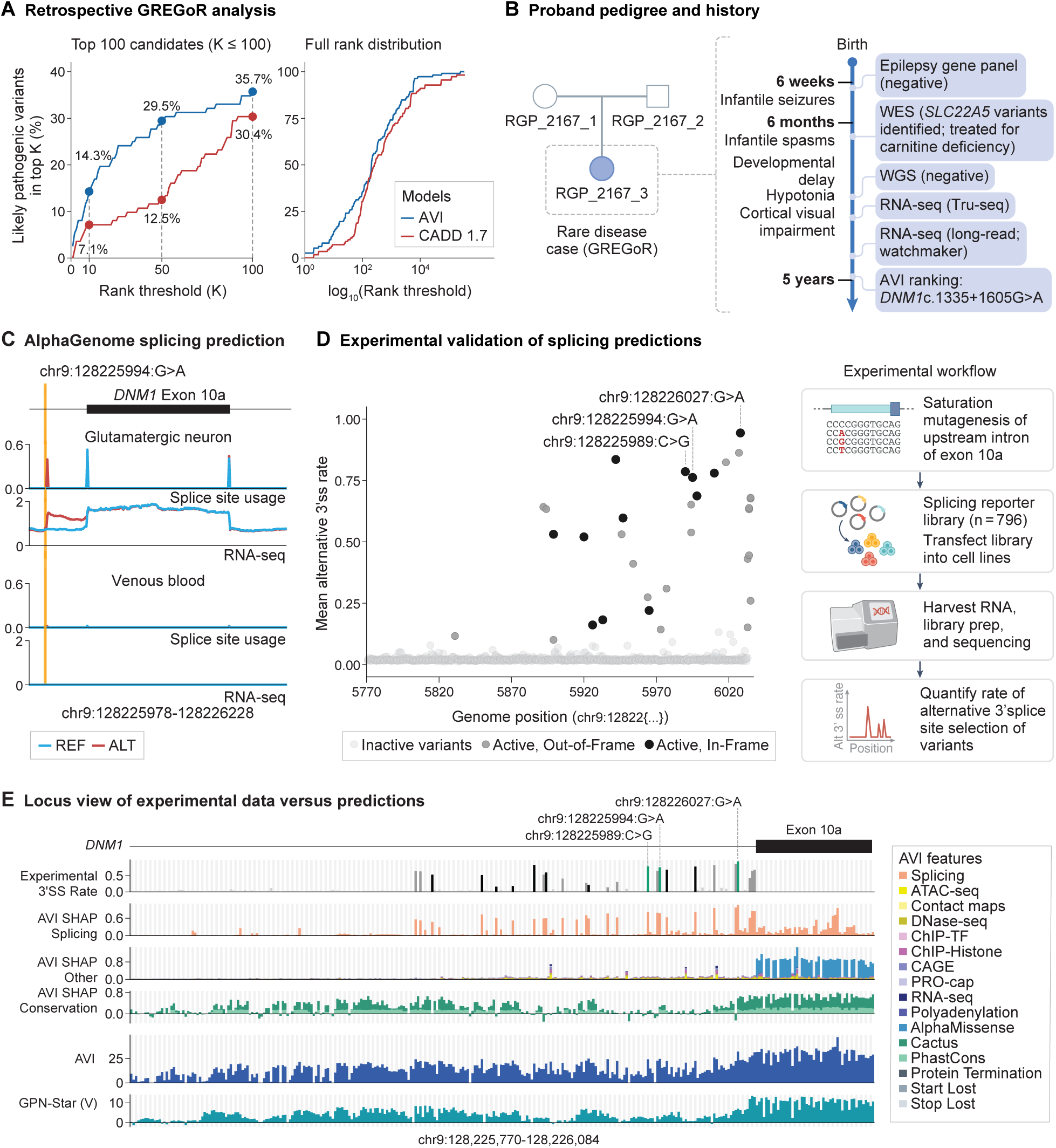
AVI prioritization and functional characterization of a *de novo* deep intronic variant in the GREGoR rare disease cohort. **(A)** Retrospective variant prioritization performance in the GREGoR cohort. AVI and CADD v1.7 were evaluated on 112 likely pathogenic variants from solved cases, ranked against intra-patient background variants. Curves plot the cumulative recall (y-axis) of causal variants captured within the top K prioritized candidates (x-axis). The left panel bounds the evaluation at K = 100; the right panel displays the full rank distribution. **(B)** Pedigree of the proband exhibiting chr9:128225994:G>A along with a case timeline, with clinical phenotypes shown on the left with molecular testing history on the right. **(C)** AlphaGenome track predictions for the reference and alternative allele of chr9:128225994:G>A. The “glutamatergic neuron” biosample was chosen because of its relationship to the epileptic encephalopathy phenotype, while “venous blood” was chosen to illustrate the lack of signal of the tissue specific *DNM1* exon 10a in blood samples. AlphaGenome splice site usage and RNA-seq coverage predictions are shown. **(D)** Left: Summary of experimental results. The y-axis shows the mean alternative 3’ splice site rate computed by first taking the average across replicates, then taking the average across the 5 cell lines used. The x-axis represents the genomic position of the variants upstream of *DNM1* exon 10a. Variants are colored by their activity status and ability to generate in frame (black) and out of frame exon (dark gray) extensions. The three variants known in the literature to cause dominant negative phenotypes are labeled. Right: Schematic of experimental mutagenesis strategy targeting 265 nucleotides upstream of *DNM1* exon 10a. **(E)** Locus view showing the experimental results from **(D)**, overlaid with AVI SHAP breakdowns of splicing, conservation and other, followed by AVI and GPN-Star (V). Each column is an individual variant. The three variants known in the literature to cause dominant negative phenotypes are labeled and highlighted in green.

**Fig. 4.**
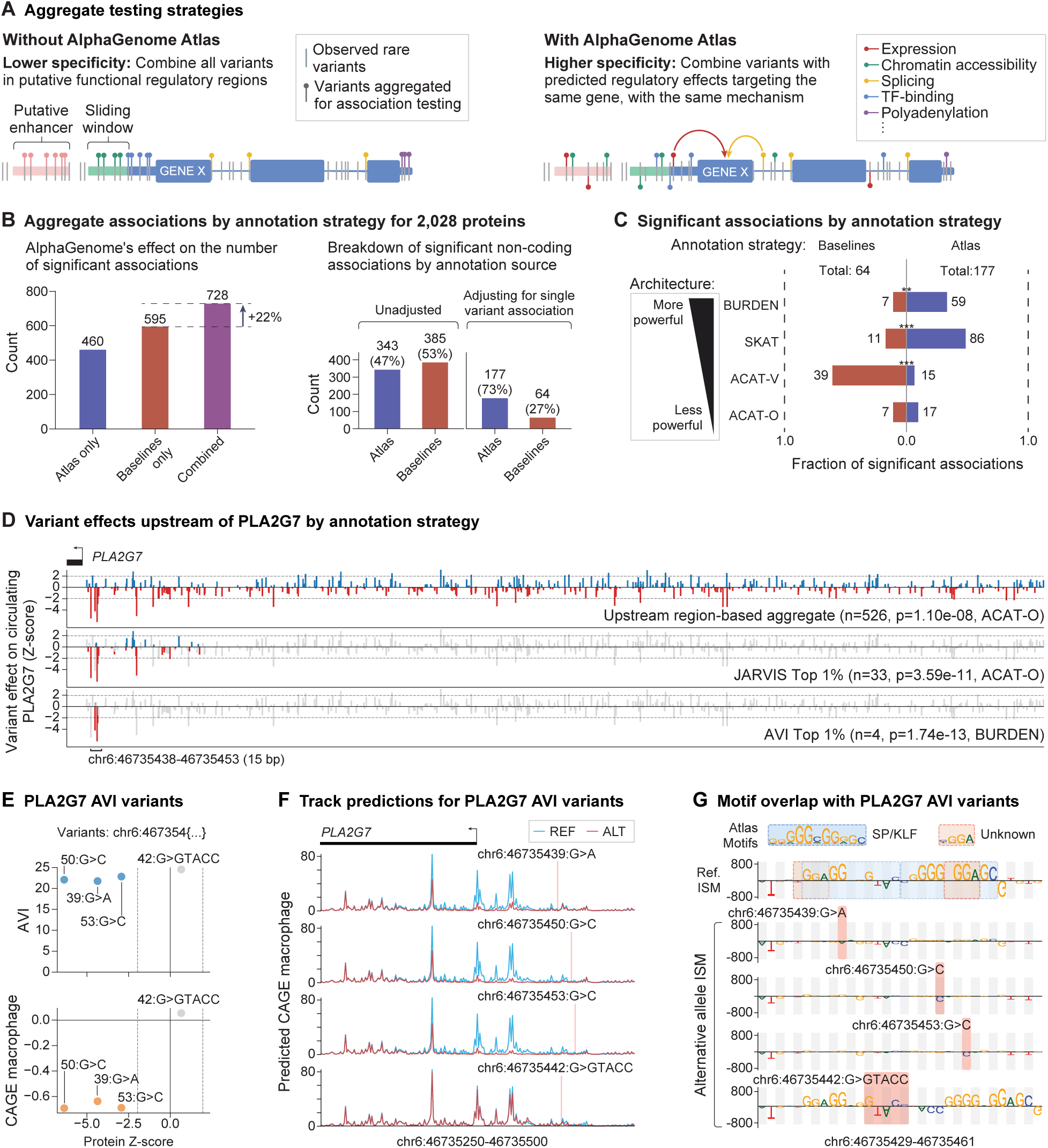
Discovery of non-coding rare variant aggregates associated with protein abundance. **(A)** Illustration of how we define sets of variants (“aggregates”) to use in association tests with, and without AlphaGenome Atlas. In both cases we start with non-coding, rare variants falling in putative regulatory and/or conserved/constrained regions, but with AlphaGenome Atlas we further subset these variants to those with high AVI scores, or high variant scores for different modalities, such as gene expression, chromatin accessibility etc. (see Methods for full details). **(B)** Summary of the impact of using AlphaGenome Atlas for discovery of associations between circulating protein levels (2,028 proteins assayed in the UK Biobank) and aggregates of rare, non-coding variants. In all cases we have adjusted for common (minor-allele-frequency *≥* 1%) single-variant pQTLs from (*49*) for each protein. Left-hand plot: the number of significant independent associations with, and without using Atlas-derived aggregates. Each bar corresponds to the number of significant associations found after effectively completing our conditional analysis (Methods), but with subsets of aggregates obscured (ablation analysis), represented by the x-axis. Right-hand plot: Counts of significant associations before and after additionally adjusting for rare (MAF<1%) single variant pQTLs, broken down by the source of the aggregate. For these counts, we included both non-AG and AG-derived annotations in defining non-coding aggregates, but show the breakdown of annotation source after the conditional analysis step. Baselines: aggregates as defined in (*49*). **(C)** Distribution of statistical association tests for conditionally independent aggregates associated with circulating protein levels. Asterisks indicate statistically-significant differences between the counts of discoveries derived from non-Atlas and Atlas aggregates (two-sided Fisher Exact test; ***= p-value <0.00025; **= p-value <0.0025; * = p-value < 0.0125; thresholds are Bonferroni-corrected for 4 tests). Odds ratios for Atlas counts vs. non-Atlas are BURDEN: 4.1; SKAT: 4.6; ACAT-V: 0.1; ACAT-O: 0.9 (n.s.). Tests are listed in order of decreasing power of their architectures; BURDEN: classical burden test, which assumes per-variant effect-size homogeneity. SKAT: allows effect-size bi-directionality. ACAT-V: allows bi-directionality and only a subset of variants to have a significant effect size. ACAT-O: an omnibus test of BURDEN, SKAT and ACAT-V. **(D)** Example of a rare non-coding variant aggregate constructed upstream of the transcript encoding the circulating protein PLA2G7. The first aggregate is the default upstream region-based aggregate, which selects rare variants in a defined 5 kb window upstream of the transcript. The direction of effect on protein levels associated with the rare variants is indicated in color (red for down, blue for up). JARVIS Top 1% and AVI Top 1% strategies apply a score-based threshold to filter variants from the upstream aggregate (filtered out variants displayed in gray). The number of variants composing each aggregate, their p-value and the statistical test associated with the p-value are shown in facet titles. After thresholding, the AVI aggregate contains four variants in a 15 bp window upstream of *PLA2G7*. **(E)** Scatterplot of AVI and macrophage CAGE-seq predictions versus PLA2G7 protein Z-scores. Each datapoint is one of the four variants identified in the AVI aggregate for *PLA2G7* in panel D. Dashed vertical lines at x=1.96 and −1.96 indicate nominal significance (P < 0.05) thresholds for sign of effect on circulating protein levels. **(F)** AlphaGenome CAGE-seq track predictions for the reference and alternative sequences associated with each variant identified in the AVI aggregate. The genomic interval shown contains the transcriptional start site of *PLA2G7*. **(G)** AlphaGenome Atlas motifs associated with the 15 bp window containing the AVI variant aggregates (as highlighted in **(D)**). Macrophage CAGE-seq Active ISM scores are shown for the reference sequence (top row) versus the four alternative allele ISM (remaining rows). The alternative allele in the sequence is highlighted in red. Two AlphaGenome Atlas motifs (SP/KLF and unknown) are highlighted in the reference sequence.

**Fig. 5.**
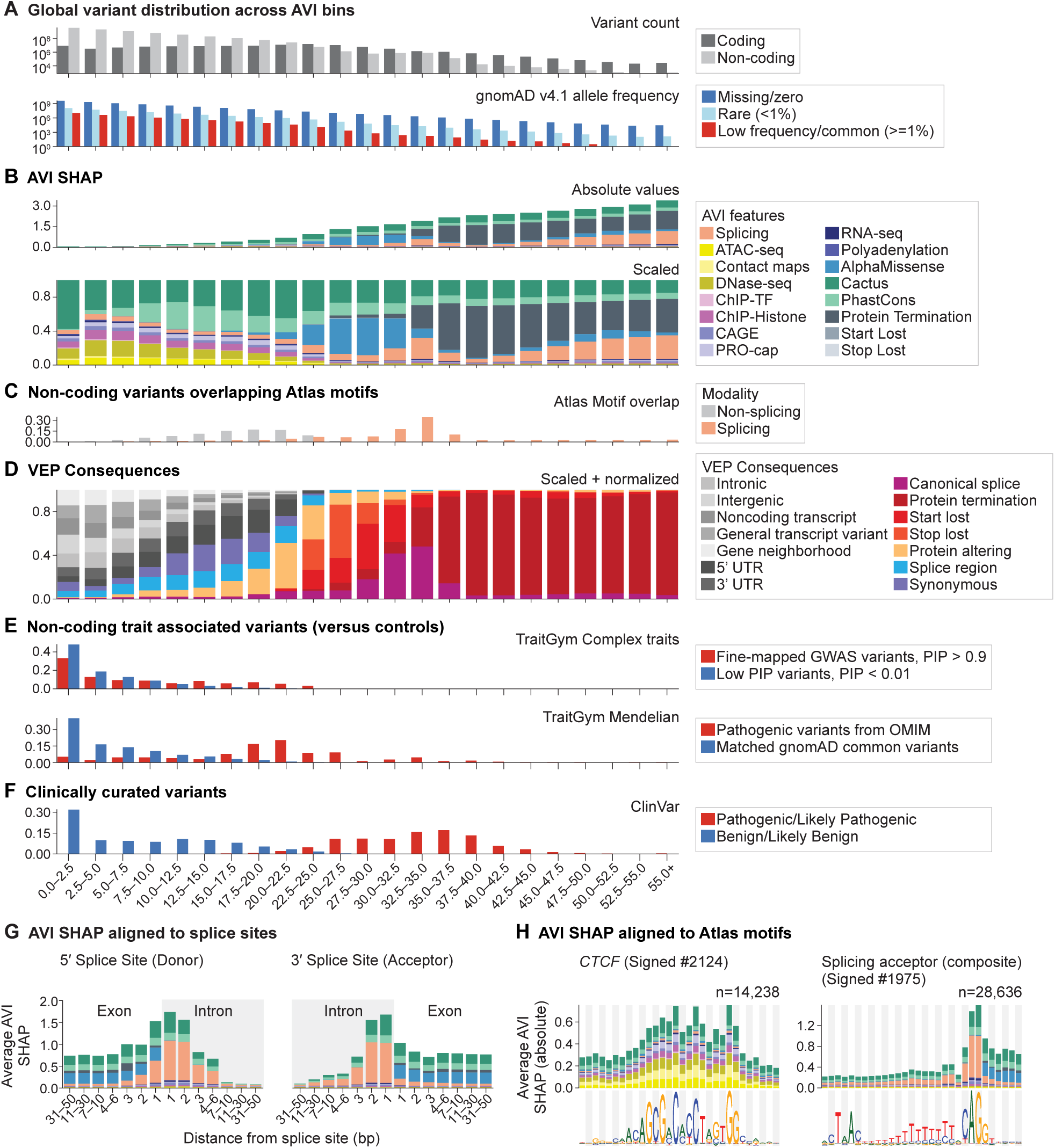
Whole genome analysis of AVI feature attributions. **(A)** Global variant distributions across AVI bins. All hg38 single nucleotide variants are partitioned into 23 bins by their AVI scores (width = PHRED 2.5). Due to the logarithmic transformation of the PHRED scale, variant density decays exponentially across successive bins. Top: Total variant counts per bin, split by non-coding versus coding, with coding as defined as overlapping a coding sequence according to the GENCODE v46 GTF. Bottom: Total counts split by gnomAD v4.1 filtering allele frequency (missing/zero, rare (<1%) and low frequency/common (>=1%)). **(B)** AVI SHAP feature attributions across AVI bins. Top: Mean absolute values of AVI SHAP feature attributions per bin. Bottom: Same as above, but with the Y-axis scaled between 0 and 1, indicating relative fractions of feature attributions per bin. **(C)** Fraction of non-coding variants (as defined in A above) that overlap Atlas motifs across AVI bins. Each variant was evaluated for motif overlap specifically within the AlphaGenome track contributing its largest absolute feature attribution. Motif categories are split by splicing modality and non-splicing modalities. **(D)** VEP consequences across AVI bins. Values are represented as bin-wise variant counts, normalized by the total number of variants in each consequence genome-wide, and scaled to 100% per bin. Multiple consequences per variant were resolved by selecting the highest-severity annotation (truncating/canonical splice > inframe/protein-altering > synonymous/splice region > others). For variants mapping to multiple consequences of equal maximal severity, fractional weights were distributed uniformly among them. **(E)** Non-coding trait-associated variants across AVI bins. Top: Variants from the TraitGym Complex evaluation, in which positive variants are fine mapped GWAS variants with posterior inclusion probability (PIP) > 0.9, and negative variants are variants with PIP < 0.01. Bottom: Variants from TraitGym Mendelian evaluation, in which positive variants are non-coding pathogenic variants from Online Inheritance in Man (OMIM), and negative variants are matched gnomAD common variants. Related to Fig. 2B. **(F)** Clinically curated variants from ClinVar across AVI bins. Positive variants are Pathogenic / Likely Pathogenic, and negative variants are Benign / Likely benign. Related to Fig. 2B. **(G)** Stacked bar plot of average feature attributions of SNVs„ binned by distance to the nearest splice boundary (exon-intron or intron-exon, GENCODE V46). **(H)** Per-position AVI feature attributions for example motifs. For each motif, genome-wide instances were retrieved, and negative strand instances were reverse-complemented. Top row: per-nucleotide AVI feature attributions, illustrating the spatial distribution of predicted variant effects across the motif footprint. Bottom row: contribution weight matrix (CWM) sequence logos. Left: CTCF motif. Right: Splicing acceptor (composite).

**Fig. 6.**
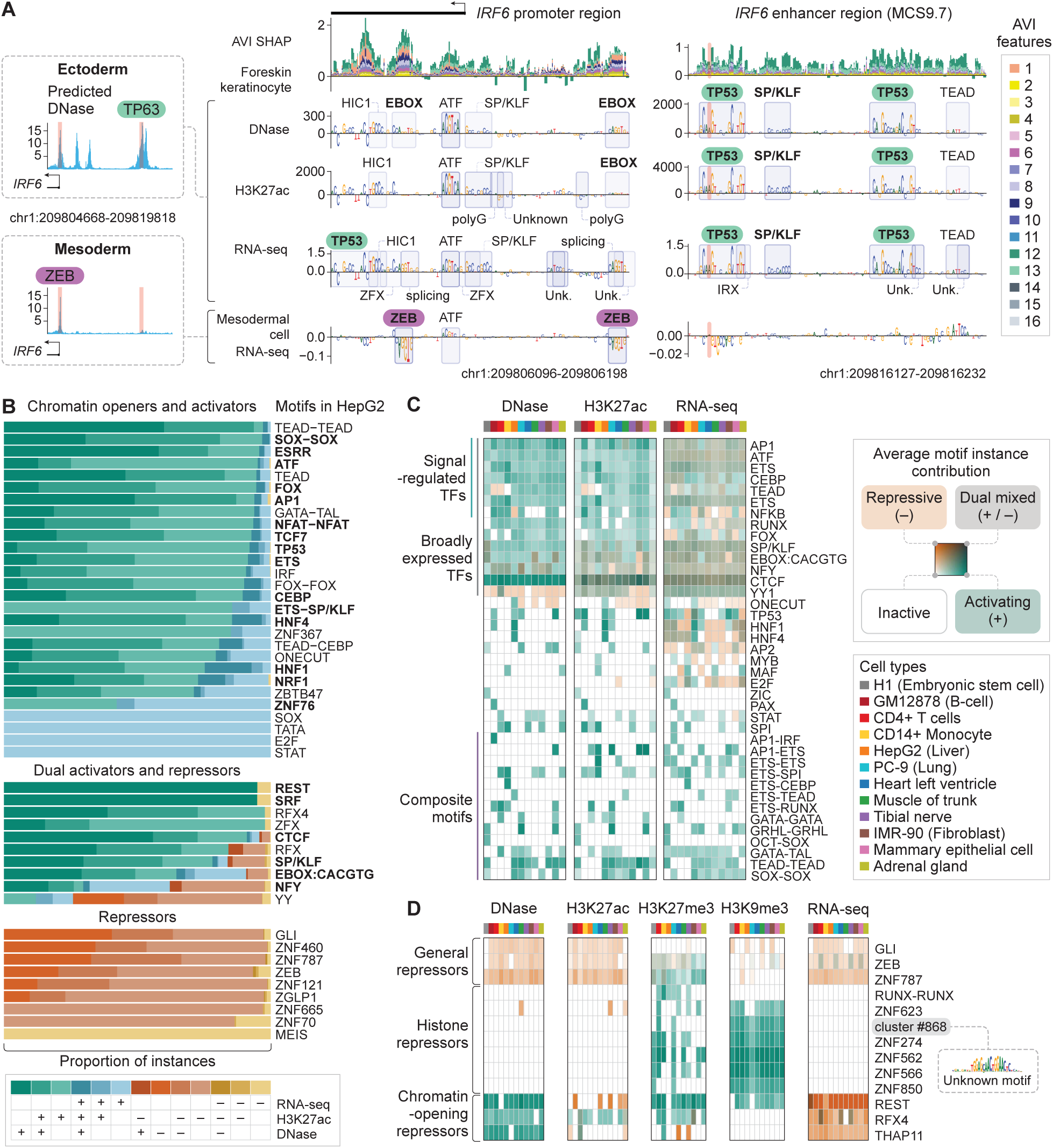
Atlas displays cell-type-specific motif maps and allows classification of motifs based on their gene regulatory function. **(A)** AlphaGenome Atlas motif maps at the *IRF6* locus highlighting the promoter and disease relevant upstream enhancer. Left: AlphaGenome predicted DNase tracks for the 15 kb region upstream of the *IRF6* promoter. Two tracks are shown: one representing the ectoderm (‘foreskin keratinocyte’) where *IRF6* is expressed, and another representing the mesoderm (‘mesothelial cell of epicardium’) where *IRF6* is silenced. Cell-type specific TFs known to regulate the core promoter and MCS9.7 enhancer (highlighted by vertical shading) are shown above their corresponding regulatory elements. Middle and right: AVI feature attributions and motif maps with Active ISM scores are shown for ‘foreskin keratinocyte’ where *IRF6* is expressed (as seen in the DNase, H3K27ac, RNA-seq modalities) and for ‘mesodermal cell’ where *IRF6* is inactive (as seen in the RNA-seq modality). The ∼100 bp core promoter region (chr1:209,806,096–209,806,198, middle) shows repressive ZEB motifs in mesoderm, while the enhancer region MCS9.7 (chr1:209,816,127–209,816,232, right) shows two activating TP63 motifs (TP53 class) in the foreskin keratinocyte, the first of which is disrupted in individuals with cleft lip and palate (*56*, *57*). Legend for AVI features: 1. Splicing, 2. ATAC-seq, 3. Contact maps, 4. DNase-seq, 5. ChIP-TF, 6. ChIP-Histone, 7. CAGE, 8. PRO-cap, 9. RNA-seq, 10. Polyadenylation, 11. AlphaMissense, 12. Cactus, 13. PhastCons, 14. Protein Termination, 15. Start Lost, 16. Stop Lost. **(B)** Modality analysis of motifs contributing to RNA-seq predictions in HepG2 cells. Motif labels are on the right, with those found by MPRA analysis in HepG2 cells (*60*) in bold (fig. S15A). For each motif, the stacked barplot represents the proportion of motif instances that were mapped across select modality combinations of DNase, H3K27ac and RNA-seq, either with positive (+) or negative (-) effects as shown below (see fig. S15B for all combinations). Each motif instance at a genomic coordinate is assigned to a single category. Most motifs, including all MPRA motifs, have instances in multiple modalities with distinct contributions towards chromatin opening and transcription, but the effects from the modalities are often clearly in one direction, allowing motifs to be classified into belonging to TFs that are activating or repressing in this cell type. Some motifs like CTCF have dual roles towards activation and repression. **(C)** Global analysis of selected motifs (y-axis) across selected cell types and modalities (x-axis). The heatmap shows the average positive and negative motif instance contribution for the DNase, H3K27ac, RNA-seq modality in 12 cell types representing different lineages. The color represents the overall effects as activating (green), repressive (orange), mixed (dark), and inactive (light or white). Signal-responsive motifs have mostly positive effects on accessibility and histone acetylation but their effect on gene expression (RNA-seq) is often mixed. Cell-type-specific motifs, many of which are composite motifs, often promote gene expression through multiple modalities. See **Data S3** for a more comprehensive heatmap. Cell type legend full names: H1; GM12878; CD4-positive, alpha-beta T cell; CD14-positive monocyte; HepG2; PC-9; heart left ventricle; muscle of trunk; tibial nerve; IMR-90; mammary epithelial cell; adrenal gland. **(D)** Classification of selected repressor motifs (y-axis) based on their effects on the modalities shown above (DNase, H3K27ac, RNA-seq) and repressive histone modifications (H3K27me3 and H3K9me3) across cell types (x-axis). The cell types and color legend for motif effects are the same as in **(C)**. General repressor motifs such as GLI were those with broadly repressing effects on DNase, H3K27ac, RNA-seq. Histone repressor motifs did not affect these modalities but promoted H3K27me3 and/or H3K9me3. H3K9me3 is often promoted by zinc finger binding motifs such as ZNF274 and included unknown motifs (e.g. cluster #868). Chromatin-opening repressor motifs such as REST positively affect accessibility but repress gene expression. See **Data S3** for a more comprehensive heatmap.

### Whole genome *in silico* mutagenesis with AlphaGenome Atlas enables variant impact scores and comprehensive motif maps

To construct a comprehensive map of non-coding variant effects, we calculated the difference between reference and alternate allele predictions for all ∼9 billion SNVs in hg38 using AlphaGenome (Fig. 1A, Methods). We also scored >100 million indels observed in major biobanks (gnomAD, UK Biobank, All of Us) using a shift augmentation strategy (*11*) (fig. S1, Methods). We used assay-specific scoring strategies to reduce these reference and alternative prediction differences to scalar values (Table S1) (*9*). For example, we scored chromatin accessibility effects focusing on the 501 bp centered on the variant, while for polyadenylation, we scored the maximum log-fold change of isoform ratios across polyadenylation sites based on predicted RNA-seq coverage. Additionally, we computed active allele scores for a relevant subset of scorers to capture the absolute activity level of the variant’s regulatory element. As a result, each variant is associated with an average of 27,000 experiment-specific scalar predictions (∼15,000 excluding active allele predictions), spanning hundreds of biosamples, each assigned to tissue- or cell-type specific ontology terms (Methods).

Next, we used the precomputed AlphaGenome scores as input features for a neural network model to derive AlphaGenome Variant Impact (AVI) scores for the prioritization of genetic variants (Fig. 1B, fig. S2, Methods). We grouped AlphaGenome predictions into ten different features based on the assay modality (DNase-seq, ATAC-seq, TF binding ChIP-seq, histone mark ChIP-seq, CAGE, PRO-Cap, RNA-seq, polyadenylation, splicing, and contact maps), which we reduced to the per-variant maximum predicted effect across tissues (Table S1). We also included four coding features consisting of AlphaMissense and three VEP protein termination features (‘stop gained or frameshift’, ‘start lost’, and ‘stop lost’) (*3*); two conservation features (PhastCons 470-way alignments and Zoonomia’s Cactus 241-way) (*12*, *13*); and two indel indicator features (‘is insertion’ and ‘is deletion’). AVI was trained to discriminate between observed human variants that have a filtering allele frequency above or below 0.1%, respectively representing proxy neutral versus proxy impactful variants (*14*) (Fig. 1C). The raw model predictions were then converted to PHRED scores (PHRED 10 indicates top 10% of predictions, 20 indicates top 1%, etc.).

This minimal 18-feature set was intentionally chosen to enable more precise feature attribution that can often become confounded in larger feature spaces. We used the SHAP framework to compute the additive contribution of each individual input feature to the AVI score (Fig. 1D, fig. S3, Methods). For each variant, the 18 associated AVI SHAP contributions sum to the raw AVI score for that variant. Because each feature corresponds to a specific modality, this effectively decomposes each AVI score into a mixture of interpretable effects. For any variant whose AVI score is driven by an AlphaGenome feature, its precomputed scores and AlphaGenome track predictions can be further explored to resolve cell type specificity and regulatory function.

To interpret the motifs underlying AlphaGenome and AVI scores, we identified, mapped, and annotated *cis*-regulatory DNA motifs from precomputed ISM contribution scores (Fig. 1E, Methods). To effectively perform motif identification genome-wide, we defined relevant genomic regions in a track-specific way and ran TF-MoDISco (*15*) on the precomputed ISM scores. The ∼900M discovered patterns were then filtered and clustered using MotifCompendium (*16*) into 2,601 motifs (Fig. 1E, fig. S4, Methods). Since homologous TFs and even unrelated TFs can have very similar motifs, we manually assigned these clusters broad motif labels, resulting in consolidated motif labels for 94 main TFs and 122 zinc finger TFs (Fig. 1E). Additionally, we identified 464 composite sequence patterns where two main motifs are separated by a fixed distance (*17*, *18*), representing 104 distinct motif combinations. The motifs overall showed high similarities to known motifs in the JASPAR database and ENCODE GRAMMAR (*16*, *19*) (fig. S4C,D, fig. S4B for an illustration of motif similarity metric values).

We next developed a strategy to identify locations of motifs (motif instances) genome-wide using FiNeMO (*20*) (Fig. 1E). Since ISM scores measure the differences between reference and alternate allele predictions, they are not in scale with the experimental signal nor have a defined significance threshold. To adjust for this, we introduced a filtering step and optional rescaling (fig. S5, Methods), allowing us to map motif instances at comparable rates as other approaches leveraging deep learning (*15*, *17*, *21*, *22*). In comparison to ChromBPNet (*22*) and accessibility footprinting (*23*), AlphaGenome’s motif instance calls demonstrated comparable results on recall of UniBind motif maps (*24*) (fig. S6A). Filtering and scaling motif maps resulted in overall higher overlap with annotated cCREs (*4*) (fig. S6B). For three TFs, we experimentally validated the mapped motif instances by performing high-resolution ChIP-nexus binding experiments (*25*) in the HepG2 cell line (fig. S6C), revealing consistent binding footprints on the mapped instances but not on those mapped only by traditional sequence similarity scores (*26*). Furthermore, we experimentally validated that mapped motif instances were cell-type-specific using CTCF as an example (*27*, *28*) (fig. S6D-E). Thus, we obtained a genome-wide map of annotated motif instances across each modality and cell type, allowing AlphaGenome Atlas to provide a direct link between AVI scores and underlying *cis*-regulatory motifs.

### State-of-the-art interpretable genome-wide variant prioritization with AVI

A robust variant prioritization score must accurately resolve variant impact across diverse genetic architectures, spanning monogenic diseases to complex traits, and common polymorphisms to rare variants. AVI achieved state-of-the-art performance across benchmarks reflecting this diversity, with particularly strong gains on non-coding variants. For clinically-asserted pathogenic versus benign SNVs from ClinVar (*29*), AVI markedly improved over next-best global variant interpretation models in intronic (AUPRC 0.76 vs. GPN-Star-V’s 0.44), synonymous (0.57 vs. CADD v1.7’s 0.35), and 3’ UTR categories (0.50 vs. GPN-Star-M’s 0.18), while also advancing on protein-altering variants (0.90 vs. GPN-Star-V’s 0.86) (Fig. 2A). GPN-Star-V outperformed AVI on 5’UTR ClinVar SNVs (0.27 vs. 0.26) (Fig. 2A). On the ClinVar test data with both SNVs and indels, AVI outperformed all alternative approaches supporting indel scoring (fig. S7A). When considering all ClinVar test SNVs and Indels separately, AVI improved over alternative methods on both (Fig. 2B). These gains, albeit smaller, extended to complex-trait fine-mapping, where AVI better distinguished putatively causal from non-causal GWAS variants (TraitGym Complex, AUPRC 0.28 vs. 0.27), (Fig. 2B) (*30*), and to a rare-variant benchmark linking variants to complex traits (AUPRC 0.69 vs. GPN-Star-M’s 0.67) (Methods, Fig. 2B, fig. S7B). On non-coding monogenic disorder variants (TraitGym Mendelian), AVI ranked second to GPN-Star-M (AUPRC 0.76 vs. 0.77) (Fig. 2B).

Beyond these performance gains, AVI feature attributions enable allele-specific interpretations of variant molecular effects. As exemplified by the *HBB* locus, which is well annotated by classified variants in ClinVar, AVI feature attributions align with the known components of the transcript, with splicing features attributed most strongly at core splicing and deep intronic regions, RNA-seq and chromatin accessibility at upstream core promoter regions, and AlphaMissense/protein termination features aligned to exonic regions (Fig. 2C). These patterns hold at allelic resolution, with splicing feature attributions aligning to high AVI deep intronic variants that are also clinically pathogenic, RNA-seq / CAGE feature attributions aligning to pathogenic promoter variants known to affect a CACCC box motif element, and polyadenylation feature attributions to pathogenic polyA motif variants (Fig. 2C). Notably, the AlphaGenome motif resource reports motif instances aligning with both of these known elements (Fig. 2C).

In addition to genome-wide benchmarks, we evaluated AVI against saturation genome editing (SGE) experiments, which measure the functional impact of every possible single-nucleotide variant across defined genomic regions. Across ten held-out screens covering clinically relevant genes (Methods), AVI achieved the highest Spearman correlation with measured variant effects in eight (Fig. 2D, fig. S7D) and achieved the best AUPRC considering all experiments merged (fig. S7E, 0.668 vs. CADD v1.7’s 0.647 on SNV’s only, fig. S7F, 0.759 vs. CADD v1.7’s 0.653 when including indels). Within saturation genome editing screens, we also observed allele-specific interpretability of *BRCA1* promoter variants identified in experimental mutagenesis (*31*) that disrupt an E2F motif (*32*) leading to decreased *BRCA1* expression (Fig. 2E). These variants had high AVI scores and interpretable AlphaGenome-driven feature attributions, with an E2F motif instance called on the K562 E2F1 ChIP-seq track that is ablated in the presence of the alternative allele, alongside decrease in expression of the *BRCA1* gene (Fig. 2E). Additional variant interpretations are shown in fig. S8.

We performed model ablations to assess the necessity of the 18 features chosen as input features to AVI (Table S2). Removing all protein features yielded significant performance drops on coding-variant skewed evaluations such as ClinVar and saturation genome editing. Conversely, ablating the core AlphaGenome features degraded performance across non-coding and regulatory evaluations. During model development, we evaluated numerous additional features, including extended VEP consequences and CADD-inspired features, which did not meaningfully affect model performance when included. Notably, although the AVI model was trained without AlphaMissense scores for indels, we found that supplying approximations of these scores during inference improved performance on coding indels and reduced reliance on the variant-info features (Table S2, Methods). Ultimately, only 18 features were required for robust performance across a diverse set of evaluations. This is a substantial reduction in features compared to CADD v1.7, which uses over 150 features.

### AVI prioritization helped resolve a rare disease case and functionally characterizes deep intronic variants in *DNM1*

We applied AVI to data provided by the GREGoR Consortium, whose aim is to resolve currently unexplained rare genetic disorders (*33*). Rare disease cases can remain unsolved due to time constraints attempting to evaluate an average of 4-5 million variants per individual, the majority of which will be benign or variants of uncertain significance (VUS) (*34*). Retrospectively analyzing past solved cases, including both SNVs and indels, AVI ranked the known likely pathogenic and pathogenic variants among all variants in each patient more highly than CADD v1.7, with a recall of 29.5% compared to CADD v1.7’s 12.5% when considering the top 50 variants ranked by either method (Fig. 3A). When considering only variants filtered by gnomAD allele frequency of 0.001, the recall at top 50 variants increased to 74.3% and 61% for AVI and CADD v1.7, respectively (fig. S9A). Next, we prioritized variants in unsolved rare disease cases in the cohort by systematically ranking and analyzing all small *de novo* variants in 814 individuals with parent and proband genomes available.

For a proband with epileptic encephalopathy (Fig. 3B), the top ranked variant by AVI was a heterozygous noncoding VUS in intron 10 of *DNM1* (chr9:128225994:G>A, HGVS: NM_004408.4:c.1335+1605G>A) affecting a brain-specific transcript isoform encoding for the dynamin protein. This variant is 5’ of exon 10a, which is exclusively present in brain-specific transcripts and absent from the MANE Select transcript (*35*). Of the variant’s AVI PHRED 24.7 score, 69% was directly attributed to AlphaGenome’s splicing feature and the precomputed scores pointed to altered splice site usage in the ‘glutamatergic neuron’ biosamples. Prediction of RNA-seq track coverage of the reference and alternative alleles showed that the variant creates a brain-specific cryptic splice acceptor site resulting in a 13 amino acid (aa) in-frame extension of exon 10a (Fig. 3C). Predictions in the ‘venous blood’ biosample showed negligible expression of exon 10a, explaining why previous RNA-seq from blood draws were inconclusive.

To probe the feasibility of this being a pathogenic variant, we performed a literature search and found that two recently published case studies showed the same brain-specific 13 aa extension of DNM1 in neurodevelopmental disorders (*36*, *37*). We also found two other nearby intronic variants resulting in a longer 15 aa (*38*) and shorter 2 aa (*39*) extensions, both of which were recapitulated in the AVI and AlphaGenome predictions (fig. S9B). Mechanistically, the first amino acid of exon 10a (R399) is critical for tetramerization of dynamin, and these in frame insertions could exert a dominant negative effect by poisoning the wild type pool of functional DNM1 in the brain (*39*). This proposed mechanism is consistent with the fact that all clinical cases are heterozygous.

To corroborate and extend the pathogenic mechanism within this intron of *DNM1*, we used AlphaGenome to search for additional variants that would disrupt this transcript and observed other variants predicted to cause in-frame exon extensions of various lengths (fig. S9C). We then performed high-throughput experiments in which the 265 nucleotides upstream of exon 10a were mutagenized and assayed in a minigene reporter assay across 5 cell lines (*40*) (fig. S9D, Table S3, Data S1 and Methods)). We identified 12 intronic variants that led to in-frame exon extensions, including all three of the likely pathogenic variants we and others have reported (Fig. 3D, Table S4). Compared to the experimental data, both the AlphaGenome merged splicing score (Methods, AUPRC: 0.943) and the AVI splicing feature attributions (AUPRC: 0.943) recapitulated experimental variant classifications as well as the lengths of the observed exon extensions. SpliceAI (*41*) and Pangolin (*42*) also performed well at this task (AUPRC: 0.940 and 0.939, respectively). Despite the high performance of the SpliceAI model for this region, the SpliceAI precomputed scores do not contain predictions around *DNM1* exon 10a, explaining why this variant was missed previously. Finally, both CADD v1.7 (AUPRC: 0.289) and conservation based methods (e.g. GPN-Star (V) with AUPRC: 0.348) failed to capture these splicing variants, most of which are not conserved (Fig. 3E, fig. S9E).

Together, this evidence was sufficient to recommend Likely Pathogenic classification of chr9:128225994:G>A. Furthermore, the out-of-frame variants we identified could be included in prospective classification of the recessive version of this disorder (*43*) (Data S1). In the long term, we hope that improving the prioritization of non-coding variants will help pinpointing the causal variant for individuals with similar hard-to-solve conditions.

### AlphaGenome Atlas features power discovery of rare, non-coding variants associated with population-scale analyses of circulating protein levels

Rare non-coding variation is not only important for interpreting rare disease cases, but also for understanding population-scale phenotypes. Large-scale whole-genome sequencing (WGS) cohorts (e.g., UK Biobank [UKB], N=490k; All of Us v9, N=535k) offer an unprecedented opportunity to study how rare non-coding variation influences human traits. Genotype-phenotype associations that aggregate rare variants under a shared molecular hypothesis (*44*, *45*) have been successful for protein-coding variants (*46*, *47*). This success relies on the predictable functional consequences of coding variants (e.g., missense, loss-of-function). However, extending this framework and success to the non-coding genome remains challenging, due to the sheer scale (∼1 billion non-coding variants in UKB) and complexity of regulatory genetic code, which is complicated by diverse regulatory mechanisms, cell type specificity, bi-directional effects on expression, and functional redundancy. Existing efforts often rely on generic regional annotations (e.g., “conserved” or “promoter”), which dilute statistical power by mixing causal variants with those having heterogeneous or no effects. While flexible statistical association tests like ACAT (*48*) can help to accommodate effect-heterogeneity, rare variants remain largely absent from eQTL and experimental data, making *in silico* predictions essential to bridge this gap.

We reasoned that application of AlphaGenome AVI and precomputed scores from the modalities (referred to together as Atlas features for brevity) would improve the discovery and mechanistic interpretation of rare variant non-coding genetic loci. As exemplar traits, we studied circulating levels of 2,028 proteins across 54,189 UKB individuals (fig. S10A). For each protein under consideration, we applied the rare-variant aggregate (per-variant minor-allele-frequency (MAF) < 0.1%) association pipeline described in (*49*) (Methods), but filtered aggregated non-coding variants within and around the protein-coding gene (extending 1 Mb either side of the transcript) to those in the top 1% of the various Atlas features (Fig. 4A; Table S5 & Table S6; Methods). This not only produces aggregates of variants directly predicted by AlphaGenome to regulate the gene’s expression (RNA-seq) but also aggregates selected through other modalities such as chromatin accessibility, since these also have the potential to regulate gene expression. We put these results in direct competition with analyses of the proteins and variant annotations without AlphaGenome Atlas feature filtering, as reported in (*49*).

We used four aggregate association testing frameworks, ordered from most to least powerful: 1) additive (burden), which assumes variant-effect homogeneity, 2) SKAT, which allows bi-directional variant effects, 3) ACAT-V, where a subset of variants may have no effect on the trait, and 4) ACAT-O, an omnibus test capturing a mixture of the three prior architectures. To isolate the effects of our rare non-coding aggregate associations, we adjusted our association analyses for common pQTLs (MAF>1%) and protein-coding variants (Methods). Next, to ensure statistical independence between rare non-coding aggregates, we performed a stepwise conditional selection procedure for each protein, whereby the variants in the aggregate with the smallest p-value are conditioned upon until no study-wide significant aggregates remain. *We highlight that our results are based on slightly out-dated releases of Atlas, due to the temporary downtime of the UK Biobank Research Analysis Platform (RAP), which we will resolve as soon as it is restored. However, we expect minimal impact on our overall findings (Supplementary Note)*.

Using aggregates filtered on Atlas features yielded a 22% increase in discovery rate for conditionally independent rare non-coding variant aggregates (Data S2.1; Fig. 4B). Furthermore, when we isolated for loci not explained by known low-frequency and rare (MAF<1%) non-coding single-variant pQTLs from (*49*), 177/241 (73%; Data S2.2) of aggregates that remained statistically significant were discovered using Atlas features, meaning Atlas-aggregates were more likely to contain multiple causal rare non-coding variants than non-Atlas aggregates (Fig. 4C). Non-Atlas aggregates, which by definition contain more variants, were thus more likely to either contain a rare pQTL by chance, contain a single causal variant, or contain variants correlated with rare pQTLs but which were themselves not causally associated (84% eliminated, compared to 55% for Atlas aggregates).

The higher density of causal variants in Atlas aggregates is likely because modality-specific filtering homogenizes the variant effect on protein levels as compared to baseline regional priors. Notably, 73% of Atlas-based associations were captured by modality-specific variant filtering. The remaining were captured by AVI alone (fig. S10B), suggesting that features external to AlphaGenome which contribute to AVI, such as conservation, are also valuable. Non-Atlas aggregates also suffered from statistical signal dilution, forcing reliance on the more-lenient ACAT-V association test (61% of non-Atlas signals; Fig. 4C), which suppresses statistical noise and null variants but sacrifices interpretability. Conversely, Atlas aggregates were significantly enriched for burden (OR: 4.1, *P* = 5.0e-4) and SKAT tests (OR: 4.6, *P* = 7.0e-6) compared to non-Atlas aggregates (Fig. 4C). Following the underlying assumption of the burden and SKAT frameworks, this architectural shift further indicates that Atlas aggregates contain a higher proportion of causal variants.

The improved discovery and interpretability of rare non-coding loci through Atlas is illustrated by the upstream region of the *PLA2G7* gene, which encodes a macrophage-secreted platelet activating factor acetylhydrolase. *PLA2G7* is down-regulated in humans undergoing caloric restriction and has been linked to lifespan via its metabolic impact (*50*). Without any variant-level filtering, the rare non-coding aggregate upstream of *PLA2G7* consisted of 526 rare variants and was not significantly associated with altered levels of circulating PLA2G7 (min *P* = 1.10e-8 in the ACAT-O framework) (Fig. 4D). In (*49*), without Atlas, this aggregate was significant in an ACAT-O framework (*P* = 3.59e-11) after filtering to the top 1% of JARVIS scores (*51*), consisting of 33 variants. Compared to these annotation strategies, filtering to the top 1% of AVI scores had increased significance in a burden framework (*β* = −1.73 SD, *P* = 1.74e-13) and consisted of only four variants, three of which exhibited negative effect sizes on circulating levels of PLA2G7 (*P* < 0.05) (Fig. 4D).

These three SNVs were strongly predicted to reduce transcription in macrophages (predicted macrophage, Fig. 4E-F). Notably, they fall within a 15 bp window overlapping a G-rich region containing two putative SP/KLF motifs. All three SNP alleles disrupt a reference guanine (G) and strongly attenuate the contribution scores across the entire region, suggesting a coordinated activity of the two sides of the motif (Fig. 4G). The fourth variant, an insertion, did not disrupt a G but instead inserted 4 nucleotides between the two SP/KLF motifs. This was predicted to have minimal predicted effect on CAGE macrophage signal (Fig. 4F), corroborated by the proteomics data, in which the variant did not have evidence to support a reduction in circulating PLA2G7 levels (*P* > 0.05, Fig. 4E). The sensitivity of the long G-rich pattern to specific guanine disruption but not to alteration of their spacing, and its proximity to the transcriptional start site, is suggestive of a cis-regulatory element, perhaps a G-quadruplex structure (*52*). Pending functional validation, we propose that this element promotes PLA2G7 transcription and consequently reduces protein expression when disrupted.

Finally, we applied our methodology to three complex human traits: standing height, body-mass index (BMI) and glycated haemoglobin (HbA1c) (fig. S10C), using only AlphaGenome-derived scores and not yet AVI (Supplementary Note). We identified 43 conditionally independent non-coding aggregates after additionally adjusting for rare variants associated with each phenotype (*53*), with 72% driven by AG filtering (Methods, Data S2.3). For example, in the 3’ UTR region of *PRRC2B*, an RNA-binding protein linked to murine adiposity, an aggregate filtered on the ChIP-TF modality achieved SKAT significance with BMI (*P* = 3.91e-10), while the unfiltered regional aggregate was null (*P* = 0.197). Despite the inherent challenge of replicating rare-variant associations in other cohorts, we found that among the 25/31 of associations with sufficient variant counts in the All of Us v8 WGS cohort (N = 415,000), 4 nominally replicated (*P* < 0.05), though not at Bonferroni significance.

In summary, by computationally filtering non-coding genomic regions to mechanistically homogeneous variant subsets, we have demonstrated that AlphaGenome Atlas is a powerful new tool for improving the rates of discovery of, and molecular insights into, associations between rare non-coding variation and human traits. Such improvements are likely due to Atlas-derived annotations identifying putatively functional non-coding variants with higher specificity than alternative approaches - both in terms of the variants included in the aggregate, as well as their shared regulatory mechanism.

### AVI feature attributions characterize functional impacts across the entire human genome, including cis-regulatory motifs

To explore how AVI can functionally inform rare non-coding variants genome-wide and to better understand its properties, we analysed the AVI scores and their feature attributions across the genome. Reassuringly, the higher the AVI scores (ranging from 0 to 55+), the more common variants were depleted and rare variants enriched, as expected based on the model’s training on population data (Fig. 5A). Moreover, higher scoring bins had higher rates of coding variants over non-coding variants, consistent with strong effects being more likely observed in protein-coding regions (Fig. 5A).

The feature attributions of the AVI scores revealed more nuanced discrete phases, suggestive of distinct functional features distributed along the AVI score ranges (Fig. 5B, fig. S11A). At the extreme high end (> 45), AVI scores are strongly driven by a combination of protein impact (protein termination or AlphaMissense), splicing, and conservation (Cactus-241 alignment and PhastCons). This trend, dominated by splicing and coding features, continues into the ranges below (AVI 22.5-45), with a distinct peak at AVI 32.5–35.0 where we observe the highest proportion of core splice sites (Fig. 5C) and canonical splice junctions based on VEP consequences (Fig. 5D). Thus, high AVI scores highlight AVI’s capacity to capture pleiotropic mutations that directly disrupt the production of functional proteins, e.g. prevent appropriate transcript processing or cause premature translation termination.

Advancing through the lower scores (AVI 5–22.5), we started to observe feature attributions characteristic of *cis*-regulatory regions. The attributions now increasingly highlight experimental modalities that capture regulatory regions, such as chromatin accessibility (DNase and ATAC), transcription factor binding (ChIP-TF) and histone modifications (ChIP-Histone), while still showing contribution from conservation features (Fig. 5B). More frequent are variants in non-splicing motifs (Fig. 5C) and in synonymous codons, 5’ and 3’ UTR regions and splice regions, which can be regulatory (Fig. 5D). This suggests that variants within the low-to mid-AVI regime capture functional *cis*-regulatory elements.

This trend is consistent with variant enrichments from TraitGym complex traits, TraitGym Mendelian disorders and ClinVar (Fig. 5E-F). ClinVar pathogenic variants are predominantly found in high AVI ranges where proteins tend to be impacted, while causal TraitGym Mendelian SNVs tend to be in lower AVI ranges, thus are more likely to affect regulatory regions. Causal variants of complex traits have even smaller AVI scores, including those in the lowest bound (AVI 0–5), which occasionally shows negative contributions from the Cactus-241 alignment and thus could belong to fast-evolving non-coding regions (fig. S11A).

Next, we analyzed AVI scores at the level of individual gene loci and motifs. At the locus level, AVI patterns reflect the functional composition of genes and their unique vulnerabilities to splicing effects, protein termination, missense and regulatory feature-driven variants, as exemplified at the *ACTA1* and *FOXL2* gene (fig. S11B). Near exon-intron boundaries, we observe a base-pair level trend in feature attributions, with protein termination and AlphaMissense signals accompanying an increasing splicing signal until reaching a peak at the canonical splice positions, then falling off into the intronic region (Fig. 5G). At the motif level, we found that splicing motifs have particularly high AVI scores (Fig. 5H) but other motifs also stand out, and their AVI pattern typically mirrors the base-specific contributions of the motif (Fig. 5H; fig. S12). Some motifs exhibited enriched feature attributions for specific modalities, including CTCF motifs for Contact maps as one would expect, but also included unexplained associations, such as the AP1-ETS motif for modalities that measure transcription initiation (Pro-CAP and CAGE) (fig. S13).

AVI’s ability to prioritize and interpret coding and non-coding variants at allelic resolution make it a powerful starting point for large-scale variant explorations. As a demonstration, we applied it to a recently released dataset of >70,000 somatic variants in oral epithelial cells (*54*). Among somatic coding variants, both loss-of-function and gain-of-function variants were systematically enriched in high AVI bins (fig. S14A). Conversely, non-coding variants exhibited a long tail of low-AVI intronic variants as well as high-AVI variants in 5’/3’ UTR and upstream gene regions (fig. S14B). For example, high AVI variants in the *TP53* 3’UTR and the *AJUBA* promoter overlapped with clusters of observed somatic mutations (fig. S14C). Notably, both of these non-coding gene regions were scored as under significant positive selection in a region-based test (*54*), suggesting that further investigation of these variants is warranted, similar to the burden testing application above.

In summary, these analyses across AVI bins suggest that causal non-coding and regulatory-motif-spanning variants appear in lower AVI bins than, for example, pathogenic protein-altering and splicing-motif-spanning variants. Thus, AVI is applicable to both coding and non-coding variant classes, but we recommend that users adopt genomic region- or application-aware thresholds when using AVI scores for variant selection, and prefer ranking metrics over hard cutoffs where possible.

### AlphaGenome Atlas captures cell-type-specific regulatory elements and reveals the role of TFs in transcriptional processes

Beyond prioritizing individual genetic variants, the AlphaGenome Atlas provides a powerful lens for interpreting *cis*-regulatory elements and the broader principles governing gene expression. Rather than being static genome-wide sequence matches, deep learning model-derived motif maps like Atlas are specific to both cell type and regulatory modality (*55*). This enables viewing regulatory *cis*-elements in a cell-type-specific manner and characterizing the effects of motifs on the gene regulatory processes captured by the Atlas modalities.

A good illustration of how to interpret *cis*-regulatory elements through Atlas is the *IRF6* locus (Fig. 6A). A well-studied enhancer 10 kb upstream of the start site mediates expression in the developing limb and orofacial ectoderm (*56–58*). The enhancer contains a TP63 motif that is mutated in a rare *cis*-regulatory variant causing cleft lip and palate (*56*, *58*), while the *IRF6* promoter contains EBOX motifs that are bound by the ZEB repressor (*59*) (Fig. 6A left). AlphaGenome Atlas indeed shows that the promoter has contributing EBOX motifs in epidermal cells (foreskin keratinocytes), while the same motifs are highlighted as negative ZEB motifs in mesoderm cells (Fig. 6A middle). Likewise, the two TP63 motifs (labeled as TP53 family) are highlighted surrounding a SP/KLF motif in the epidermis, but not in mesoderm (Fig. 6A right). Thus, AlphaGenome Atlas can be used to examine *cis*-regulatory elements for given cell types and identify potentially new candidate motifs of interest.

Atlas can also be used to analyze how motifs are predicted to contribute to the various gene regulatory processes leading up to the transcription of a gene, as measured by RNA-seq. Such regulatory processes include making chromatin accessible, measured by e.g. DNase-seq, and histone acetylation, measured by e.g. H3K27ac ChIP-seq (Fig. 6B, examples in Fig. 6A). Systematically linking motifs to the way they contributed to distinct modalities has the potential to reveal the regulatory roles played by the corresponding TFs, a concept we tested in the well-studied HepG2 liver cell line. Here, the motifs mapped as contributing to RNA-seq covered most motifs independently discovered by large-scale gene reporter assays (*60*) (bold motifs in Fig. 6B, fig. S15A). We then recorded the modality combinations that the motif instances positively or negatively contributed to, with each genomic motif instance assigned to a single category (Fig. 6B, see fig. S15B for rare categories).

Most motifs had instances in multiple modalities with distinct contributions towards chromatin opening and transcription, but the effects of a motif were often in only one direction, allowing us to classify motifs as belonging to activating or repressing TFs. Notably, motifs of well-known liver-specific transcriptional activators such as HNF4, HNF1, CEBP, and ONECUT (bound by HNF6) showed positive contributions towards all three modalities, including accessible chromatin, which has mechanistic implications (*61*, *62*). Other motifs contributed more toward H3K27ac (e.g. ETS-SP/KLF and ZNF76) or just RNA-seq (e.g. TATA) (Fig. 6B). Among the repressing motifs were GLI, ZEB or ZNF121 bound by zinc finger TFs. Some motifs, including CTCF and REST, had dual roles towards activation and repression, suggesting context-specific functions of these motifs. Overall, this classification is in agreement with the known roles of these TFs, while also suggesting avenues for future investigations.

We then computed how much each motif contributed on average to each modality across various cell types chosen from different lineages (Fig. 6C, Data S3). This revealed that some motifs like TP53, HNF4 and HNF1 contributed more strongly in a cell-type-specific manner, likely dependent on TF expression, while other motifs showed more consistent trends. Among them, motifs of signal-responsive TFs like AP1, ATF, ETS and TEAD tended to consistently contribute to all three modalities (Data S3, Fig. 6C), as supported by some experimental evidence (*63–65*). Dual regulators like CTCF consistently played both positive and negative roles. Some composite motifs like OCT-SOX (*66*) and AP1-IRF (*67*) were cell type-specific, while double motifs like ETS-ETS, SOX-SOX, GRHL-GRHL and TEAD-TEAD were more common (Fig. 6C). Composite motifs can be overlooked in genomic regions because individual motif instances are often degenerate versions of the motif, but they are readily discovered through deep learning models (*68*, *69*) and are well represented in Atlas.

Finally, we leveraged AlphaGenome’s breadth of predicted modalities to more broadly classify the function of repressors (Fig. 6D). Notably, repressors could promote repressive histone modifications, such as the H3K27me3 (Polycomb repression) or H3K9me3, and may not directly repress gene expression. Looking for consistent trends, we classified motifs as belonging to general repressors, histone repressors and chromatin-opening repressors. General repressors, like ZEB (*70*), had repressive effects across all modalities and sometimes had additional contributions towards histone modifications. Motifs of histone repressors were predicted to exclusively promote repressive histone modifications, especially H3K9me3. These included the motif of the zinc finger repressor ZNF274 (*71*), as well as unknown motifs (e.g. cluster #868 in Fig. 6D). Finally, motifs for REST, RFX4 and THAP11 fell into a repressor category that positively contributed to accessibility but negatively towards gene expression. We conclude that Atlas not only highlights relevant motifs across loci but can be used to more generally explore the regulatory role of motifs across cell types.

## Discussion

Understanding the biological mechanisms linking genetic variants to phenotypes remains the primary challenge in human genetics (*1*). Machine learning has emerged as a defining paradigm to tackle this challenge by distilling functional genomics and population-scale data into genome-wide, allelic-resolution maps of variant function. In this regard, AlphaGenome Atlas represents a milestone through its unprecedented scope by predicting regulatory variant effects for all possible human (hg38) single nucleotide variants and observed biobank indels across 11 modalities and hundreds of cell types. We showed how other resources can be built on top of these precomputed scores, exemplified by AVI and the motif maps across cell types and modalities, providing mechanistic insights into the genetics of both rare diseases and complex traits. Both of these resources are interlinked because they were derived from the same set of variant effects, meaning that high AVI scoring variants can directly be linked to motifs, cell types and regulatory roles.

Despite these advances, our models remain constrained by several limitations. First, AlphaGenome has gaps in its training data: standard poly(A)-selected RNA-seq excludes non-polyadenylated non-coding RNAs (exemplified by *RNU4-2* in Fig. 2D), many relevant cell types remain absent, and the distribution of cell types profiled across different assay modalities is uneven. Furthermore, because AlphaGenome is trained to capture *cis*-regulatory grammar, it does not directly model *trans*-acting mechanisms such as changes in TF expression, which mediate a substantial proportion of complex trait heritability (*72*). For AVI, the inclusion of evolutionary conservation features boosted performance, but may have obscured the molecular effect interpretation for variants where conservation is the predominant feature. Future models could address some of these limitations.

Caution should also be exercised around the practical use of variant impact models in real-world settings. Atlas and AVI are research tools that predict molecular effects, and therefore can only act as part of the evidence chain leading to clinical diagnoses, and are not sufficient evidence on their own. We hope that community-driven efforts will continue to make *in silico* models like AVI more useful by establishing robust standards for calibration of non-coding variants (*73*) and evaluating these tools across diverse genetic ancestries. In our association testing application, the coverage of traits remains limited, and ultra-rare variants intrinsically suffer from poor replication. Future directions would include exploring deep learning-enhanced rare-variant association testing methods such as DeepRVAT (*74*) and molecular characterizations of significant variants.

Likewise, the motif resource and its interpretation have limitations and opportunities for future improvement. The motif maps are inherently bounded by the fidelity of model predictions and may not always reflect direct causal interactions *in vivo*. Methodologically, the algorithms and parameter choices for motif discovery, instance calling, and cluster curation could be further advanced. Unlike axiomatic attribution frameworks such as Shapley values (*75*, *76*), ISM computes local counterfactual perturbations rather than an additive decomposition of total model output. Additionally, our curated motif labels are broad and agnostic to both the cell type and to cell-type-specific TF expression. Future work could more deeply harmonize our annotations with curated resources such as JASPAR (*77*) and ENCODE GRAMMAR (*16*, *19*) to eventually link discovered motifs to TF homolog(s) that mediate the predicted effects. Moreover, motif annotations for splicing and other RNA features remain to be improved. Finally, we have only begun to explore the underlying logic of the *cis*-regulatory motif maps and the biology they represent, e.g. the combinatorial rules of motif effects, the drivers of cell type specificity and the nature by which complex gene loci are predicted to be regulated have yet to be investigated. Despite these limitations, to our knowledge, the Atlas provides the most comprehensive sequence-level interpretation to date spanning modalities from gene expression, histone modifications, to splicing and chromatin accessibility, establishing an expansive foundation for future functional exploration.

In summary, despite these limitations, the Atlas resource can drive biological impact in the near term by augmenting statistical genetics workflows for interpreting the billions of uncharacterized germline variants, to resolve variants of unknown significance, and identify novel disease-gene associations. In regulatory genomics, Atlas could generate hypotheses about previously overlooked motifs and *cis*-regulatory elements and guide the design of high-throughput molecular biological screens to validate these elements. Longer term, this first-generation Atlas represents a baseline rather than an endpoint, and as sequence-to-function models mature, they will yield increasingly comprehensive and precise maps of the regulatory genome.

## Materials and methods

### Genome-wide inference of AlphaGenome

To compute the atlas database, several optimizations were applied to scale *in silico* mutagenesis (ISM) to the full human genome using the AlphaGenome distilled model (*9*). All variant scores were computed on the GRCh38 (hg38) reference genome assembly using GENCODE v46 gene annotations. The variant scoring methodology and the default set of variant scorers are the same as those described in Avsec et al. (*9*) and accessible via the AlphaGenome API (Table S1). We note that the ‘active allele scores’ were only calculated for variant scorers that perform aggregation across the positional axis first (for example calculating the total number of reads around the variant) before taking the difference between the reference and alternative allele. These were ATAC, DNase, ChIP-TF, ChIP-Histone, CAGE, PROCAP, and RNA-seq. Active allele scores are defined as taking the max operation instead of difference between the spatially aggregated predictions for the reference versus alternative allele. Active allele was not calculated for scorers for which the difference between reference and alternative allele was calculated first at each spatial position, followed by the average or max operation across spatial positions. These scorers were Polyadenylation, Splice Sites, Splice Site Usage, Splice Junctions, and Contact Maps.

### Sliding Window Approach

We applied a sliding window approach with a fixed step size of *W* = 128 bp to anchor the scoring intervals. The genome is tiled into non-overlapping 128 bp windows starting from position 0 (0-based). For each 128 bp window, the reference sequence is extracted and all possible single nucleotide substitutions are generated: at each position where the reference base is not N, the three alternative alleles (i.e., the three bases from {A,C,G,T} that differ from the reference) are enumerated, yielding up to 384 variants per window. The interval is then expanded to 1 Mb from the window’s interval center, compared to centering the interval on each variant as was done in previously reported evaluations (*9*).

### Reference Prediction Caching

Within a window, the reference forward pass is performed once and cached for all variants in the window. The 1 Mb input sequence is centered on the 128 bp scoring window, so variant positions deviate by at most ±64 bp from the exact interval center. We showed previously (*9*) that the model is insensitive to whether the variant is centered on the interval, thus allowing this kind of reference caching. We further verified that the small shift within a window for a variant position within the input interval does not meaningfully affect model predictions for the small shifts in the scoring window. Specifically, we re-ran the AlphaGenome paper evaluations with variants placed at their windowed positions rather than centered on the interval, and observed no statistically significant differences in any evaluation metric. This invariance to variant positioning near the interval center enables the reference caching strategy described above.

### Variant Mask Reuse

All variants within a 128 bp window share the same 1 Mb context interval. As a result, genomic annotation masks used in the scorers (such as exon boundaries, polyadenylation sites, and splice junctions) are computed once and reused for all 384 variants to amortize mask construction cost within the window. For center mask scorers and contact maps, the masks are computed for each variant.

### Indel Scoring

Insertion and deletion variants (indels) are scored separately from SNPs. The set of indels was determined as the union of variants observed in gnomAD v4.1 (*78*), UK Biobank (*79*), All of Us (*80*) and indels from all evaluation datasets used in the study. All indel variant lists are left-normalized to GRCh38 coordinates. No reference caching was performed.

### Indel shift augmentation

Deep learning architectures like AlphaGenome use convolutional encoder and decoder layers with pooling or strided downsampling. In these models, naive indel scoring, which uses a single forward pass for reference and alternate alleles, creates an artificial coordinate frameshift between the reference and alternate sequences downstream of the variant (*11*). This shift forces homologous downstream sequences into different pooling receptive fields. Consequently, the unaligned representations suffer from numerical artifacts and score distribution shifts that artificially inflate variant effect scores compared to SNVs.

To eliminate these artifacts, we perform intermediate activation stitching on the base-resolution representation of the model before evaluating the prediction heads (illustrated in fig. S1). This requires performing three forward passes instead of two for SNVs. In the first pass, all bases prior to the variant are aligned as normal, and in the second pass, the sequence is shifted to align all bases after the variant to the reference coordinates. We then stitch the output embeddings after shifting back the second pass, and feed them to the heads to compute the predictions. Similar to (*11*), a deletion is treated as an insertion on the reference sequence. The third pass is the unstitched reference or alternate allele, depending on the indel type.

### Base Resolution

Base-resolution and splice junction heads evaluate using the stitched trunk embeddings directly. Shifting and stitching operate at single-nucleotide resolution (fig. S1A), therefore, the augmentation happens seamlessly.

### Coarse Resolution

Coarse-resolution heads operate on bins representing multiple base pairs, making head stitching less precise than base-resolution operations. We deviate from the shift augmentation method of (*11*) for coarse-resolution representations (128 bp bins for 1D tracks and 2,048 bp bins for pairwise 2D contact maps). For indels smaller than the coarse resolution bin size, we match the method of (*11*) where representations remain consistent within the containing bin. When an indel is longer than the coarse resolution bin size, we shift downstream representations starting from the final bin where the indel terminates so that splitting occurs cleanly at that last bin boundary (fig. S1B).

### Indexing precomputed scores

Each variant score record is indexed by chromosome, position, and alternate allele (i.e., chr:pos: ref>alt). The reference allele is implicitly defined by the GRCh38 assembly and is not part of the lookup key. Queries must specify the alternate allele relative to GRCh38. Swapping reference and alternate alleles (e.g., based on minor allele frequency) will result in a lookup miss. Reference bases with N or n were not scored. No non-reference-to-non-reference substitutions (e.g., if the reference base at position X is A, the variant C>T at that position) were computed.

### AlphaGenome Variant Impact Scores

#### Training Data

The AlphaGenome variant impact model is trained on genomic variants derived from the Genome Aggregation Database (gnomAD) v4.1 genome data (gs://gcp-public-data-{}-gnomad/release/4.1/vcf/genomes/). For each chromosome, variants are extracted and classified based on their filtering allele frequency group maximum (FAF95_GRPMAX; specifically, the 95% lower bound of the maximum filtering allele frequency across genetic ancestry groups):

- **Proxy neutral (negative, label 0):** Variants with FAF95_GRPMAX *≥* 0.001 and < 0.999.
- **Proxy impactful (positive, label 1):** Variants with FAF95_GRPMAX < 0.001.

These variants constitute the pool of variants from which we sample actual training variants described in the following sections. From the pool of variants we sample 10 training datasets, each training dataset has the proxy impactful variants resampled to match the number of proxy neutral variants. The sampling strategy is described below.

#### Balanced Sampling

To mitigate sequence-context confounding biases, we perform stratified, balanced sampling independently for single-nucleotide variants (SNVs) and insertions/deletions (indels):

- **SNVs** are stratified by their trinucleotide context. Within each trinucleotide stratum, positive and negative variants are downsampled to match the size of the minority class.
- **Indels** (restricted to |len(REF) - len(ALT)| *≤* 10 bp) are stratified by the lengths of indels so that each training dataset has an equal number of positive and negative variants per indel length.

The balanced SNV and indel subsets are subsequently concatenated to form the training dataset for each chromosome.

#### Trinucleotide context

A mutational trinucleotide context is defined by concatenating the 5’ flanking base, the reference allele, the 3’ flanking base, and the substituted alternative allele (formatted as [5’-flank][3’-flank][ALT]). Considering all 64 possible trinucleotide sequences and 3 possible single-base substitutions per reference allele yields 192 potential mutational contexts (4^3^ *×* 3 = 192). To eliminate strand bias and account for double-stranded DNA symmetry, reverse-complement mutational contexts were treated as equivalent classes (for example, mapping 5’-ACG-3’ with alt T to its opposite-strand equivalent 5’-CGT-3’ with alt A), thereby collapsing the 192 contexts into 96 unique, strand-invariant trinucleotide contexts. Each SNV has a defined trinucleotide context and the context is used to sample matched proxy impactful variants.This context-matching strategy controls for local sequence-dependent mutational biases (such as CpG hypermutability) and ensures that baseline mutation frequencies do not confound downstream modeling.

#### Resampling Strategy

To account for sampling variability, we generate 10 resampled datasets using random seeds 0 through 9. Each resampled dataset is produced by running the balanced sampling procedure described above independently with a different seed, resulting in a different random subset of variants in each trinucleotide/indel-length stratum.

#### Data Split

To prevent data leakage and evaluate generalisation, variants are split into training, validation, and test as follows:

- **Training:** proxy neutral and proxy functional variants from chromosomes 1, 4, 7, 8, 10, 13, 15.
- **Validation:** variants on chromosomes 2, 5, 11, 14, 17, 20, 22, X from saturation genome editing, and TraitGym Complex traits datasets.
- **Test:** benchmark variants from genomic positions not used in the training or validation set. More details in *Overlap Filtering during Evaluation* section.

#### Overlap Filtering during Evaluation

To compare with other methods in downstream evaluation (e.g., on ClinVar, saturation genome editing, and TraitGym), we apply a coordinate-based filter: any benchmark variant is excluded from evaluation if its genomic position overlaps with any variant present in the training set or the validation set, regardless of whether they share the same reference or alternative alleles. For indel variants, the exclusion zone is expanded to encompass all genomic bases spanned by the indel, and any evaluation variant overlapping this spanned interval is discarded. To ensure equivalent variant sets, we applied this filtering of AVI training and evaluation variants across all methods (training/evaluation variants for other methods were not removed). Because the same training-set exclusion was not performed for external comparator models, any baseline overlap with benchmark sets presents a conservative bias against AVI.

#### Input Features

The model receives a concatenated feature vector comprising the following feature groups, with a total of 18 input features:

1. **AlphaGenome Scores (10 features)** This group extracts maximum genomic effect scores predicted by the AlphaGenome model across tissues, cell-type ontologies, genes, and junctions. Active allele scorers are excluded. Splicing scores are combined into a single feature computed as used in the AlphaGenome paper and are referred to as AlphaGenome merged splicing scores:

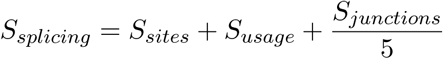

where each component is the maximum score observed across all coordinates and genes. The remaining 9 scorers (ATAC, contact maps, DNase, ChIP TF, ChIP histone, CAGE, PRO-Cap, RNA-seq, and polyadenylation) each contribute one feature computed as the maximum absolute value across all relevant dimensions of the scorers (i.e. tracks and genes). Indel variants are scored using the AlphaGenome model in the same manner as SNVs.
2. **AlphaMissense Score (1 feature)** Pre-computed scores for missense variants from the AlphaMissense model (https://zenodo.org/records/10813168). Because AlphaMissense exclusively scores single-amino-acid substitutions resulting from missense SNVs, indels do not have precomputed scores and are trained by imputing 0. During inference, we instead impute the mean AlphaMissense score for SNVs in the interval spanning the deleted bases for indel variants (or for insertions, the interval spanning the positions where bases are inserted). We primarily look for predictions with the canonical transcript isoform (AlphaMissense_hg38.tsv.gz), then isoform specific predictions (AlphaMissense_isoforms_hg38.tsv.gz). Variants without AlphaMissense predictions are imputed with 0.
3. **Zoonomia Cactus Conservation score (1 feature)** The zoonomia 241-way Cactus multi-species alignment phyloP score at the variant position, extracted from a BigWig file (https://hgdownload.soe.ucsc.edu/goldenpath/hg38/cactus241way/). The scores range from −20 to 8.9, with higher scores meaning more conserved. For deletions, the conservation score is extracted across the entire deleted interval, and the maximum value observed across those coordinates is returned. For insertions, the conservation score is extracted at the variant’s start position (corresponding to the anchor base), identical to SNVs.
4. **PhastCons 470-way Conservation (1 feature)** PhastCons conservation probability based on a 470-way vertebrate alignment, extracted from a BigWig file (https://hgdownload.soe.ucsc.edu/goldenpath/hg38/phastCons470way/). For deletions, the conservation probability is extracted across the entire deleted interval, and the maximum value observed across those coordinates is returned. For insertions, the conservation probability is extracted at the variant’s start position (corresponding to the anchor base), identical to SNVs.
5. **Consequence (LOF) (3 features)** Binary indicators derived from Variant Effect Predictor (VEP) Sequence Ontology (SO) annotations and ClinVar descriptors for loss-of-function (LOF) consequences, obtained for both SNVs and indels:

- **Protein termination:** maps from stop_gained, or frameshift_variant, or ClinVar nonsense.
- **Stop lost:** maps from stop_lost.
- **Start lost:** maps from start_lost or ClinVar initiator_codon_variant. If a variant is annotated with multiple distinct fine-grained consequences within the same coarse category, the binary indicator is combined using a logical OR operator (set to 1.0). VEP annotations were generated using the Ensembl VEP tool (ensembl.org/ensembl-vep:release_115.0 with --distance 200 --pick arguments set).
6. Indel Type (2 features) Binary indicator variables for insertion and deletion.

| Feature Group | Category | Number of Features | Raw Feature range | Normalization | Description / Source |
| --- | --- | --- | --- | --- | --- |
| AlphaGenome Scores | Sequence-to-function | 10 | [0, +inf) | MaxAbsScaler | Aggregated variant effect scores across 9 scorers and one merged splicing score |
| AlphaMissense | Coding | 1 | [0, 1] | None | AlphaMissense pathogenicity score |
| Cactus | Conservation | 1 | [-20, 8.9] | MaxAbsScaler | 241-way Cactus multi-species alignment |
| PhastCons 470-way | Conservation | 1 | [0, 1] | None | 470-way vertebrate conservation probability |
| Consequence (LOF) | Coding | 3 | {0, 1} | None | Loss-of-function consequence indicators |
| Indel Type | Indel Type | 2 | {0, 1} | None | Insertion/deletion indicators |

### Feature Summary and Normalization

The MaxAbsScaler divides each feature by its maximum absolute value observed in the training set:

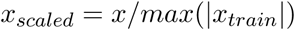

All other features are used without additional transformation.

#### Model Architecture

The model architecture consisted of three components summed together to produce the output logit: (1) a linear model, (2) a linear hypernetwork (*81*), and (3) an indel embedding used as an indel-specific offset (fig. S2A). The exact hyperparameters and architecture varied by model used within the ensemble (see Ensemble Models section below). The inputs to the model were the primary features (x) of dimension 16 and the indel type indicator (v) of dimension 2. The linear model operated solely on the primary features x and consisted of trainable weight and bias terms. The linear hypernetwork employed an encoder, a two-layer neural network with multiple output heads, that generated the weights and bias of a target linear model. Unlike the standard hypernetwork setting, the same input feature vector x served both as input for weight generation and as the operand of the target linear model, with indel type v used exclusively as an input to the encoder. Lastly, an indel-type offset was learned by embedding v to enable the model to account for baseline numerical differences between SNVs and indels.

The encoder produced the weights and bias of the target linear model by first embedding **x** and **v** into a hidden state, **h** = **h***_φ_*(**x**, **v**) (fig. S2B). It then used separate linear projection heads to generate the weights **w***_φ_* and bias *b_φ_* from **h**. Specifically, **w***_φ_* used a linear layer followed by an activation function (exponential or softplus) to enforce non-negativity. This was found to improve both training stability and performance. The bias term *b_φ_* was computed from two linear projections of **h**. The first, *g*(**h**), was a linear projection with bias term and sigmoid activation, serving as a gate. The second, *b^′^*(**h**), was a linear projection with no activation function or bias term. These were combined as either an additive or standard gated bias:

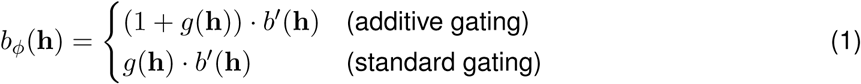

This choice differed across the ensembled models. The encoder, h*_φ_*, consisted of two fully connected layers operating as a function of both x and v (fig. S2C). Each layer was regularized using L2 activity regularization (*λ* = 10*^−^*^5^) and dropout (rate = 0.4), followed by a Gaussian Error Linear Unit (GELU) (*82*) activation function. Weights were initialised using He normal initialisation.

The model’s output logit for an individual variant was computed as:

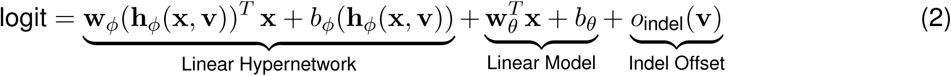

where:

- x *∈* ℝ^16^ denotes the primary feature vector (the 16 features excluding indel type features)
- v *∈* {0, 1}^2^ denotes the input vector indicating indel type (v = [1, 0] for insertion, v = [0, 1] for deletion, and v = [0, 0] for SNVs)
- h*_φ_*(x, v) *∈* ℝ*^dℎ^*, *d*_ℎ_*∈ {*16, 32*}*, denotes the hidden state output from the encoder network
- 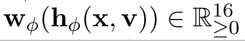 denotes the vector of non-negative weights generated by the hypernetwork
- *b_φ_*(h*_φ_*(x, v)) *∈* ℝ denotes the scalar bias generated by the hypernetwork
- w*_θ_ ∈* ℝ^16^ and *b_θ_ ∈* ℝ denote the trainable weights and bias of the linear model component
- 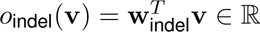 denotes the learned offset, w_indel_, for the variant’s indel type (equal to 0 for SNVs)

During training, a sigmoid activation was applied to the logit within the binary cross-entropy loss, whereas during inference the raw logit was used directly for predictions and the computation of PHRED-scaled scores. The following model hyperparameters were used:

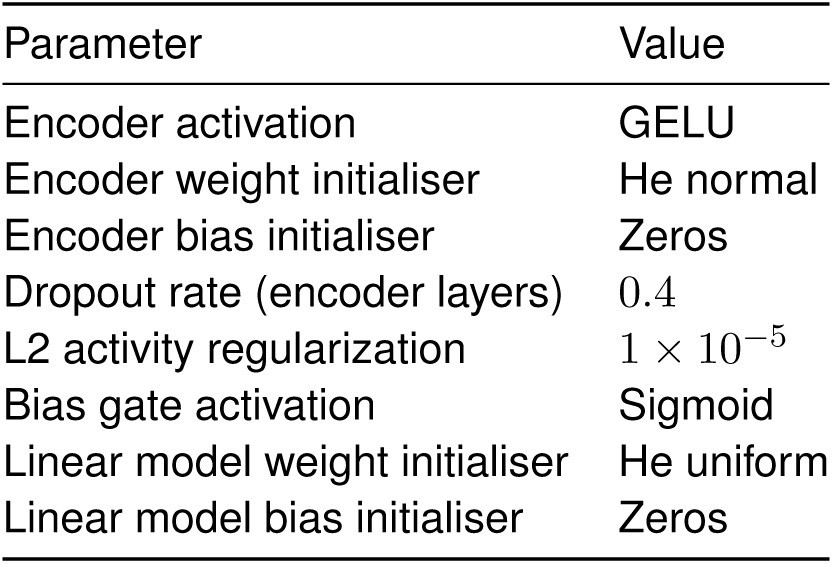

#### Training Procedure

Each ensemble member was optimised independently to minimise binary cross-entropy loss using the AdamW optimiser. Training hyperparameters were standardised as follows:

| Hyperparameter | Value |
| --- | --- |
| Batch Size | 500k |
| Training set size | 37M |
| Learning Rate ($\eta_0$) | $1 \times 10^{-4}$ |
| Learning Rate Schedule | Linear warmup (75 steps) followed by continuous exponential decay |
| Decay Rate | 0.99 per 300 steps |
| Weight Decay | 0.01 |
| Global Gradient Clipping | 0.5 |
| Maximum Epochs | 1500 |
| Steps Per Epoch | 10 |
| Early Stopping Patience | 150 epochs (monitoring validation metric) |

The learning rate followed a two-phase schedule. During the first 75 optimiser steps, the learning rate increased linearly from 0 to *η*_0_, after which it decayed exponentially at a rate of 0.99 per 300 steps.

Model checkpoint selection was determined using the average AUPRC across the validation set of TraitGym Complex Traits, SGE SNVs, and SGE indels. Training was terminated early if the overall validation average AUPRC failed to improve for 150 epochs.

#### Ensemble Models

The final model was an ensemble of six models selected from a large hyperparameter sweep over 10 resampled datasets (seeds 0–9), with the output computed as the average of the six model logits. The sweep evaluated encoder hidden layer sizes of (16, 16) or (32, 32) units, exponential or softplus weight activation functions, and if additive bias gating was used. For each configuration and data resampling seed, models were trained using 50 random initialisation seeds (seeds 0–49). The six models with the highest validation AUPRC, each from a distinct training seed, were selected for the final ensemble. All other architecture and training hyperparameters were shared across ensemble models. The selected models differed in the following hyperparameters:

#### Feature Attribution Analysis

To quantify the contribution of individual features to variant pathogenicity predictions, we calculated feature attribution scores using the SHAP framework (*10*). This framework decomposes the model output logit for any individual genomic variant by allocating the difference between the prediction and its baseline expectation across the input features.

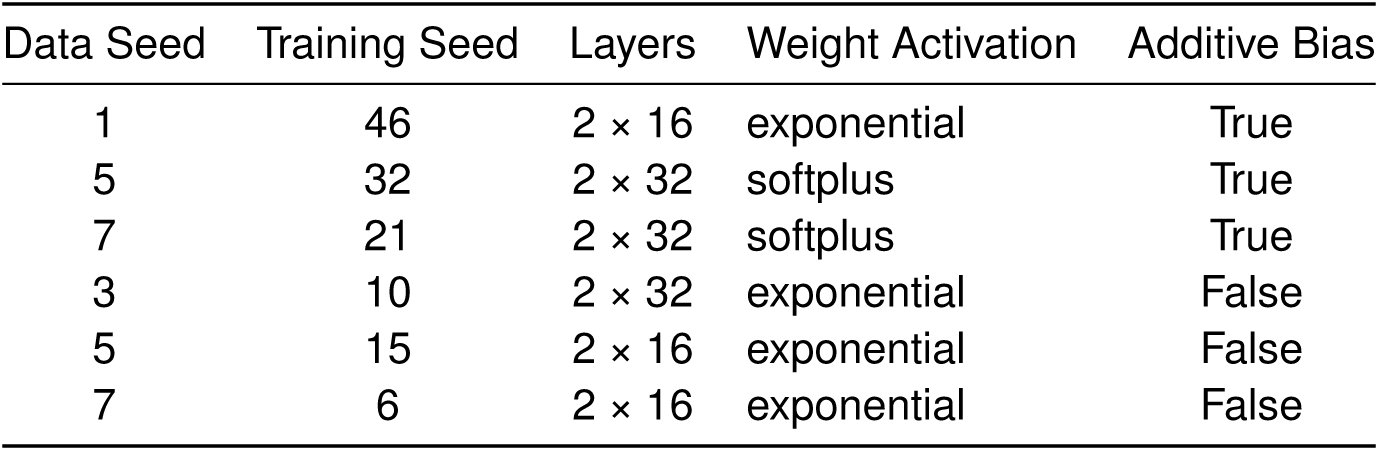

We used the expected gradients algorithm (using shap.GradientExplainer), which approximates Shapley values by integrating the gradients of the model’s output logit with respect to the input features along paths between the target variant feature vector and a background reference distribution:

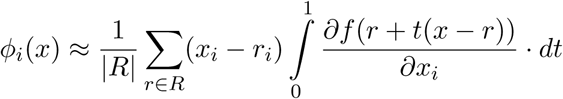

where *x ∈ R^d^* denotes the feature vector of the target variant, *R* represents the background reference set, and *f*(*x*) is the model output logit. The background reference set *R* = *{*0*}* consists of a single zero vector, representing the absence of all feature signals.

#### Obtaining baseline scores

##### CADD

We used CADD v1.7 (GRCh38) precomputed scores. For single-nucleotide variants (SNVs), scores were obtained from the publicly available genome-wide precomputed file distributed by the CADD authors (https://krishna.gs.washington.edu/download/CADD/v1.7/GRCh38/). For indels, we first looked up scores from the CADD-precomputed gnomAD v4.0 indel file. Benchmark indels absent from this file were scored by running the CADD v1.7 scoring pipeline via its Docker image (https://github.com/kircherlab/CADD-scripts) on VCFs containing the missing variants. The resulting scores were stored in a tabix-indexed file. At query time, the three sources (genome-wide SNVs, gnomAD indels, and benchmark-specific indels) are searched sequentially and the first exact match (on chromosome, position, reference allele, and alternate allele) is returned.

##### GPN-Star

We scored all possible SNVs in GRCh38 with the author provided code for GPN-Star (mammalian) and GPN-Star (vertebrate) (https://github.com/songlab-cal/gpn) (*5*). Scores are calibrated with mutation rate as described by the authors. Indels cannot be scored by these methods. Therefore GPN models are only included for benchmarks with SNVs only.

##### AlphaMissense

We used the publicly available AlphaMissense (hg38) precomputed scores, supplemented by the isoform-level score file for variants not covered by the canonical transcript file. AlphaMissense only scores missense (protein-altering) single-nucleotide variants. All other variant types — including synonymous, intronic, UTR, and all indels — are assigned missing values.

##### Conservation scores

We evaluated four evolutionary conservation baselines which we used as features: phastCons 470-way, and Cactus 241-way (phyloP method, noted as Cactus). Scores were extracted from publicly available BigWig files described above. Because the BigWig format stores a single conservation value per genomic position, these scores are position-level and do not distinguish between alternate alleles at the same locus. Insertions are scored as the conservation score at the basepair where insertion happens. For instance, the conservation score for variant chr1:1000:G>GA is scored with the conservation score at position 1000. Deletions are scored as the max conservation score across the deleted region.

##### gnomAD allele frequency

We used gnomAD v4.1 genome allele frequencies as a baseline (gs://gcp-public-data--gnomad/release/4.1/vcf/genomes/). Allele frequency was negated so that rarer variants receive higher scores (consistent with the expectation that pathogenic variants are depleted from the population). For variants not observed in gnomAD v4.1, allele frequencies are set to be 0.

##### Feature Ablations

To evaluate the contribution of individual features to the final performance of the AVI model, we performed two types of feature ablations: inference-time zeroing and model retraining. For inference-time ablations, features were set to zero during the evaluation of the frozen AVI model. For retrained ablation models, feature inputs were set to zero during training and evaluation. We aimed to keep model capacity and weight initialization constant across all ablated and non-ablated models. Each ablated model was an ensemble consisting of six retrained models using the same hyperparameters as the ensembled AVI model. Checkpoint selection was performed using the same method as for the AVI model.

##### AVI Evaluation Benchmarks

To comprehensively assess clinical and functional predictive performance, model predictions are systematically evaluated across a diverse suite of benchmarks. This evaluation matrix spans both coding and non-coding variations, capturing clinical phenotypes and quantitative functional disruptions across multiple molecular layers.

##### ClinVar Pathogenicity Classification

Model predictions were evaluated against clinical variant assertions archived in the ClinVar database (June 15, 2025 release). The evaluation dataset was filtered to include only variants with the following labels:

- Positive Class (Pathogenic): Variants with aggregate VCV-level classifications designated as “Pathogenic” or “Likely Pathogenic”.
- Negative Class (Benign): Variants with aggregate VCV-level classifications designated as “Benign” or “Likely Benign”.

Variants with aggregate VCV-level classifications characterized as Variants of Uncertain Significance (VUS) were excluded. Variants from the Y chromosome and the mitochondrial genome are removed. Indels length more than 10bp are not included. No filter based on review status (stars) was applied. To resolve performance across distinct molecular mechanisms, variants were annotated via the Ensembl Variant Effect Predictor (VEP) and stratified into structural and regulatory categories: missense (protein-altering), synonymous, intronic, 3’ untranslated region (3’ UTR), 5’ untranslated region (5’ UTR), and gene neighborhood classes. Classification accuracy was quantified using the Area Under the Precision-Recall Curve (AUPRC).

##### Saturation Genome Editing (SGE)

We obtained saturation genome editing data from a combination of source publication supplemental tables and the MaveDB database (*83*). The data sources were:

| Study | Gene/Target |
| --- | --- |
| Findlay et al. 2018 (84) | *BRCA1* |
| Huang et al. 2025 (85) | *BRCA2* |
| Waters et al. 2024 (86) | *BAP1* |
| Olvera-Leon et al. 2024 (87) | *RAD51C* |
| Buckley et al. 2024 (88) | *VHL* |
| Radford et al. 2023 (89) | *DDX3X* |
| Dejonghe et al. 2025 (90) | *RNU4-2* |
| Lee et al. 2025 (91) | *ATM* |
| Dace et al. 2025 (31) | *BRCA1* |
| Woo et al. 2026 (92) | *BARD1* |
| Tejura et al. 2026 (93) | *PALB2* |
| Tejura et al. 2026 (93) | *XRCC2* |
| Tejura et al. 2026 (93) | *SFPQ* |
| Tejura et al. 2026 (93) | *CTCF* |

For Dace et al. 2025 (*31*), we used the screen they performed in HAP1 cells.

The thresholds proposed by the authors within the original source tables were used to define functionally normal and functionally abnormal classes within each study. For example, for the RNU4-2 study, a cutoff of −0.3 was applied as suggested in the text of (*90*). In cases where authors would also designate an ‘uncertain’ or ‘intermediate’ category, those variants were filtered out.

The data split was as follows: Four studies were used as online evaluation (not training) of the AVI model during training: FINDLAY_2018_BRCA1, LEE_2025_ATM, OLVERA_LEON_2024_RAD51C, and RADFORD_2023_DDX3X. The remaining ten studies were held out as a test evaluation of the model. For these test evaluations, positional filtering was applied to filter out variants present in the AVI training data. For Spearman correlations, the AVI score was compared to the reported variant rankings within each study. For precision recall calculations, the functionally normal vs. abnormal class designations were used.

##### TraitGym Phenotypic Association

To establish the clinical and epidemiological relevance of the model’s predictions, we used the TraitGym benchmarking suite (*30*), which segregates human phenotypic associations into dual components:

- TraitGym Mendelian: Validated high-penetrance pathogenic variants underlying classical Mendelian disorders from OMIM matched 9:1 against gnomAD common variants.
- TraitGym Complex: Fine-mapped genome-wide association studies (GWAS) variants from UK BioBank (PIP > 0.9), matched with control variants that have PIP < 0.01 across all traits.

To neutralize confounding genomic architectures (e.g., local mutational rates and selective constraints), causal variants were matched with non-causal control variants through a multi-dimensional propensity score strategy based on three key parameters: distance to the nearest transcription start site (TSS), minor allele frequency (MAF), and local linkage disequilibrium (LD) scores. Evaluation performance was summarized via AUPRC.

##### Complex Trait Benchmark

The model’s resolution across varying phenotypic effect sizes was characterized using conditionally-independent variants associated with complex traits based on a meta-analysis of UK Biobank (*79*) and All of Us (*80*). To isolate performance as a function of genetic architecture, variants were partitioned into discrete strata based on their absolute linear regression effect sizes (*|β|* coefficients):

- High-Effect Architecture (High *β*): Causal variants with absolute effect sizes exceeding the 80th percentile of the trait-specific distribution.
- Low/Medium-Effect Architecture (Low/Medium *β*): Causal variants capturing subtler polygenic signals, falling below the 80th percentile.

Control variants were dynamically sampled to match the allele frequency spectra and sample sizes (cases) amongst the associated variants. Performance was evaluated using AUPRC.

##### ncVarDB Benchmark

Pathogenicity prediction within non-protein-coding domains was benchmarked using the curated ncVarDB (*94*). This non-coding metric consists of:

- Positive Class: 721 high-confidence non-coding variants with validated clinical pathology or experimental evidence of transcriptional/post-transcriptional disruption.
- Negative Class: 7,228 benign, common non-coding variants exhibiting high allele frequencies and no documented phenotypic associations.

#### Variant analyses and SGE examples

##### Locus view figures

Genome browser style plots were created by querying the AVI score, feature attributions, AVI input features, saturation genome editing data, and annotation tools such as VEP Consequences and ENCODE cCRE4 data. These data points were plotted with the x-axis indicating chromosome position. Two resolutions were used across the study: 1) basepair resolution, in which the max was taken across the three alternative alleles corresponding to that position, and 2) allelic resolution, in which each allele is plotted independently as separate columns.

##### AlphaGenome REF/ALT plots

For selected variants we also chose to show the basepair resolution predictions for the reference and alternative allele in track coverage format. This uses the standard open source visualization library that was published alongside AlphaGenome (https://www.alphagenomedocs.com/api/visualization.html). Tracks were selected for visualization based on the following various criteria: best biosample substitute (e.g. HAP1 was the cell line used in many SGE screens, but has limited track data in the AlphaGenome model, so K562 was used as a next-best substitute), or using the max(abs(track)) nominated by the AVI score.

#### GREGoR analysis

##### GREGoR retrospective

Individual cases annotated as solved from the Broad Institute Center for Mendelian Genomics subset of the GREGoR rare disease consortium were collected. These cases included 112 SNV or indel variants classified as pathogenic/likely pathogenic according to ACMG guidelines (*95*) with submissions to ClinVar. All small variants (<50 base pairs) called in these genomes from autosomal and X chromosomes were annotated with AVI and CADD v1.7. Within each solved case, the pathogenic or likely pathogenic allele was ranked by AVI and CADD against a background of all other variants within that individual. Both SNVs and Indels are considered, therefore models that can only score SNVs are not included. The distribution of ranks across different rank bins is reported in Fig. 3A. We also applied a gnomAD allele filter (AF) to model rankings when only considering variants with AF < 0.001. Cases for which the causal variant filtered out in this process were not considered. 105 variants were included and the results are reported in fig. S9A.

##### GREGoR case analyses

Bulk-called VCFs of genomic variants from the GREGoR consortium cohort were annotated with AVI scores. For probands with parent genomic data available, variants predicted to have arisen *de novo* were identified and ranked by AVI. *De novo* variants with AVI scores in the top 1% were subsequently analysed on a case-by-case basis for possible pathogenicity based on minor allele frequency in the gnomAD population database, gene constraint, and fit of variant predicted mechanism and gene function with the individual proband phenotypes.

Prior to this study, the RGP_2167_3 participant with epileptic encephalopathy underwent two independent rounds of blood-derived RNA-seq that yielded ambiguous results. The leading candidate after RNA-seq was a 5’UTR variant in *CSRNP1*, which was not a known haploinsufficient epilepsy disease gene, thereby falling short of ACMG/AMP criteria for Likely Pathogenic (LP).

#### Analysis of AVI feature attributions

##### Genome-wide AVI feature attribution analyses

In Fig. 5A-F, all human SNVs are considered. Allele frequency is the FAF95_GRPMAX (see Training Data) and bucketed into Missing/Zero, Rare (<1%), and Low Frequency or Common (>=1%).

The absolute feature attributions are the mean absolute values of the feature attributions across all variants in each PHRED bin, and the scaled feature attributions are the fraction of the total absolute value of feature attributions attributed to each feature in each PHRED bin.

The VEP consequences are the number of variants belonging to each consequence, normalized by the overall number of variants in that consequence genome-wide, then scaled to 100% in each PHRED bin.

If a variant belonged to more than one consequence, we first selected the highest severity consequence based on the following order: [CANONICAL_SPLICE, PROTEIN_TERMINATION, START_LOST, STOP_LOST] > [INFRAME_INDEL, PROTEIN_ALTERING] > [SPLICE_REGION, SYNONYMOUS] > All others. If a variant still belonged to more than one consequence, we distributed its weight equally among the highest-severity consequences.

In Fig. 5G, we binned all SNVs by their distance from the nearest exon-intron (splice donor) or intron-exon (splice acceptor) boundary as defined in GENCODE GTF V46. We then calculated the average feature attributions for all variants in each distance bin.

##### Buccal swab somatic mutation data

Somatic mutation data from Supplementary Table 8 of (*54*) was downloaded, which contained a table of all somatic mutations that were called in oral epithelium. Genomic coordinates were converted from human genome assembly GRCh37 (hg19) to GRCh38 utilizing the pyliftover library and the UCSC hg19ToHg38.over.chain.gz chain file. Lifted coordinates were cross-referenced against the GRCh38 reference genome using pysam. Successfully lifted coordinates were formatted into standard Mutation Annotation Format (MAF). Unmapped variants and indels were discarded, thus retaining only single nucleotide variants.

The July 2026 version of the OncoKB database API (*96*) was used to annotate each variant via the oncokb-annotator framework. The appended features that were used in downstream processing included:

- HUGO_SYMBOL: name of the gene associated with each variant
- MUTATION_EFFECT: We only used the Gain and Loss of function annotations (merging ‘Loss-of-function’ and ‘Likely Loss-of-function’, as well as ‘Gain-of-function’ and ‘Likely Gain-of-function’, into their respective single categories).
- CONSEQUENCE: Sequence ontology consequence category provided by OncoKB (*96*).

To derive an annotated set of variants used for plotting, we performed the following filters and variant annotations:

1. Supplementary Table 3 of (*54*) had gene-level selection calculations that report 49 positively selected genes as calculated by the dndscv package (https://github.com/im3sanger/dndscv). Using the HUGO_SYMBOL column, we filtered to those variants which were mapped to those genes.
2. Variants whose MUTATION_EFFECT was mapped as LOF or GOF by OncoKB retained those annotations.
3. Of the variants unannotated by OncoKB, we then labeled all variants called in Supplementary Table 4 of (*54*), which reported a list of variants with dndscv.sitednds dN/dS positive selection coefficients with q-value <0.01. These variants were labeled as ‘Somatic positive selection’
4. Of the remaining variants, we split into two categories:

1. Other, indicating other coding or splicing mutations that would have been considered by sitednds but whose q-values were not reported. This was done by considering the OncoKB CONSEQUENCE column and mapping to the three consequences required by sitednds (which are Synonymous, Missense, or Nonsense)
2. Non-coding, non-splicing: Remaining variants (by exclusion, these consequences included 3’UTR, 5’UTR, Downstream, Upstream and Intron).

We visualized the distribution of AVI values for all variants across each of the five major categories (Known LOF, Known GOF, Somatic Selection, Other, and Non-coding, non-splicing), using the same AVI bins as defined in Fig. 5A. For the non-coding specific breakdown, we faceted the Non-coding, non-splicing variants into their respective OncoKB CONSEQUENCE labels.

For choosing which loci to plot, we leveraged the withingenednds table provided by Andrew Lawson (personal correspondence) that accompanies the data outputs of the original study (*54*). From here, we selected gene regions that scored as under significant positive selection.

#### Motifs and motif instances

To identify and map the regulatory sequence motifs learned by the AlphaGenome model across diverse genomic modalities, we implemented a pipeline consisting of contribution score definition and rescaling, region selection, motif discovery, hit calling, compendium curation, mapping, and threshold filtering.

##### Generating the contribution scores from AlphaGenome *in silico* mutagenesis

The positional contribution score was defined as the negative average of all possible SNV scores at a given position (e.g., for reference A, the negative average of A>C, A>G, A>T). For strictly non-negative tracks (e.g., active allele, splicing, contact maps), we omitted the negation to maintain positive final scores, facilitating downstream sequence logo visualization.

##### Contribution Score Rescaling with Active Allele

ISM contribution scores are calculated independently per base, unlike popular additive attribution methods such as DeepLIFT or SHAP, which simultaneously allocate overall prediction differences from background to an entire prediction window of sequences. This means that additive attribution methods typically produce local contribution magnitudes that are inherently proportional to the overall predicted signal (e.g. DNase peak height or RNA-seq gene expression), whereas ISM contribution scores may not. To make the ISM contribution scores more comparable to these methods and thereby more familiar to their users, we developed an alternative, rescaled version of the contribution scores alongside the original unscaled version. This rescaled version exhibits the property of being proportionally scaled by the overall signal. In this rescaled version, the local contribution scores are multiplied by the “active allele” signal: Active Allele=max(Signal_REF, Signal_ALT).

This “times active allele” rescaling strategy was applied as the default for all scorers that support active allele calculations (including DNase, ATAC, ChIP-TF, ChIP-Histone, CAGE, PRO-cap, and RNA-seq). For specific scorer types where active alleles were not computed (including splicing junctions, splicing sites, splicing site usage, polyadenylation, and chromatin contact maps), no active allele rescaling was performed, and the raw contribution scores were retained without this adjustment. The rescaled ISM scores were pre-computed by element-wise multiplying contribution scores with the active allele scores at each base pair.

##### TF-MoDiSco: Discovering motifs from contribution scores

We performed motif discovery using TF-MoDISco-lite (v1.0.0; https://github.com/jmschrei/tfmodisco-lite) on the unscaled contribution scores independently across all tracks and scorers, excluding the active allele tracks.

##### Region selection

Previous motif discovery approaches typically rely on a predefined set of peaks, such as those from DNA accessibility assays, to select candidate regions. Because our goal was to systematically discover motifs across all 5,000+ AlphaGenome tracks, we developed a peak-independent strategy for region selection. To select a subset of the most informative regions for motif discovery, we tiled the genome into 1,000 bp intervals. For gene-specific or strand-specific scorers, intervals were considered separately for each associated gene or strand to ensure multi-gene regions were appropriately represented. To identify candidate motif sites (“seqlets”) within each interval, we applied the seqlet detection procedure from TF-MoDISco-lite: the contribution scores were smoothed using a 21 bp sliding window, and local peaks in the smoothed signal were identified by iterative greedy peak finding. At each iteration, the position with the highest absolute smoothed contribution score was recorded, and a suppression window was applied around the peak to prevent double-counting. This process was repeated to extract the top 5 seqlet scores per interval. For tracks generating signed contribution scores, the analysis was restricted to positive scores to focus on motifs associated with increased model predictions (such as active enhancers or promoters). To determine the regions to retain for TF-MoDISco-lite execution, we applied the Farthest Point Method (a global elbow thresholding algorithm) to the distribution of all extracted positive seqlet scores. A region was selected if at least one of its top 5 seqlet scores exceeded this dynamically determined elbow threshold, typically resulting in approximately 100,000 selected regions per track.

##### TF-MoDISco Execution

We ran TF-MoDISco-lite on the selected regions independently for each track using default parameters (sliding window of 20 bp, flank size of 5 bp, and target seqlet FDR of 0.05). To capture a larger and more comprehensive set of motifs, the maximum number of seqlets per metacluster was set to a non-default limit of 200,000. In total, 898,874 unique per-track motif patterns were discovered across all tracks. Each pattern or motif is represented in TF-MoDISco by a contribution weight matrix (CWM) representing the average contribution scores of seqlets and the position frequency matrix (PFM) representing the sequence base frequency of seqlets.

We note that the version of TF-MoDISco-lite used required both positive and negative contribution scores, which we did not have for unsigned variant scorers. To work around this technical code requirement, we artificially created a mirrored copy with an inverted sign and then only extracted the discovered motifs for the negative scores. Given a lower number of discovered motifs for unsigned scorers, it is likely that the TF-MoDISco-lite hyperparameters might be sub-optimal for these scores.

##### FiNeMo motif instance calling

Using the discovered per-track motif patterns, we performed genome-wide instance calling using FiNeMo (commit 5c6f521; https://github.com/austintwang/finemo_gpu) on the unscaled contribution scores. For each called instance, the location with the highest score was used to define its genomic position. For each track, we used the complete set of original TF-MoDISco motifs discovered for that track. By using the full original set of motifs, FiNeMo can account for both biological motifs as well as motifs corresponding to assay-specific biases, which may help absorb assay-specific bias contributions that would otherwise be incorrectly attributed to regulatory motifs.

To scan the genome, we divided it into windows of 1,050 bp, starting a new window every 1,000 bp to create a 50 bp overlap. This padding ensures that motifs falling near window boundaries are not missed during scanning. Within each window, FiNeMo modeled the local contribution scores using non-negative Lasso regression with a regularization penalty of *λ* = 0.7 to call motif instances.

For each called instance, the position of the highest score was used to define its genomic coordinate. Instances were deduplicated by assigning each hit to the non-overlapping 1,000 bp core interval containing its start position, ensuring each unique instance is reported exactly once.

For every called motif instance, FiNeMo output three primary metrics:

- Hit Coefficient: The scaling factor determined by the Lasso optimization, representing the fitted amplitude of the motif in explaining the local contribution signal.
- Hit Similarity: The cosine similarity between the local contribution scores and the reference motif CWM, reflecting how well the contribution score shape and sequence matches the motif profile.
- Hit Importance: The sum of absolute contribution scores across the base pairs of the trimmed motif window, representing the total contribution magnitude localized to the motif instance.

To account for regional variation in score scales (ensuring that hits in regions with naturally stronger ISM signals are prioritized), FiNeMo first normalizes the input contribution signal by its local root mean square (RMS) before fitting. The resulting Lasso coefficients are then multiplied back by this local RMS to yield the unscaled global hit importance.

For RNA-seq and other gene-specific tracks where a single genomic region can be associated with many genes, we sped up the runtime by restricting instance calling to the top three genes with the highest maximum absolute contribution score in the region, while requiring that at least two of these genes be protein-coding.

##### Motif clustering using MotifCompendium

To create a unified, non-redundant dictionary of motifs across all tracks, we used the MotifCompendium (https://github.com/kundajelab/MotifCompendium, 10.5281/zenodo.17123347) to cluster the motifs discovered by TF-MoDISco across all modalities and cell types.

Raw MoDISco patterns were aggregated by reading and merging the pattern files generated across all independent track runs. To manage the computational scale, patterns were initially processed in separate compendium objects for each model output type (corresponding to the multi-task ‘heads’ of the model, such as DNASE or RNA_SEQ. CHIP_HISTONE and CHIP_TF heads were further split into approximately similarly sized slices due to the number of tracks present in these modalities).

We applied a stringency filter requiring a minimum seqlet support of 100 seqlets per pattern to remove low-confidence motifs. Modisco motifs were then filtered using several metrics calculated with the MotifCompendium package and threshold values selected independently per each output type. Motifs failing any of these thresholds were only retained if they showed matches to known matrices in reference databases (including JASPAR2026, expanded with splicing and RNApolII motifs from 2018 and 2020 editions), defined by a similarity score of at least 0.85 (or 0.7 for secondary rescue of composite motifs).

The filter-passing patterns were then pre-clustered within each output type with a high similarity threshold of 0.97 to reduce redundancy and computational load. Aggregated patterns were then clustered using a two-phase approach:

1. First, it performed a global round of community detection using Leiden clustering with the Constant Potts Model (cpm) at a similarity threshold of 0.88.
2. Second, it iteratively refined the clustering using a combination of k-centroid clustering and densely connected-component (dcc) analysis at a stricter similarity threshold of 0.92 until convergence. During this refinement, the tool enforced a minimum similarity improvement of 0.02 to trigger pattern reassignment and maintained a strict boundary to prevent the co-clustering of monomeric patterns with composite motifs.

Throughout the process monomers and composite motifs clustered separately, as were motifs coming from signed and unsigned output types.

Following initial assignment, we performed a post-clustering check to identify any patterns that exhibited higher similarity to a centroid of a different cluster than the one they were assigned to, marking them for potential reassignment. A second filtering round using the same metrics as above and a common set of thresholds was then applied to remove any remaining noisy patterns that had been improperly aggregated during clustering.

Finalized motifs were then annotated using reference motif databases (e.g. JASPAR, ENCODE) using similarity 0.85 for primary matching and similarity 0.7 for secondary annotation for composite motifs.

The identified motif instances were subsequently mapped to the unified Motif Compendium clusters (translating pattern_id to cluster_id). During this step, the appropriate coordinate shifts and trims were applied to align the instances with the cluster centroids.

The final clusters were annotated by comparing their centroids against reference TF databases to assign biological labels. These were then used to annotate the FiNeMo instances in the previous section.

##### Generating Motif Compendium Instance Calls from Original MoDISco Pattern Instance Calls

We translated the track-specific pattern instances into compendium cluster instances:

- **Coordinate Alignment:** Per-track pattern IDs were mapped to compendium cluster IDs. Because extracting exact shift coordinates directly from the compendium clustering was challenging, we computed the relative shift and orientation by scanning each per-track pattern against its compendium cluster centroid CWM. This alignment was determined using the continuous Jaccard similarity metric on the CWM. The resulting positional shift and orientation (forward or reverse complement) were then applied to translate and place the instance coordinates into the same reference frame as the compendium cluster centroid CWM.
- **Flank Trimming:** Flanking regions of the aligned instances were trimmed by applying the same motif trimming strategy to motif compendium motifs that we did to TF-MoDISco motifs using the methodology implemented in FiNeMO.
- **Post-hoc Rescaling:** For scorers that support active allele calculations, we generated an additional rescaled copy of the motif instances. In this copy, the global hit importance (defined on the original unscaled signal level) was multiplied by the regional active-allele rescale average factor. Conceptually, for these rescaled instances, the local RMS normalization factor described above was additionally multiplied by the regional active-allele rescale factor. This formulation is equivalent to running FiNeMo directly on the rescaled contribution scores, ensuring that global hit coefficients scale quadratically to maintain model reconstruction consistency. The only main difference was that we used the average active allele score as a scaling factor computed over 1 kb intervals tiling the genome (same intervals as used for FiNeMO) for non-gene specific scorers, and over the whole receptive field of AlphaGenome around the gene (+-500kb) for RNA-seq scorers, which are gene specific. While these two versions are not exactly identical, the differences in practice are small because the active allele scorer already aggregates model predictions over receptive fields of approximately 500 bp (or whole gene body), producing a spatially smooth signal that changes little under further local averaging.

Both the original non-scaled and rescaled versions of the motif instances are preserved and provided in the final resources. We note that this rescaling procedure was performed post-hoc (after motif discovery and instance calling) and was used both for visualization of the rescaled ISM scores and motif instance thresholding. For the scorers where the rescaled version by active allele does not exist, we used the unscaled version for motif interpretation throughout the manuscript.

##### Motif instance filtering

FiNeMo is highly sensitive and, when applied genome-wide rather than restricted to regions with high genomic signal, can call a large number of motif instances, including weak hits that may represent noise. To establish a more stringent threshold for what qualifies as a real motif instance, we implemented a refinement strategy leveraging the high-contribution regions selected for TF-MoDISco as a reference set. Specifically, because these regions represent the most informative loci for motif discovery, we rationalized that functional mapped motif instances located outside these regions should exhibit comparable importance to those inside and used the in-region ‘hit_importance’ distribution as a reference to determine the minimum threshold for retention.

Concretely, for each track and motif, we split all mapped motif instances into two groups based on whether their genomic coordinates overlap a high-contribution region. We then computed the empirical cumulative distribution function (ECDF) of ‘hit_importance’ separately within each group, evaluated at every per-instance score. To reduce sensitivity to individual instances, the ECDF values were aggregated into fixed-width (0.01) quantile bins, with the median out-of-region ECDF taken within each in-region ECDF bin. Next, we applied the Kneedle elbow-detection algorithm to the concave, increasing curve of binned in-region ECDF quantile (x) against median out-of-region ECDF quantile (y) to identify the point at which the out-of-region ECDF curve plateaus relative to the in-region ECDF, representing the score level above which out-of-region instances exhibit comparable ‘hit_importance’ to in-region instances (fig. S5A). The elbow point on this curve determined a ‘hit_importance’ score threshold for the track and motif at hand: all instances with scores at or above this threshold were retained, regardless of whether they fell in-or out-of-region, and the rest were discarded (fig. S5A). Partitions with fewer than 3 binned points or a curve with an ill-defined elbow point were not thresholded, and all their instances were retained.

Because RNA-seq ISM scores were mapped not only by motif identity, but also by associated gene, region overlaps were computed in a gene-aware fashion, classifying an instance as in-region only if it overlapped a high-contribution region for that same target gene. Additionally, we further stratified the cross-distribution comparison of ‘hit_importance’ scores by binned levels of overall gene expression, selecting the elbow point between the two mapping set distributions. Specifically, genes were ranked by their median rescale factor as a proxy for model-predicted expression and assigned to 50 equal-frequency quantile bins. Thresholding was then applied independently per track, per motif, and per expression bin, yielding expression-aware thresholds.

Thresholding was conducted under two rescaling strategies: (1) unscaled, using raw ‘hit_importance’ scores as output by FiNeMo, with high-contribution regions also selected using unscaled ISM scores, and (2) scaled, with both ‘hit_importance’ and high-contribution region scores multiplied by the active-allele average, emphasising instances in regions with high predicted activity. Overall, per-motif elbow thresholding consistently preserved the vast majority of in-region motif instances while selectively pruning out-of-region background noise across all evaluated scorers and rescaling strategies (fig. S5B).

Additionally, because many transcription factor motifs share high sequence similarity, a single genomic hit could plausibly match multiple motifs in our compendium. To account for this ambiguity, we did not restrict our analysis to the single best-matching motif reported by FiNeMo. Instead, we computed the ‘hit_similarity’ for all related motifs in the compendium that represented plausible matches for that location.

In addition to the track-specific elbow thresholds, we defined a global significance filter to label low-confidence instances. An instance is classified as low confidence (indicated by the low_hit_importance flag in the exported tables) if its hit importance is less than 10% of the expected Phred30 score for that track. This expected Phred30 score is defined as the track’s Phred30 y-axis calibration limit multiplied by the length of the motif template. This global filter is not applied by default to exclude hits from the final tables; instead, downstream users can choose to apply it by filtering on the low_hit_importance column.

#### Motif instance evaluation

##### Evaluating mapped motif instances against ChIP-nexus data

To validate the biological relevance of our motif mappings, we compared predicted instances of representative factors (CTCF, HNF4A, and CEBPG) in HepG2 cells against published high-resolution ChIP-nexus binding profiles (*25*). Motif instances in this analysis had the Phred30 global significance filter applied to scaled ISM thresholding. As a control, we further confirmed that the highest-contributing mapped motif instances removed by thresholding (described above) produced weak, non-specific binding footprints. As an independent control, we performed position weight matrix (PWM) scanning genome-wide using FIMO (*26*) and retained the highest-scoring PWM motif instances that did not overlap with FiNeMo mapped motif instances derived from AlphaGenome ISM scores. With these PWM-only mapped motifs, we verified that ChIP-nexus binding produced weak, non-specific binding footprints. Despite being conducted on a limited set of experiments, these high-resolution ChIP-nexus binding footprints validated that the thresholded motifs mapped by FiNeMO from AlphaGenome ISM scores are experimentally bound *in vivo* across multiple TFs.

##### UniBind motif recovery

To demonstrate that the mapped motif instances were of comparable robustness to existing methods, we benchmarked their ability to recover mapped UniBind motif sets (*24*) across reproducible TFs in HepG2, K562, and IMR-90 cell lines. We compared our FiNeMo motif mappings to two independent motif mapping approaches. As the first benchmark, we leveraged SHAP scores derived from ChromBPNet models trained on ATAC-seq data in HepG2 (ENCSR380YGX), K562 (ENCSR467RSV), and IMR-90 (ENCSR978WIX) cell lines (*22*), performing TF-MoDISco using tfmodisco-lite (v.2.2.1) (*15*) and CWM-scanning using BPReveal (v.5.1.0) (*21*) across provided accessible regions in order to map contribution-derived motifs, as previously described (*17*). As the second benchmark, we leveraged digital genomic DNase-seq footprinting motif maps (*23*) across these same cell types of interest. When examining the ability of each approach to recover binding-derived UniBind motif maps to the relevant TF/motif pairs across shared genomic regions, our final thresholded motif maps for the DNASE modality performed comparably to ChromBPNet motif maps and consistently outperformed digital genomic DNase-seq footprinting motif maps. This demonstrated that, for motif maps derived from accessibility genomics assays, our motif mapping approach is comparable to existing methods.

Because our motif mapping approach extends beyond accessibility-derived genomics assays, we also sought to examine how well our thresholded motif mappings recovered UniBind motifs across transcription factor binding (CHIP_TF), accessibility (DNASE), activation (H3K27ac CHIP_HISTONE), and gene expression (RNA_SEQ) regulatory modalities. Thus, we examined across six well-studied cell types (HepG2, IMR90, K562, H1, HCT-116, GM12878) how well motifs mapped across these different regulatory modalities recovered binding-derived UniBind motif maps. As one would expect, motifs mapped to explain binding (CHIP_TF) modalities were best-performing with median UniBind motif recovery rates around 80%.

##### Evaluating in-cCRE rates

In order to demonstrate that our thresholded mapped motif instances were biologically meaningful, we sought to show that they were mapped at higher rates to biologically relevant regions, thus represented as annotated candidate cis-regulatory element (cCRE) regions from ENCODE (*4*). We measured cCRE overlap rates across six cell types (HepG2, IMR90, K562, H1, HCT-116, GM12878) for regulatory modalities representing binding (CHIP_TF), accessibility (DNASE), activation (H3K27ac CHIP_HISTONE), repression (H3K9me3 CHIP_HISTONE) and gene expression (RNA_SEQ). Thresholded mapped motif instances had higher overlap rates with annotated cCREs than raw FiNeMo mappings, validating that our thresholding approach produced a higher confidence set of mapped motifs. We note that the regulatory modality representing repression in this comparison (H3K9me3 CHIP_HISTONE) had notably lower cCRE overlap rates, which is consistent with repressive domains being associated with heterochromatin genomic regions that are inactive and possess closed chromatin (*97*).

##### Validating CTCF motif instance specificity using ChIP-nexus

In order to demonstrate that genome-wide, AlphaGenome Atlas motifs can be mapped in a cell-specific fashion, we compared motif instance mappings of CTCF, CTCFL and CTCF-UpstreamCore for HepG2 and K562 cells, as cell-type-specific binding of CTCF has previously been reported (*27*, *28*). Motif instances considered in this analysis had the Phred30 global significance filter applied to scaled ISM thresholding and were required to be mapped both by the CTCF ChIP-seq and the DNase-seq assay modalities for the pertinent cell type. Mapped CTCF instances were stratified based on whether they were specifically mapped by either HepG2 or K562 cells, or whether their genomic coordinates were mapped by both cell types. Reads-per-million normalized CTCF ChIP-nexus experiments in HepG2 and K562 cells were used to validate the binding of the top 250 CTCF motifs. These top motifs were selected to be strongly bound (top 50th percentile of bound motifs by experiment) and then ordered by the most differential (cell-specific) or least differential (shared) binding effects. Individual instances of these cell-specific and shared CTCF motifs were then examined and showcased motif examples were selected based on their overlaps with TraitGym Mendelian and Complex variants (*30*).

#### Motif analysis

##### Manual annotation of motif clusters and consolidated motif labels

Following automated clustering with MotifCompendium, which yielded 2,601 non-redundant motif clusters, expert manual curation was performed to consolidate clusters into broader, biologically interpretable transcription factor families. Automated matches against reference databases (JASPAR 2026 and ENCODE compendia) at similarity thresholds of 0.70 and 0.85 were used as a baseline guide. During curation, 262 clusters identified as technical or spurious artifacts were filtered out. The remaining 2,601 clusters were partitioned into consolidated groups: 94 broad motif labels representing canonical main transcription factor families (including core splice regulatory motifs), 122 zinc finger transcription factors (including ZNF and ZBTB families), and 464 composite motif patterns representing 104 distinct dimeric motif combinations. Clusters lacking confident matches to reference databases were categorized as unannotated motifs.

##### Motif compendium UMAP embedding

To map the global sequence and structural diversity of the motif compendium, we constructed a two-dimensional Uniform Manifold Approximation and Projection (UMAP) embedding of all 2,601 non-redundant consensus motif clusters in the atlas. Pairwise similarity scores were extracted from MotifCompendium, and the pairwise distance matrix was defined as D = 1 - similarity, setting diagonal self-distances to zero. The 2D manifold was embedded using UMAP with hyperparameters n_neighbors = 60, min_dist = 0.2, spread = 1.0, and random_state = 42.

##### Motif prevalence across AVI PHRED bins

For the motif overlap analysis in Fig. 5C, non-coding SNVs (excluding GENCODE v46 protein-coding CDS intervals) were evaluated for overlap with significant motif instances. Because the compendium spans hundreds of experimental tracks across diverse cell types, querying all motifs genome-wide would cause many variants to overlap a motif purely by chance. To ensure functional relevance, each variant was evaluated for motif overlap specifically within the experimental track contributing its largest absolute feature attribution. Overlaps were partitioned into splicing motifs (motifs identified in splicing assays, such as splice sites and splice regulatory elements) and non-splicing motifs (motifs identified in transcription factor and chromatin accessibility assays), and reported as the percentage of all variants within each PHRED bin.

##### Base-resolution AVI motif footprint profiling

To profile positional impact across binding sites, base-resolution feature attributions and AVI Phred scores were evaluated across the 30 bp motif window for all high-confidence instances (relative hit importance >= 0.1). Instances on the negative strand were reverse-oriented so relative positions 0–29 consistently run 5’ to 3’ to align with the motif CWM. At each nucleotide position, we computed the mean absolute SHAP value across modalities and the median AVI Phred score with interquartile range (25th–75th percentile whiskers), plotted alongside the aligned CWM sequence logo.

##### Identification of top motifs per AVI feature modality

To identify key sequence motifs driving each model modality, feature attributions were calculated by averaging absolute SHAP values across the 30 bp window and across all high-confidence genomic instances with >=100 instances per cluster. For each of 15 prediction modalities (Stop Lost was omitted due to near-zero signal), the top 5 motif clusters with the highest mean attribution were selected. Horizontal bars are color-coded by coarse functional annotation class, accompanied by motif CWM sequence logos.

##### *PLA2G7* locus motif disruption and alternate allele ISM

To determine how rare variants disrupt regulatory grammar at the PLA2G7 promoter (Fig. 4G), ISM was performed directly on sequence backgrounds carrying the alternate variant alleles (ALT ISM) across macrophage CAGE-seq tracks (CL:0000235). For both SNVs and the 4 bp insertion (chr6:46735442:G>GTACC), single nucleotide substitutions were evaluated across the mutated sequence window (chr6:46735429–46735461) to generate alternate contribution matrices. Scores were rescaled by the local active-allele average (times active allele) and plotted with synchronized y-axes to directly compare motif disruption between reference and alternate sequences.

##### Cross-modality and lineage motif attribution profiling

To systematically link sequence motifs to the specific modalities in HepG2 cells, motif instances contributing to HepG2 RNA-seq predictions were evaluated for co-occurrence across chromatin accessibility (DNase-seq) and active histone acetylation (H3K27ac ChIP-seq) in the same cell type. For each motif, individual genomic instances were classified based on which combinations of these three assay modalities mapped a shared motif instance at the same genomic coordinates, as well as the positive (activating) or negative (repressive) directionality of the predicted effect. The proportion of instances falling into each modality combination was quantified for each motif.

Motifs were grouped into three functional classes based on their instance distributions: Chromatin openers and activators (dominated by instances driving accessible chromatin, H3K27ac, and transcriptional activation, or acting primarily at the level of RNA expression like TATA), Dual activators and repressors (displaying substantial fractions of both activating and repressive instances across different genomic contexts), and Repressors (predominantly associated with closed chromatin, loss of active histone marks, and transcriptional repression).

##### Global Motif Activity Heatmap

To systematically profile regulatory motif activity across human tissues, cell types, and functional assays, we constructed a comprehensive compendium heatmap detailing regulatory motif dynamics. Heatmap values were aggregated from track-by-motif summary statistics utilizing high-confidence, filtered motif instances (see Motif Threshold Calibration and Filtering section above). These values account for 285 biosamples (functionally grouped into ten canonical tissues and cell identities) across 17 genomic assays. Motif activities can be evaluated at two resolutions: a broad view aggregating motifs into coarser, manually annotated motif clusters (usually representing major transcription factor families), and a granular view profiling individual motifs.

We quantified motif activity primarily using the hit importance score (described above) across all motif instances. To provide additional context, we included two supplementary metrics in the interactive dashboard: amount (the log-transformed instance count) and fraction (the instance count for a given assay normalized to the motif’s maximum observed element count across all assays within the same biosample). To equalize dynamic ranges across diverse genomic modalities while preserving genuine biological variance, we scaled values from 0 to 1 per assay. This was achieved by clipping at empirically determined percentiles of non-zero activities to prevent large outliers from compressing the visual signal: the 99th percentile for chromatin accessibility and histone marks, the 95th percentile for transcription initiation, splicing, and 3D contact maps, and the 90th percentile for RNA expression. For chromatin and transcription assays, percentiles were calibrated exclusively on transcription factor motifs to prevent highly abundant RNA-binding motifs from skewing the scale.

Critically, averaging opposing regulatory signals can mask underlying biological complexity. We therefore tracked activating and repressive contributions independently. Rather than using a divergent scale where strong positive and negative signals might visually cancel each other out, we employed a continuous bivariate color map. This approach simultaneously blends positive activation (teal) and negative repression (vermilion), with overlapping regulatory effects aggregating towards dark slate and neutral inactivity remaining white.

To highlight lineage-specific regulatory programs, biosamples were arranged by their ten canonical tissues and cell identities and hierarchically clustered internally using eight core epigenomic assays. Motifs were then partitioned into three biological tiers based on their regulatory breadth across these ten tissue groups. For each motif, a lineage activity profile was calculated by averaging combined positive and negative activity (u + v) across core assays and biosamples within each lineage; a lineage was defined as active if its activity reached *≥* 20% of the motif’s maximum lineage activity. Motifs were classified as: (i) ubiquitous if active across *≥* 6 lineages with a maximum-to-mean activity ratio *≤* 2.2 (reflecting uniform activity across lineages); (ii) broad if active across *≥* 4 lineages; or (iii) lineage-specific for all remaining motifs. To form a diagonal staircase of cell-type specific regulatory programs, lineage-specific motifs were assigned to the lineage of their peak activity and ordered following the 10 developmental lineages. Finally, within each tier and staircase step, motifs were hierarchically clustered (Ward’s linkage, Euclidean distance) and ordered by their net regulatory polarity (Σ*u −* Σ*v*), segregating predominantly activating elements from those driving repression.

While the interactive HTML dashboard (Data S3) incorporates the complete compendium—spanning all 285 biosamples, 17 modalities, and the entire motif repertoire utilizing the comprehensive hierarchical 3-tier staircase described above—the heatmaps presented in the main text (Fig. 6C,D) visualize a curated subset of biosamples and motifs, with manually adjusted color limits. Specifically, the average motif instance contribution x was log-transformed and scaled as log(1 + x) / log(1 + vmax), with assay-specific saturation thresholds (vmax) calibrated as: 20.0 for DNase accessibility, 5.4 for H3K27ac activation, 2.0 for RNA-seq expression, 3.2 for H3K27me3 Polycomb silencing, and 7.1 for H3K9me3 heterochromatin silencing (Fig. 6C displays DNase, H3K27ac, and RNA-seq; Fig. 6D additionally incorporates H3K27me3 and H3K9me3).

#### Population-Scale Genetic Aggregate Testing for Circulating Proteins and Complex Traits

##### UKB WGS

The WGS performed for UKB had an average coverage of 32.5× using Illumina NovaSeq 6000 sequencing machines with 150-bp paired-end reads (*79*). The genome build used for sequencing was GRCh38: single-variant nucleotide polymorphisms and short indels were jointly called using DRAGEN 3.7.8.

##### UK Biobank Genetic data filtering

As previously described (*49*, *98*), we set any DRAGEN WGS genotype calls to missing if either the *sum*(*LAD*) < 8 (where LAD is local allele depth) per sample genotype or genotype quality (*GQ*) < 10 for each of the 154,430 project variant call formats (pVCFs) provided by UKB using bcftools v.1.2 (*99*). A multi-allele splitting procedure was applied and each variant was assigned a unique ID (CHR:BP:REF:ALT) before merging all VCFs per chromosome and indels were normalized and left aligned using bcftools based on a 1000G b38 reference available at https://ftp.1000genomes.ebi.ac.uk/vol1/ftp/technical/reference/GRCh38_reference_genome (accessed 30 March 2024). Each merged pVCF was then converted to plink (v.2.0) p(gen/var/sam) format (*100*).

##### Genetic Variant Annotation

Before applying Atlas scores, we annotated all genetic variants using VEP v110, LOFTEE (*78*) and UTRannotator (*101*) as in (*49*). Where possible, we assigned each variant to one of three classifications: coding, proximal-regulatory or intergenic-regulatory. A variant was classified as coding if it had a predicted impact on the coding sequence of any transcript; proximal-regulatory if the variant lay within a 5kbp window of the UTRs of a transcript, and was not already a coding variant in any transcript, and finally intergenic-regulatory if was not coding or proximal-regulatory (details below). We additionally tested variants in sliding windows of size 2000 base pairs, regardless of the number of variants in each window, with coding variants excluded to minimise hypothesis overlap.

As in (*49*), we then assigned each variant to groupings, which we refer to as masks, according to their predicted consequence and location. As a comparator to AlphaGenome Atlas, we used the same published variant scores applied in (*49*) to further group variants by deleteriousness and consequence:

1. **Genomic Evolutionary Rate Profiling (GERP)** (*102*) A measure of conservation at the variant level. Variants are classified as highly conserved if they possess a GERP score > 2.
2. **phastCons Score** (*103*) A window-based measure of conservation across species (either strictly mammalian phastCons30 or all species phastCons100). Analysis was restricted to non-coding genome windows (excluding any window containing an exon) scoring in the 99th percentile.
3. **Constrained Score** Calculated in 1 kbp windows based on the local mutability and observed mutation rate of each window. Analyzed windows required a constraint Z-score >= 4.
4. **SpliceAI Score** (*41*) Predicts the probability of a variant within a pre-mRNA region acting as a splice donor, acceptor, or neither. Variants were classified as high-confidence splice sites if they possessed a SpliceAI score > 0.50.
5. **Combined Annotation Dependent Depletion (CADD)** (*7*) Predicts the deleteriousness of a variant. Applied exclusively to coding variants; loss-of-function variants were considered only if tagged as high-confidence by VEP. Missense variants with CADD > 25 were segregated into a separate testing mask.
6. **JARVIS Score** (*51*) A machine learning model derived from constraint metrics, designed to prioritize non-coding genetic variation for association studies.

Each genome mask consisted of a number of variants with different consequences, based on their location, one of the above scores and/or predicted coding consequences. For example, for a variant to be classified as missense *CADD* > 25, it must change a codon of an exon of a gene transcript and be predicted to be highly deleterious by CADD.

##### Transformation of AlphaGenome Scorers for Aggregate Testing

We exclusively selected non-active AlphaGenome scorers for aggregate testing, as we wanted to test the effects of grouping specific *variants*, not positions; it is well established that a causal variant does not imply all variations at the same multiallelic site are causal. To test the tissue-specificity of AlphaGenome, for each variant and score we calculated the maximum absolute score across different subsets of tissues/cell-types (“biosample_name” in AlphaGenome metadata) we considered most relevant to a given phenotype. To re-calibrate these max-scores, we calculated the absolute-value centiles for each tissue-maximisation based on the set of values assigned to all variants on chromosome 2 observed in UKB. We then took forward only variants with an absolute predicted score passing the 99th centile for aggregate testing.

We then overlapped variants passing each score’s tissue-aggregation threshold with the region-based annotations detailed in Table S5 (disregarding other score-filtering from e.g. JARVIS) - see Table S6 for which scores were tested within which regions.

#### Association analyses

##### Statistical significance

Statistical significance was defined based on the minimum *P* value observed for a WGS analysis of 20 randomly generated normally distributed continuous traits, where we tested the following tissue-aggregations: ‘all’, ‘Proteins’ and ‘CONTROL’. The minimum *P* values for aggregate association analyses were treated as independent: *P* = 3.63 × 10^−9^. Given that this was more stringent than the simulation-derived p-value derived in (49), we employed this threshold for all analyses.

##### Human protein expression levels

Protein levels of 2,932 proteins for up to 54,219 UKB participants were profiled using Olink technology, as described in (*104*) by the UKB Pharma Proteomics Project. Quality control procedures were applied to the data before being made available for researcher use, including outlier removal. Protein levels were additionally *log*_2_ transformed before release. After quality control filtering, up to 54,189 individuals with protein expression data were taken forward for analysis. Sun et al. found no evidence of batch- or plate-confounding effects.

We performed aggregate tests within *cis*-loci (1MB for 2,028 proteins measured in UKB with >99% coverage at the gene and locus level, continuing on from (*49*) with a slightly lowered coverage threshold (99%), using regenie v4.1 (*105*). To define the *cis*-window, as previously described, we first mapped each protein to a protein-coding gene (see Supplementary Table 1 of (*49*) for a small number of exclusions) and for each gene determined the longest transcript recorded by Ensembl. Based on the longest transcript, we then defined the *cis*-window as a 1-Mb window either side of the 5’- and 3’-UTRs of the transcript gene (limited by the beginning and ends of chromosomes), as well as the variants within the coding and intronic sequences between the UTRs. All association analyses were corrected for age, sex and age^2^, UKB recruitment center (as a proxy for geography), the first 40 genetic PCs, WGS batch, Olink plate, fasting time and time since blood draw. As in (*49*), we additionally adjusted for coding variants annotated to the cognate protein-coding gene.

##### Complex Traits

For non-protein complex traits, we performed a genome-wide association analysis, as per our previous work (*53*). We did not adjust for any coding variation by default. Association analyses were corrected for age, sex and age^2^, UKB recruitment center (as a proxy for geography), the first 40 genetic PCs and WGS batch. As in (*53*), height was rank inverse-normalised at run time, whilst we tried to account for the skew in BMI by applying the –mcc test-statistic correction implemented in regenie. To further insulate against the effects of phenotype skew, we performed a second round of association testing for studywide conditionally independent rare-variant BMI aggregates with rank-inverse normalised BMI, and only considered an aggregate as a novel finding if it also had *P*_RINT <1e-5. HbA1c was rank-inverse normalised.

##### Rare variant genomic aggregate testing

We followed the same methodology as (*49*) to identify conditionally independent aggregate tests. Briefly, to generate our primary discovery results, we adjusted for the common lead variants identified as independent signals in the joint (CoJo) analysis (at MAF > 0.1%, ∼MAC = 40). We did not condition on rare single variants from the offset.

Per phenotype, we identified independent aggregate associations if at least one aggregate passed our significance threshold. We performed a forward stepwise regression, starting from the most strongly associated aggregate (by *P* value) per phenotype and chromosome, and then performed an additional aggregate analysis for aggregates reaching genome-wide significance, adjusting for all variants in the top signal. This process was repeated, with more variants added from the next most strongly associated aggregate, until no aggregate existed that had genome-wide significance. As a sensitivity step, to establish which aggregates are not driven by singular single variants which we had power to detect, we performed a further step where we additionally adjusted the conditionally-independent aggregates for rare (MAF<1%) single variants from (*49*).

Gene unit testing was performed for variants with a maximum per-variant allele frequency threshold of 0.1%, using regenie v4.1, based on the genetic units specified. Regenie performs four types of genome unit tests: (1) standard BURDEN tests, under the assumption that each variant in a given gene unit mask has approximately the same effect size and sign on the phenotype; (2) SKAT tests, where the sign of association of each variant in the unit is allowed to vary; (3) ACAT tests, where the sign of association of each variant in the unit can differ and only a small number of variants in the mask need be associated; and (4) ACAT-O, which is an omnibus test of BURDEN, SKAT and ACAT that aims to maximize the statistical power across the three tests.

We performed each of the four statistical tests above for each mask for which a gene unit has at least one variant. In addition, an association test was performed for all singleton variants (with *MAC* = 1) in each unit; regenie also estimated an ‘all-mask’ association strength for each genome unit, which is an aggregation of the test statistics of the individual masks. To ensure that this did not result in a mixing of noncoding and coding association statistics, we split each gene transcript into a coding transcript, which we tested for all coding masks, and a proximal transcript, which we tested for all proximal masks. Regulatory genome units were classified by their ENSR assignment, the extent of a 1-kb constrained window or a phastCons-conserved window. We named sliding-window masks by the region of the respective chromosome that they covered.

##### Replication in All of Us

The whole genome sequencing performed for AoU had average coverage 37.9x using Illumina NovaSeq 6000 sequencing technology. The genome build used for mapping was GRCh38: single variant nucleotide variants and short indels were called using DRAGEN 3.7.8. Joint calling was performed via the Genomic Variant Store (*80*). The full AoUv8 WGS data was provided exclusively in Hail VDS format (*106*), with VETS filtering flags and site-level filters as described in the documentation (https://support.researchallofus.org/hc/en-us/articles/29390274413716-All-of-Us-Genomic-Quality-Report). We exported the Hail VDS to VCF format using Hail v0.2.134-952ae203dbbe, filtering on VETS at the variant-level, and additionally filtering genotypes (GQ<20 or sum(LAD)<8) as previously described (*49*, *98*). We performed association analyses for conditionally independent aggregates passing studywide significance for each complex trait, stratified by genetically-inferred ancestry. We performed a fixed-effect meta-analysis across the genetic strata, using the metafor R package (*107*), the result of which is reported as replication.

#### Experimental splicing screen of DNM1 intron

Methods are based on the work in (*40*).

##### Mammalian cell culture

HEK293FT cells (referenced as HEK293 in the text and figures, Thermo Fisher Scientific, R70007) were cultured in Dulbecco’s Modified Eagle Medium with GlutaMAX (Gibco, 10564011) supplemented with 10% (v/v) fetal bovine serum (FBS) (Sigma-Aldrich, F4135) and 1x penicillin-streptomycin (Gibco, 15140122). All other cell lines were obtained from the PRISM Platform at the Broad Institute. The PRISM cells were cultured in RPMI 1640 medium (Gibco, 11835030) supplemented with 10% (v/v) FBS and 1× penicillin-streptomycin-glutamine (Gibco, 10378016). Cells were maintained at confluency below 80%–90% at 37°C and 5% CO2.

##### Transfection and electroporation of splicing libraries into cell lines

The day before transfection, adherent cells were seeded into 6-well (HEK293) or 10-cm plates (all others, Corning) at a density so that the cells are approximately 70% confluent 24 h later. Cells were transfected using TransIT-X2 (KELLY, T47D), or TransIT-2020 (A172, DAOY, GAMG; Mirus Bio), following the manufacturer’s instructions, with three biological replicates per condition. Following transfection, cells were cultured for 48 h prior to RNA extraction.

##### Cloning of splicing libraries

Library elements were purchased as an oligonucleotide pool (Twist Biosciences) and amplified using KAPA HiFi ReadyMix Hot Start (Roche, KK2601). Cloning of the libraries happened using two steps. First, the oligonucleotide pool was cloned into two different custom-designed parent plasmid vectors that contained the sequence from *DNM1* exon 9 through 265 bp upstream of exon 10a using NEBuilder HiFi DNA Assembly Master Mix (NEB, E2621L). The two different parent plasmids were identical except for the upstream promoter driving the splicing reporter, which contained either EF1a or SFFV. The libraries were transformed into Endura Electrocompetent Cells (Sigma-Aldrich, LGC6024221) and plated onto two 30-cm carbenicillin-containing agar plates to grow overnight. The next day, cells were scraped off and the plasmid DNA was harvested using the ZymoPURE II Plasmid Maxiprep Kit (D4203). To ensure sufficient library diversity, the total number of colonies was estimated using serial dilutions of the transformed bacteria. After the first transformation, a second cloning step was performed to add in the 10a exon sequence using a gBlock (Twist Biosciences) and the NEBridge Golden Gate Assembly Kit (BsmBI-v2, E1602L). The same transformation protocol was used again.

##### Extraction of RNA and next-generation sequencing of amplicon splicing libraries

Total RNA was extracted from transfected or electroporated cells using the RNeasy Mini Kit (Qiagen, 74106), with on-column DNase digestion (Qiagen, 79254) to remove residual plasmid and genomic DNA. cDNA synthesis was performed with Maxima H Minus reverse transcriptase (Thermo Fisher, EP0743) using a library-specific primer containing a 10-nt unique molecular identifier (UMI). cDNA was purified using 1.5x AMPure SPRI beads (Beckman Coulter, A63882). To avoid overamplification, the number of PCR cycles was predetermined by qPCR of cDNA (Luna Universal qPCR, NEB, M3003).

Libraries were generated using a one-step PCR protocol where cDNA was amplified directly with Titanium Taq (Takara, 639210) using a mixture of P5 primers and a P7 index primer, under the following cycling conditions: 95°C for 5 min; predetermined number of cycles of 95°C for 30 s, 66°C for 30 s, and 72°C for 1 min; final extension at 72°C for 5 min. PCR products were purified with a double-sided SPRIselect cleanup (0.5× reverse elution followed by 1.2× forward elution, Beckman Coulter, B23319). Primer sequences used are listed in Table S3.

Libraries were quantified with the Qubit 1X dsDNA High Sensitivity Assay Kits (Thermo Q32851) and checked for purity with Tapestation using the high sensitivity D5000 kit (Agilent, 5067-5592). Amplicons were pooled and prepared for sequencing on a NextSeq 1000 (Illumina) with paired-end reads (read1, 240bp; index1, 8bp; index2, 8 bp; read2, 60bp). 20% PhiX was added to the sequenced libraries. Reads were demultiplexed and analyzed with appropriate pipelines.

##### Processing of amplicon splicing library NGS reads

Raw paired-end FASTQ files were processed to splicing outcomes for each reporter element. For each read, R1 contained the reporter sequence spanning the exon-exon junctions, while R2 contained the barcode for each element. The barcode and UMI from each sequence was first extracted using UMI-tools (v1.1.2) using a whitelist of known barcodes. Reads not matching the barcode whitelist were discarded. A custom Python script was used to map each reporter sequence to its corresponding splicing outcome. UMI-level counts were deduplicated and aggregated to produce element-barcode count matrices.

Read 1 sequences were aligned to the reference sequences for Exon 10B (MIDDLE_EXON) and Exon 10A (DOWNSTREAM_EXON) using edlib semi-global alignment. Reads in which both exons were detected were classified as COMBO_EXON and excluded from downstream analyses. Reads corresponding to unspliced introns or other splicing events were also excluded from downstream analyses. For DOWNSTREAM_EXON reads, the shift of the observed 3’ splice site from the canonical Exon 10A 3’ splice site was determined, where negative indicates intronic sequence retained upstream of the canonical site, positive indicates 5’ truncation of Exon 10A, and 0 indicates use of the canonical splice site.

Cell line–promoter combinations were filtered to those with an average UMI count per element > 300 across all 3 replicates, resulting in 5 different passing cell line–promoter combinations. Library variants were then filtered to those with at least 30 UMIs across all splicing outcomes per replicate.

##### Quantification of alternative splicing rates

The alternative splicing rate per variant = [downstream exon reads with >= 2 offset from the canonical site] / [all downstream exon reads]. We consider only cases where the alternative splice sites are in the intronic region between exon 10b and exon 10a. Variants were filtered to require at least 2 valid replicates per element. The “mean alternative splicing rate” is first computed by taking the average across replicates, and taking the average across the 5 cell lines used. Variants are called “active” if they have both a mean alternative splicing rate of > 0.10 across all passing conditions and a minimum coherence of 0.5, where the dominant alternative 3’ splice site accounts for at least 50% of alternative splicing reads.

#### ChIP-nexus experimental details

##### Cell culture

Human cell line HepG2 (purchased from ATCC; negative for mycoplasma contamination) was cultured at 37 °C in 5% CO2 in EMEM supplemented with 1x GlutaMAX, 1x non-essential amino acids (NEAA) and 10% FBS. Human cell line K-562 (purchased from ATCC; negative for mycoplasma contamination) was cultured at 37 °C in 5% CO2 in Iscove’s Modified Dulbecco’s Medium supplemented with 1x L-glutamine and 10% FBS.

##### ChIP-nexus experiments

ChIP-nexus was performed following the most recent version of the protocol (08/12/2021 https://stowers-institute.files.svdcdn.com/production/files/20210812_ChIP-nexus-protocol-1.pdf) with minor modifications described below. For each ChIP-nexus experiment, 10 million cells were fixed with 1% formaldehyde in 1 ml culture medium and 9 ml PBS at room temperature for 10 min with rotation. Fixed cells were washed with cold PBS containing protease inhibitors and incubated in 300 µl ChIP A2 buffer (15 mM HEPES, pH 7.5, 140 mM NaCl, 1 mM EDTA, 0.5 mM EGTA, 1% Triton X-100, 0.5% N-lauroylsarcosine, 0.1% sodium deoxycholate, 0.1% SDS, supplemented with protease inhibitors) for 10 min on ice. Chromatin was sonicated using a Diagenode Bioruptor Pico2. For K-562 cells, sonication consisted of four cycles of 30 s on and 30 s off at HIGH power. For HepG2 cells, chromatin was sonicated for eight cycles of 30 s on and 30 s off at HIGH power, followed by an additional four cycles under the same conditions using the residual pellet resuspended in an additional 300 µl ChIP A2 buffer. Chromatin extracts were centrifuged at 15,000 × g for 1 min at 4 °C, and the supernatants from the same biological replicate were pooled and used for ChIP. For each ChIP-nexus experiment, 60 µl of Invitrogen Protein G Dynabeads were washed three times with ChIP A2 buffer, resuspended in 500 µl ChIP A2 buffer, and incubated with 10 µl antibody for 2 h at 4 °C with rotation.

Chromatin extracts were added to the antibody-bound beads and incubated overnight at 4 °C with rotation. Immunoprecipitated chromatin was washed with ChIP-nexus wash buffers A–D (wash buffer A: 10 mM Tris, 1 mM EDTA, 0.1% Triton X-100; wash buffer B: 150 mM NaCl, 20 mM Tris-HCl, pH 8.0, 5 mM EDTA, 5.2% sucrose, 1.0% Triton X-100, 0.2% SDS; wash buffer C: 250 mM NaCl, 5 mM Tris-HCl, pH 8.0, 25 mM HEPES, 0.5% Triton X-100, 0.05% sodium deoxycholate, 0.5 mM EDTA; wash buffer D: 250 mM LiCl, 0.5% IGEPAL CA-630, 10 mM Tris-HCl, pH 8.0, 0.5% sodium deoxycholate, 10 mM EDTA). End repair and dA-tailing were performed using the NEBNext End Repair Module and the NEBNext dA-Tailing Module. ChIP-nexus adapters containing fixed barcodes (CTGA, TGAC, GACT, and ACTG) and UMIs were ligated using NEB Quick Ligase, followed by barcode extension with NEB phi29 DNA polymerase. Samples were subsequently treated with NEB lambda exonuclease. After each enzymatic reaction, chromatin was washed with the ChIP-nexus wash buffers A–D followed by Tris buffer (10 mM Tris, pH 8.0). Chromatin was then eluted and reverse-crosslinked overnight in ChIP-nexus elution buffer (25 mM Tris, pH 8.0, 5 mM EDTA, 0.5% SDS) containing Proteinase K. DNA was purified by phenol:chloroform extraction followed by ethanol precipitation. Purified DNA was denatured and circularized using CircLigase ssDNA Ligase (LGC Biosearch Technologies). Libraries were generated using 16 cycles of PCR amplification with NEB Q5 High-Fidelity 2x Master Mix and primers including Illumina TruSeq indexes. ChIP-nexus libraries were purified using Beckman SPRIselect beads and sequenced on an Element Biosciences AVITI sequencing system using an AVITI 2×75 Sequencing Kit Cloudbreak Freestyle High Output configured for single-end libraries with 100 sequencing cycles.

##### Antibodies for ChIP-nexus

The following commercial antibodies were used in this study. CTCF ChIP-nexus experiments were performed using the Cell Signaling Technology rabbit monoclonal anti-CTCF antibody D31H2 (3418S, lot 6). CEBPG ChIP-nexus experiments were performed using the Atlas Antibodies rabbit polyclonal anti-CEBPG antibody (HPA012024, lot A113977). HNF4A ChIP-nexus experiments were performed using the Atlas Antibodies rabbit polyclonal anti-HNF4A antibody (HPA004712, lot 000035854).

##### ChIP-nexus data processing

Single-end sequencing reads from ChIP-nexus experiments were pre-processed by trimming off fixed and random barcode components from the sequenced reads, reassigning barcode information as read names for downstream processing. Next, reads were pre-trimmed for adapters using Cutadapt (v.4.2) (*108*) and aligned to the human genome hg38 using bowtie2 (v.2.3.5.1) (*109*). Alignments were then deduplicated from aligned coordinates and their corresponding barcodes. Coverage was generated pooling the first “stop” base of each deduplicated read, separated into two tracks by read orientation. Normalized reads-per-million (RPM) ChIP-nexus tracks were generated by scaling the aligned coverage to the weighted total reads as described previously (*110*). Data is available at GEO accession number: GSE343944.

## Data Availability

AlphaGenome Atlas and AVI predictions are provided for research purposes only. Not for use in diagnostic procedures for medical decision-making.

All primary experimental datasets utilized for the training and evaluation of AlphaGenome in this study were obtained from publicly accessible sources.

The Atlas datasets generated and analyzed in this study are distributed across specific access tiers and licensing models and available at https://alphagenome.google/downloads. They are currently limited to all valid genome-wide SNVs in hg38 with potential expansion to indels upon final publication. In particular, the static download of AVI (Tabix format) is permissively licensed for commercial and non-commercial use.

| Access Level | Data Type | Format / Availability |
| --- | --- | --- |
| Permissive | AVI Data | Static download (Tabix) |
| Non-Commercial | AVI Data | API |
| Non-Commercial | Splicing Data | Static download (Tabix), API |
| Non-Commercial | AVI Feature Attributions | Static download (Tabix), API |
| Non-Commercial | Raw Atlas Features | API only |

Motif compendium can be browsed in Data S3 and motif instances can be browsed through the online portal. Motif compendium and motif instances will be available for bulk download soon.

ChIP-nexus data is available at GEO accession number: GSE343944.

## Code Availability

AlphaGenome Atlas is available for non-commercial use via an online portal at https://alphagenom e.google/atlas, with an accompanying Python software development kit (SDK) to programmatically access the resource available from https://github.com/google-deepmind/alphagenome. The AVI model source code and weights will also be provided upon final publication.

## Acknowledgments

**UKB** This research has been conducted using the UK Biobank Resource under Application Number 103356. UK Biobank protocols were approved by the North West Multi-centre Research Ethics Committee (MREC) (REC reference 21/NW/0157, previously 16/NW/0274).

**All of Us** We gratefully acknowledge All of Us participants for their contributions, without whom this research would not have been possible. We also thank the National Institutes of Health’s All of Us Research Program for making available the participant data examined in this study.

**Others** Amanda Stafford created educational materials to support the Atlas website launch and had critical feedback on the manuscript.

Rachael Tremlett provided figure editing and compilation assistance to the manuscript. Kathryn Tunyasuvunakool provided technical reviews of the paper.

Chang Yun and Salil Deshpande created the MotifCompendium package which was used to annotate motifs, and Anshul Kundaje provided critical feedback on the manuscript.

Alexander Karollus, Samantha Bryen, Greg Findlay, Phoebe Dace, Kinga Bujakowska, Emma Sherrill, Aubrie Soucy Verran, Boxun Zhao, and Tim Yu gave scientific feedback on this study.

Risha Patel set up the Trusted Tester program that enabled us to collaborate with the external community on this manuscript, as well as securing funding for the collaborative work. Alisha Eastep, Doga Fadillioglu and Francesca Pietra provided contract operations support for the Trusted Tester program, license and agreements.

Anshul Kundaje, Brendah Namugamba, Cassie Gray, Daniel MacArthur, Lingyi Wang, Marc Mansour, Mohamad Hajjari, Mounica Vallurupalli, Philip Montgomery, Phoebe Dace, Roisin Sullivan, Sam Bryen and Teresa Niccoli tested early versions of the website.

Charlie Taylor, Raphael Aboyeji, Uchechi Okereke, Gemma Gibbs and Juan Mateos-Garcia supported the manuscript preparation and review process. Mariana Felix managed the operations of the Atlas API and Website team.

Alexander Kenzior and the staff of Sequencing Core at the Stowers Institute for Medical Research for the help with the ChIP-nexus experiments.

Gemini (Google) was used for assistance with language editing and improving the clarity of the manuscript.

## Funding

G.H. is supported by the MRC [UKRI327]. This study was supported by the Wellcome Trust [226083/Z/22/Z], Medical Research Council [MR/Y003748/1] and National Institute for Health and Care Research Exeter Biomedical Research Centre. The views expressed are those of the author(s) and not necessarily those of the NIHR or the Department of Health and Social Care. H.C. and C.A.L. are supported by NIH Grants P30CA008748 and R00HG012579. L.E.C, A.O’D-L, H.L.R. were supported by NIH/NHGRI Grant U01HG011755. L.E.C. was supported by a postdoctoral fellowship from the Manton Center for Orphan Disease Research. F.C. acknowledges support from the Searle Scholars Award, the Burroughs Wellcome Fund CASI award, the Merkin Institute, the New York Stem Cell Foundation (NYSCF). F.C. is an NYSCF Robertson Investigator. This study was supported by the Stowers Institute for Medical Research through J.Z.

## Competing interests

E.A., K.R.T, G.N., L.E.N., T.W., Ž.A, and J.C. have filed patent applications relating to AlphaGenome (PCT/US2026/017924, PCT/US2026/017862). C.A.L. is a consultant to Cartography Biosciences, unrelated to this work. J.Z. owns a patent on ChIP-nexus (no. 10287628). H.L.R. has received funding for rare disease research from Microsoft, unrelated to this work. A.O’D-L. has received research support from Pacific Biosciences, unrelated to this work. F.C. is an academic founder of Curio Biosciences, Alive Bio, and scientific advisor for Amber Bio. F.C.’s interests were reviewed and managed by the Broad Institute in accordance with their conflict-of-interest policies.

## Author contributions

The Atlas project was a large collaborative effort. Our author contributions follow the consortium model, which lists authors as members of working groups. Authors can belong to more than one group.

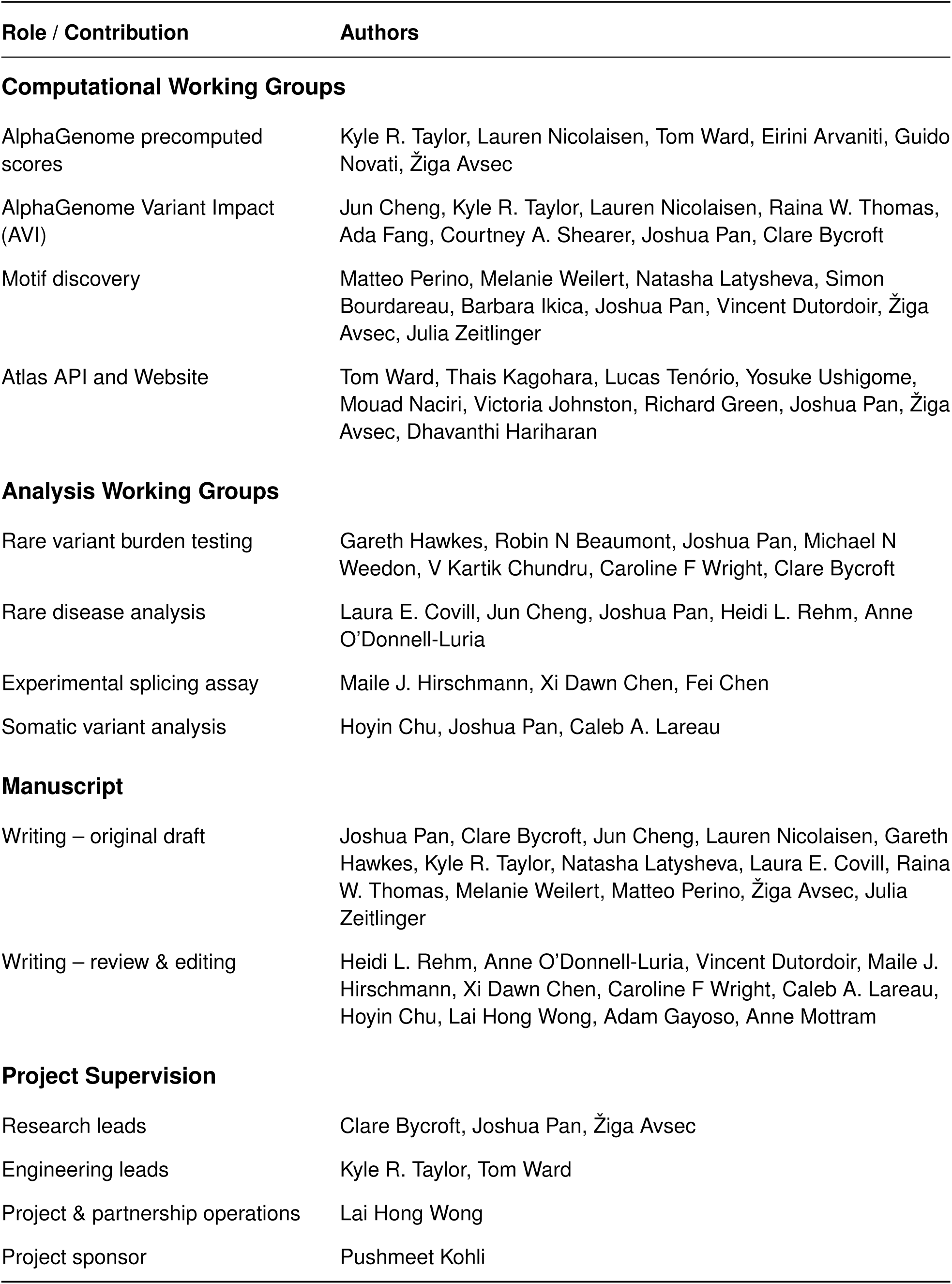

| **Role / Contribution** | **Authors** |
| --- | --- |
| **Computational Working Groups** |  |
| AlphaGenome precomputed scores | Kyle R. Taylor, Lauren Nicolaisen, Tom Ward, Eirini Arvaniti, Guido Novati, Žiga Avsec |
| AlphaGenome Variant Impact (AVI) | Jun Cheng, Kyle R. Taylor, Lauren Nicolaisen, Raina W. Thomas, Ada Fang, Courtney A. Shearer, Joshua Pan, Clare Bycroft |
| Motif discovery | Matteo Perino, Melanie Weilert, Natasha Latysheva, Simon Bourdareau, Barbara Ikica, Joshua Pan, Vincent Dutordoir, Žiga Avsec, Julia Zeitlinger |
| Atlas API and Website | Tom Ward, Thais Kagohara, Lucas Tenório, Yosuke Ushigome, Mouad Naciri, Victoria Johnston, Richard Green, Joshua Pan, Žiga Avsec, Dhavanthi Hariharan |
| **Analysis Working Groups** |  |
| Rare variant burden testing | Gareth Hawkes, Robin N Beaumont, Joshua Pan, Michael N Weedon, V Kartik Chundru, Caroline F Wright, Clare Bycroft |
| Rare disease analysis | Laura E. Covill, Jun Cheng, Joshua Pan, Heidi L. Rehm, Anne O'Donnell-Luria |
| Experimental splicing assay | Maile J. Hirschmann, Xi Dawn Chen, Fei Chen |
| Somatic variant analysis | Hoyin Chu, Joshua Pan, Caleb A. Lareau |
| **Manuscript** |  |
| Writing – original draft | Joshua Pan, Clare Bycroft, Jun Cheng, Lauren Nicolaisen, Gareth Hawkes, Kyle R. Taylor, Natasha Latysheva, Laura E. Covill, Raina W. Thomas, Melanie Weilert, Matteo Perino, Žiga Avsec, Julia Zeitlinger |
| Writing – review & editing | Heidi L. Rehm, Anne O'Donnell-Luria, Vincent Dutordoir, Maile J. Hirschmann, Xi Dawn Chen, Caroline F Wright, Caleb A. Lareau, Hoyin Chu, Lai Hong Wong, Adam Gayoso, Anne Mottram |
| **Project Supervision** |  |
| Research leads | Clare Bycroft, Joshua Pan, Žiga Avsec |
| Engineering leads | Kyle R. Taylor, Tom Ward |
| Project & partnership operations | Lai Hong Wong |
| Project sponsor | Pushmeet Kohli |

## Supplementary Materials

### Supplementary Text

#### Supplementary Note on UKB analyses

Unfortunately we were unable to complete our UK Biobank (UKB) analysis using the release-version of Atlas before the temporary downtime of the UK Biobank Research Analysis Platform (RAP) in April 2026. Instead, we have presented results from a slightly earlier internal release of Atlas used during our initial analysis. Before the downtime, we were able to extract marginal results for some of the tested aggregates using release-Atlas scores.

We found that the number of variants included in each aggregate was slightly higher in some release-Atlas aggregates selected from our list of presented conditionally independent results, where we were able to compare (202 aggregates), but still highly correlated with our presented data (fig. S10E; Pearson’s Correlation 0.769 [0.706, 0.820], *P* <1.11e-40). Despite these small differences in variant counts, we found that the release-Atlas aggregate association results were highly-correlated with our presented results (fig. S10D); Pearson’s Correlation 0.904 [0.875, 0.926], *P* = 1.30e-75).

Once access to the UKB Research Analysis Platform is restored, we expect to be able to update our results to use the released version of Atlas and revise this manuscript. Any unexpected changes in our conclusions will be highlighted at that stage.

---

**fig. S1.**
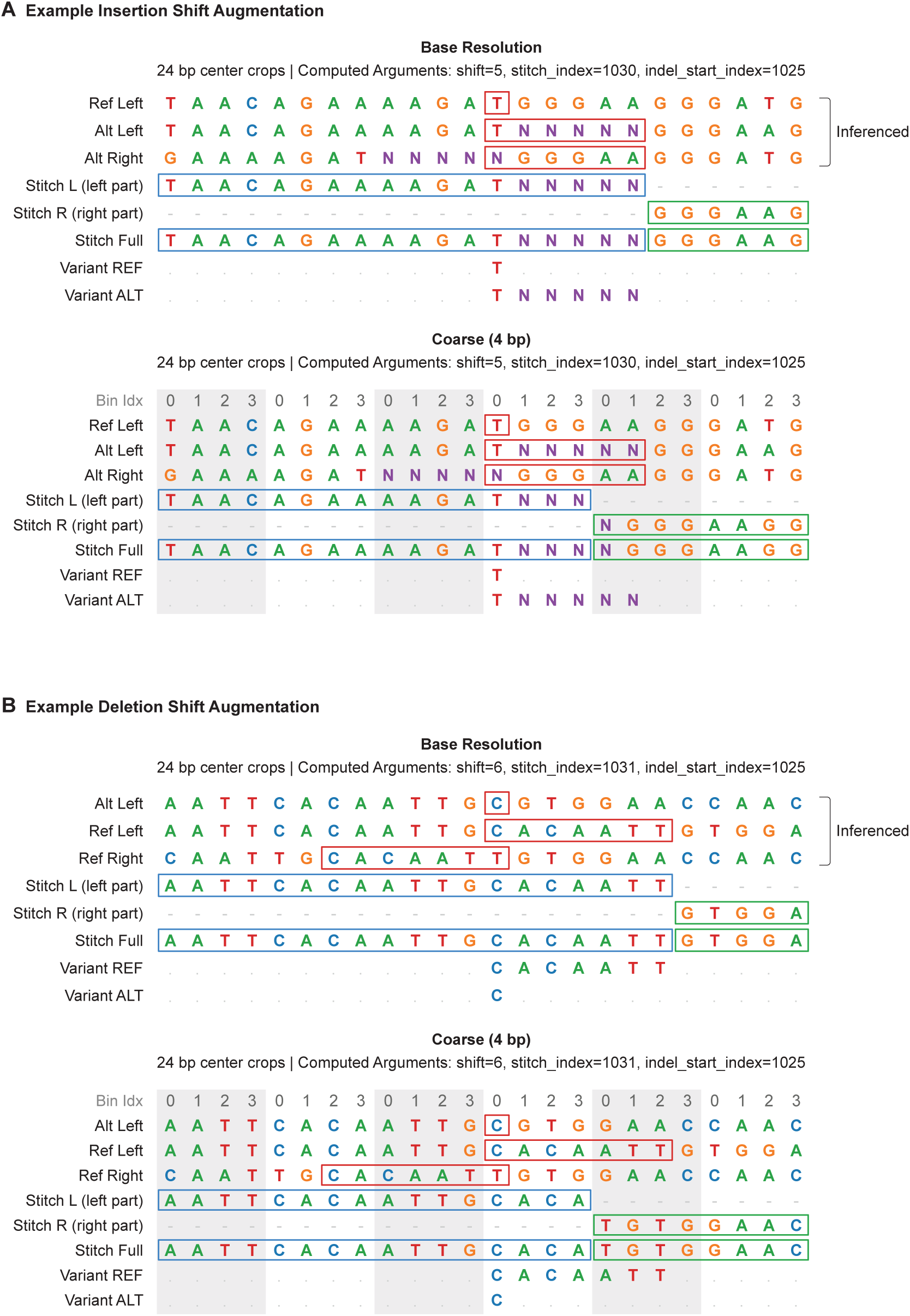
Indel shift augmentation to correct indel scoring artifacts. **(A)** Stitching for an insertion (chr7:3854822:T>TNNNNN, 5 bp insertion) at base resolution (1 bp, top) and coarse resolution (4 bp bin size for illustration, compared to 128 bp for the AlphaGenome model; bottom) across a 2,048 bp context (24 bp center crop shown). Inference is performed on the reference sequence alongside two alternate allele sequences (Alt Left and Alt Right), with the right-aligned sequence shifted prior to inference. Inserted bases are represented as N characters for visual clarity. Upstream predictions are taken from the left-aligned alternate sequence (Stitch Left, blue box), while downstream predictions are taken from the right-aligned alternate sequence (Stitch Right, green box) to restore positional alignment with the reference. At coarse resolution, predictions reuse the base-resolution inference, with stitching occurring at the enclosing bin boundary, which can result in minor boundary clipping of the inserted sequence. **(B)** Stitching for a deletion (chr7:13494802:CACAATT>C, 6 bp deletion) at base resolution (1 bp, top) and coarse resolution (4 bp bins, bottom). Deletions are considered as reference insertions, inferencing the Alt Left, Ref Left, and Ref Right sequences and stitching the reference across the deletion boundary after shifting to Ref Right. At coarse resolution, bin boundaries can cause the stitched reference sequence to differ slightly from the unsegmented reference.

**fig. S2.**
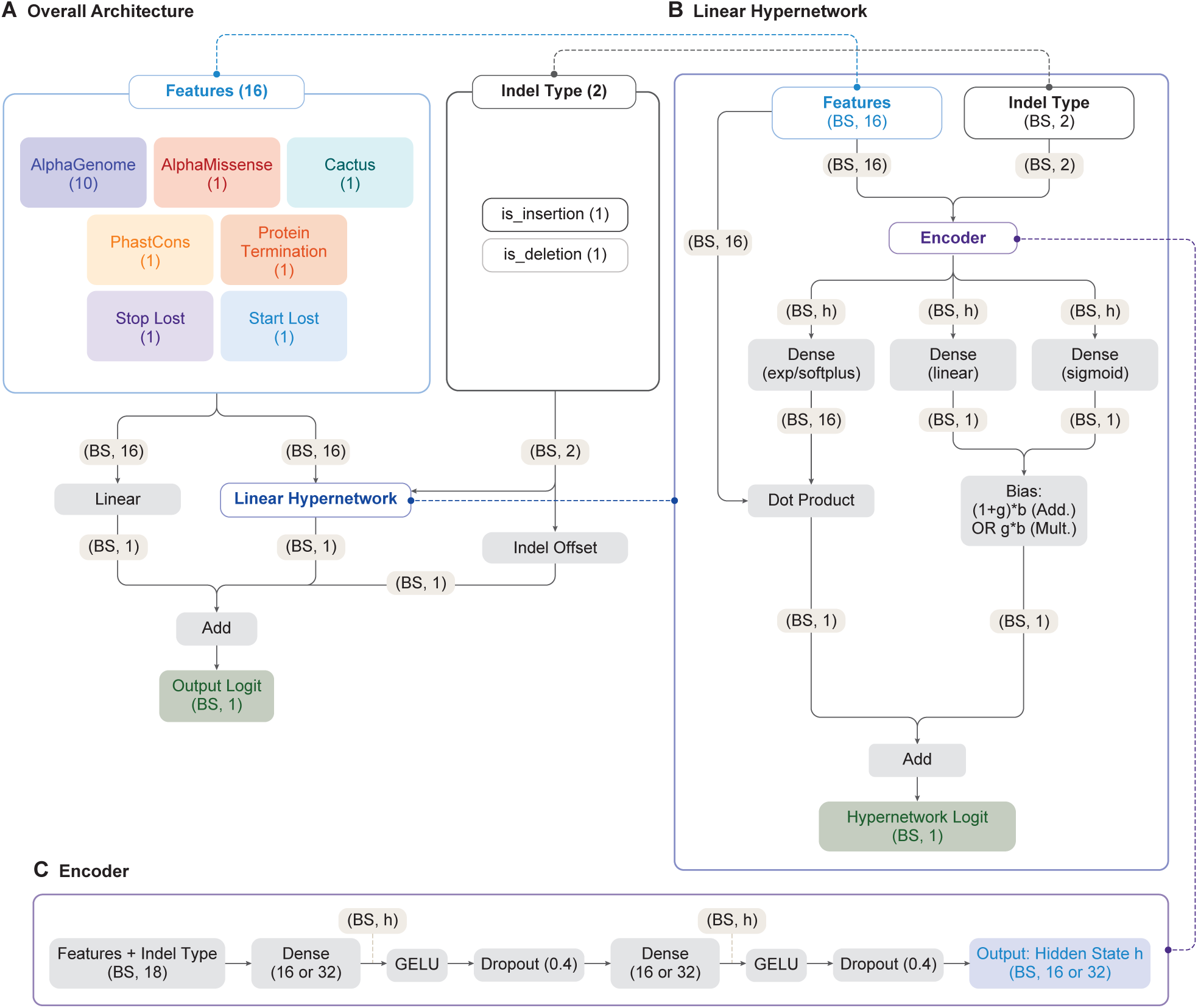
AVI model architecture. **(A)** The AlphaGenome Variant Impact model is a linear hypernetwork trained on 18 features. Ten features were derived from the AlphaGenome precomputed scores. The remaining features were derived from AlphaMissense, conservation scores, VEP consequence, and indel type. The two features indicating the indel type (binary indicators for whether the variant is an insertion or deletion) were added to enable the model to account for small distributional differences between single-nucleotide and indel variant scores. The indel type indicators v are used to condition the encoder and add a learned offset to the final output. See Methods for more details on model training, hyperparameters and evaluations. (BS = batch size, h = hidden dimension). **(B)** Linear Hypernetwork: The linear hypernetwork uses an encoder to dynamically generate the parameters of a linear layer (including a bias and bias gating term) based on the primary features x and indel type indicators v. The parameters are produced via separate projection heads branching from the output of the encoder. **(C) Encoder:** The encoder is a two-layer fully connected network (16 or 32 units), with GELU activations and dropout (rate = 0.4), that maps the concatenated input [x; v] to the hidden state h.

**fig. S3.**
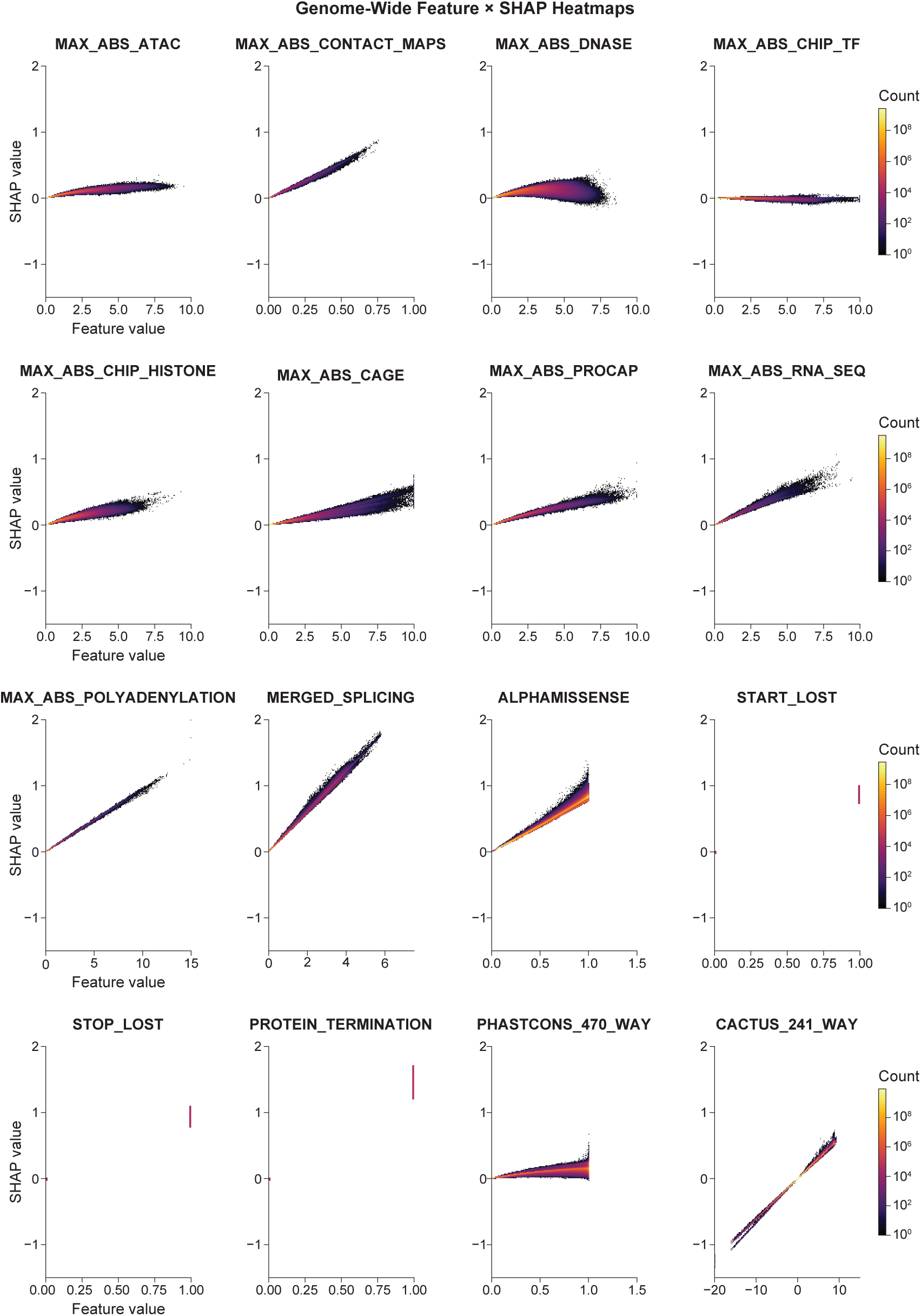
Feature dependence plots of AVI SHAP values versus input feature values. SHAP attributions are plotted against underlying input values for all possible genome-wide SNVs, faceted by feature. Attribution dependencies vary: features such as Splicing and Cactus exhibit linear proportionality between input magnitude and SHAP value, whereas other features exhibit higher variation. The three categorical VEP consequence features (start lost, stop lost, and protein termination) act as binary indicators conferring consistently high impact on the AVI score when active. Cactus is the sole feature utilizing negative inputs to encode evolutionary trajectory (positive: conserved; negative: fast-evolving).

**fig. S4.**
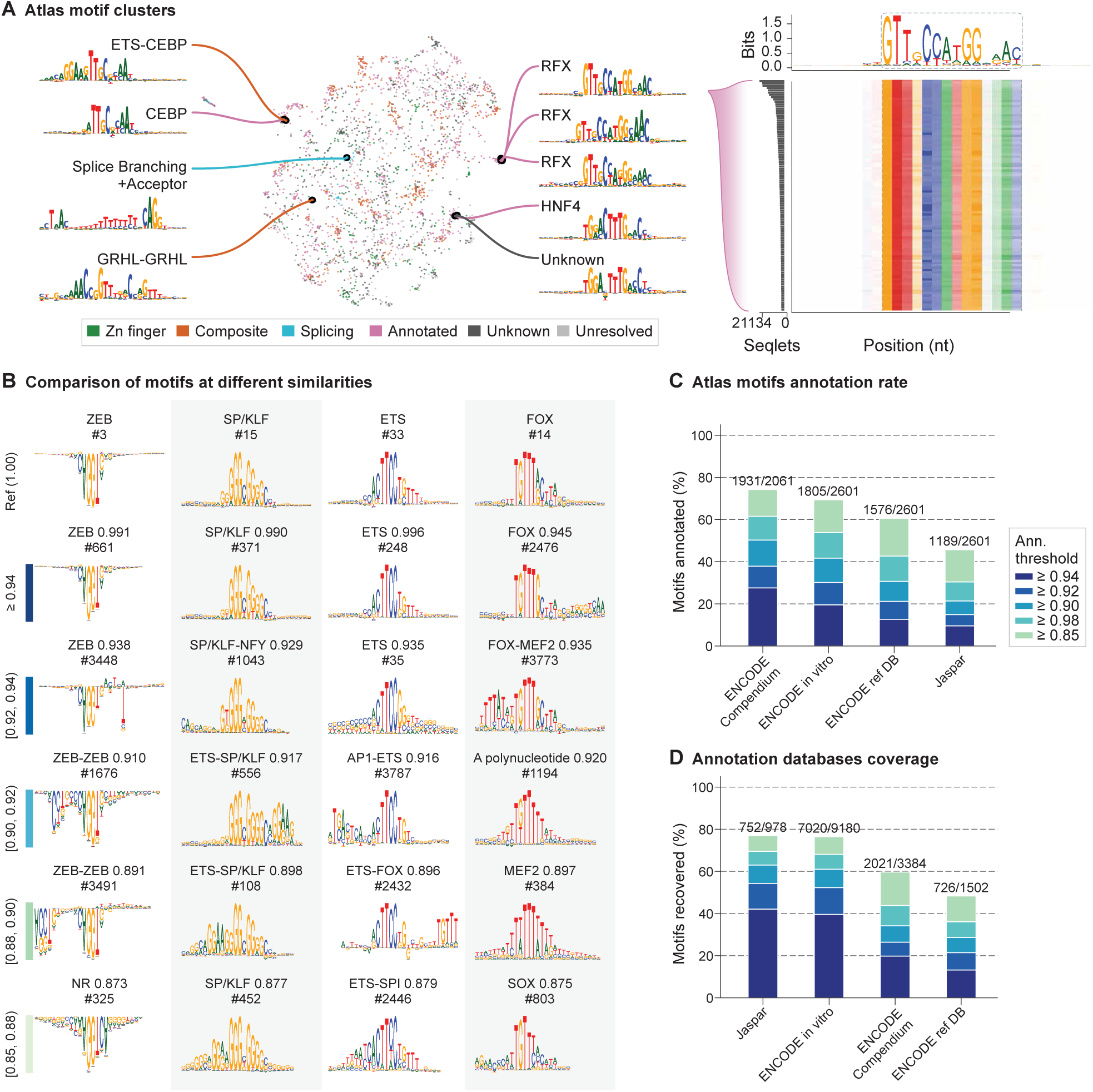
Motif similarity and annotation details. **(A)** Atlas motif clusters similarity. UMAP representation of motif distances (defined as 1-similarity) across the Atlas compendium, with highlighted examples. Motifs close in the UMAP space are similar to each other and highlight the similarity of motifs from the same family (RFX), between monomeric and composite motifs (CEBP, CEBP-ETS) and the potential use for identification of unannotated motifs (HNF4, Unknown). Heatmap inset shows the top 100 patterns, ranked by seqlet support, contributing to the indicated RFX motif. **(B)** Comparison of motifs at different similarities. Examples of motif similarity at different values. For each column the top row represents a motif taken as reference, and lower motifs are examples from the similarity range on the left. **(C)** Atlas motifs annotation rate. Number of Atlas compendium motifs annotated with the indicated database at multiple annotation thresholds. Thresholds reflect the upper boundary of the similarity brackets from B. **(D)** Annotation database coverage. Number of motifs in the indicated databases used to annotate at least one Atlas compendium motif at multiple thresholds (i.e. database recovery). Thresholds reflect the upper boundary of the similarity brackets from B.

**fig. S5.**
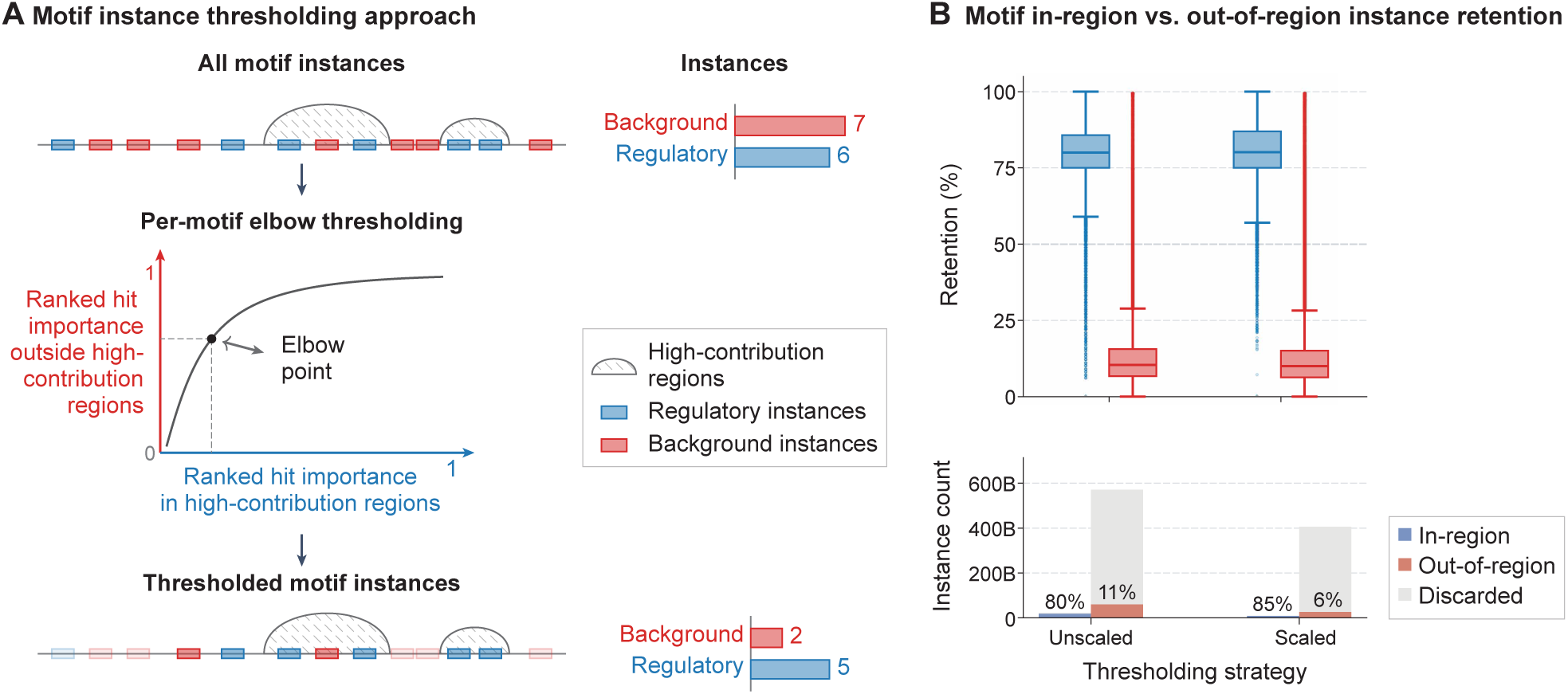
Motif thresholding. **(A)** Per-motif elbow thresholding for motif instance filtering. *(Top)* All initially mapped motif instances, categorized by regulatory status: regulatory instances (blue) represent biologically meaningful hits, while background instances (red) represent non-functional hits. High-contribution regions (hatched) are genomic intervals enriched for regulatory signals, used as calibration intervals for threshold selection. *(Middle)* Per-motif elbow point threshold derived from ranked hit importances inside versus outside high-contribution regions. *(Bottom)* The threshold selectively preserves high-confidence candidate motif instances while removing background noise. **(B)** Motif in-region versus out-of-region instance retention across thresholding strategies. *(Top)* Distribution of retention rates (%) across instances occurring within high-contribution regions (blue) versus outside them (red). *(Bottom)* Total instance counts per strategy within high-contribution regions (blue) versus outside them (red). The lower colored segment indicates retained instances with the corresponding percentage annotated above it, whereas the upper gray segment represents instances discarded by thresholding.

**fig. S6.**
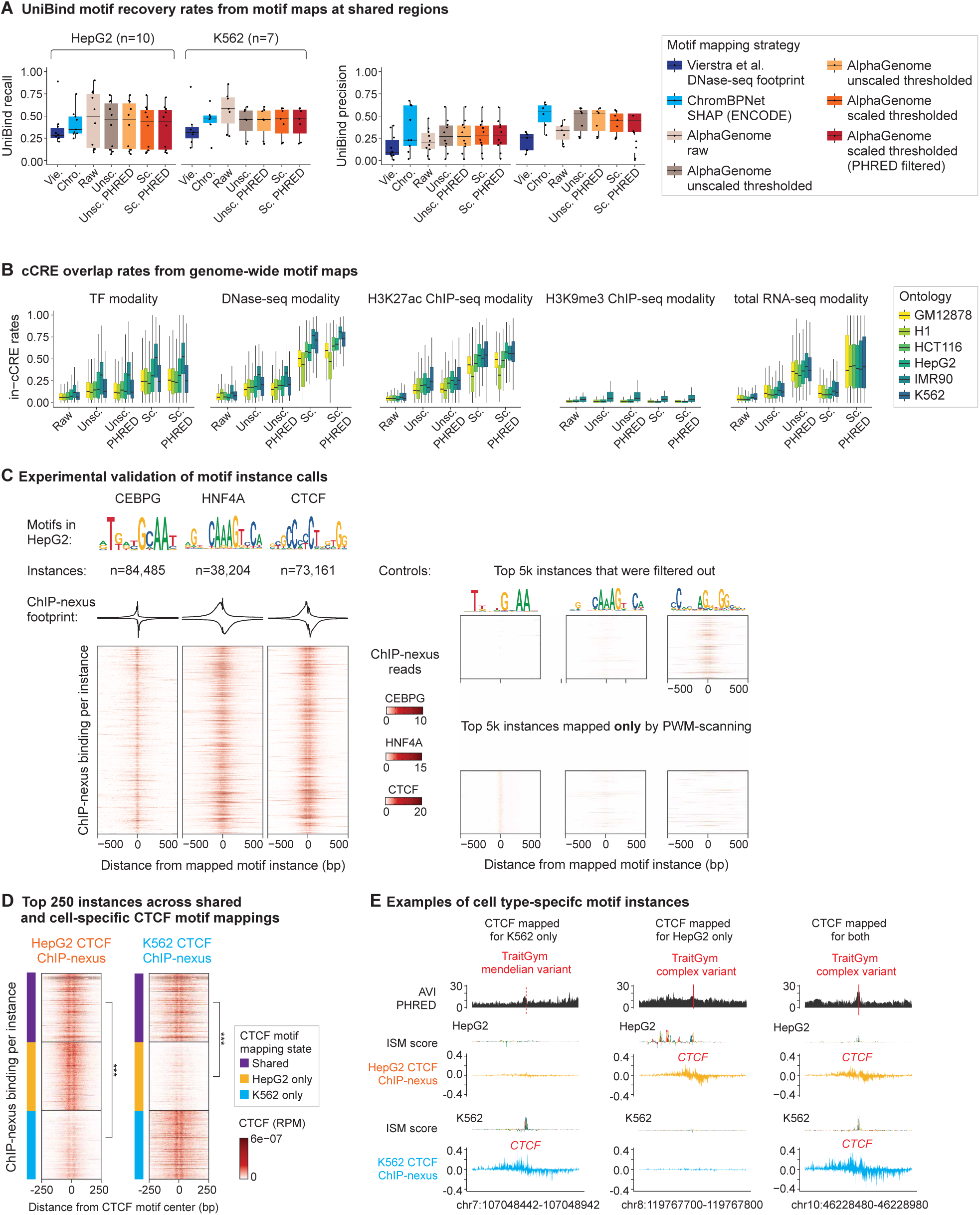
Validation of motif mappings. **(A)** Precision and recall motif recovery levels of reproducible UniBind TF motif maps, compared against motif maps (DNase-seq modality) produced by the AlphaGenome Atlas and controls (ChromBPNet and DNase-seq footprints (*23*)) across a shared set of regions in HepG2 and K562 cell lines. Each motif label that could be linked to a UniBind TF is treated as an observation. **(B)** Overlap rates of cCREs for motif maps produced by the AlphaGenome Atlas across key regulatory modalities. As expected, AlphaGenome scaled and thresholded motif maps outperformed unfiltered motif maps. We note that motif maps produced by the H3K9me3 modality produced low cCRE overlap rates, consistent with its role in marking repressive heterochromatin, indicating the AlphaGenome Atlas is mapping motifs in a modality-sensitive fashion. These comparisons were conducted across five modalities (TF ChIP-seq, DNase-seq, H3K27ac ChIP-seq, H3K9me3 ChIP-seq, and total RNA-seq) for six cell types (HepG2, IMR90, K562, H1, HCT-116, GM12878). Unsc., unscaled; sc., scaled. **(C)** Experimental validation of mapped motif instances (TF ChIP-seq modality) by high-resolution ChIP-nexus binding experiments (*25*) for CEBPG, HNF4A, and CTCF in HepG2 cells with corresponding PWM logos and motif instance frequencies reported. Controls are shown on the right: the top 5k of raw FiNeMO mapped motif instances that were filtered out (scored by instance importance) and the top 5k of motif instances mapped by genome-wide PWM scanning but not by the AlphaGenome Atlas (scored by PWM matches) here show minimal binding. To flatten outliers, all heatmap coverage is thresholded to the upper 90th percentile of ChIP-nexus coverage per TF/motif pair. **(D)** Top 250 CTCF motif instances (DNase-seq modality) that were either mapped for both HepG2 and K562 cell lines or in a cell-type-specific way to either cell line were confirmed by CTCF ChIP-nexus binding (*25*). To flatten outliers and compare across experiments, all ChIP-nexus experiments were reads-per-million normalized, then heatmap coverages were thresholded to the upper 90th percentile of ChIP-nexus coverage. Differences in coverage were confirmed by Wilcoxon rank sum tests (***, p < 3e-16). Cell-type-specific binding of CTCF has previously been reported (*27*, *28*) **(E)** Individual CTCF motif instances that were mapped (DNase-seq modality) in a cell-type-specific way (shown as DNase-seq ISM scores) were confirmed by CTCF ChIP-nexus binding in HepG2 and K562 cell lines. Shown instances overlap with TraitGym mendelian or complex variants and have typically higher AVI scores across CTCF motif instances.

**fig. S7.**
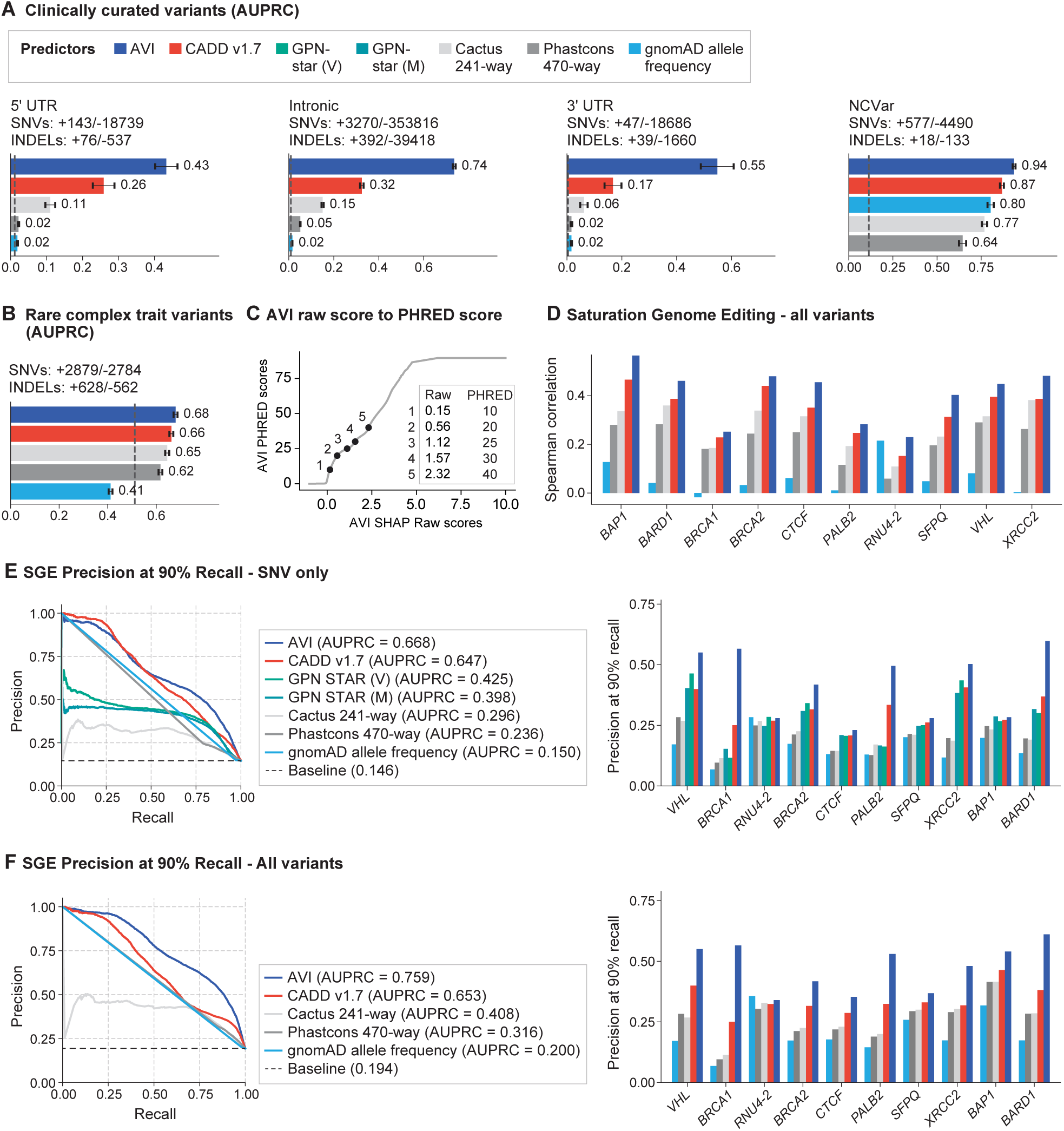
Additional evaluations of AVI performance. **(A)** Comparing AVI with other methods on clinically curated variants (AUPRC). Including ClinVar variants stratified by molecular consequences (5’UTR, intronic variants, 3’UTR), and non-coding variants from the ncVar database (which is mostly derived from ClinVar). **(B)** Distinguishing rare variants associated with complex traits from unassociated control variants (AUPRC). **(C)** Mapping AVI raw scores (equals to sum of SHAP scores per variant) (x-axis) to PHRED scores (y-axis). The raw scores corresponding to PHRED score 10, 20, 25, 30, and 40 are marked as dots on the curve. **(D)** Spearman correlation between model predictions and experimental readouts per protein, including all variants (SNVs and indels). **(E)** Performance of AVI and alternative methods on saturation genome editing SNV data. (Left) Precision-recall curves and AUPRC metrics for all screens pooled into a single evaluation. Note: 95% of the included variants lack a gnomAD allele frequency and were therefore imputed as zero. Similarly, Phastcons 470-way scores are largely binary, with approximately 50% of scores being one and 20% of scores being zero for variants included. (Right) Precision at 90% recall (which quantifies the false discovery burden incurred when a model is constrained to high sensitivity settings), computed independently for each screen. **(F)** Same as **(E)** but including all SGE variants (SNVs and indels) and a subset of models with indel scores.

**fig. S8.**
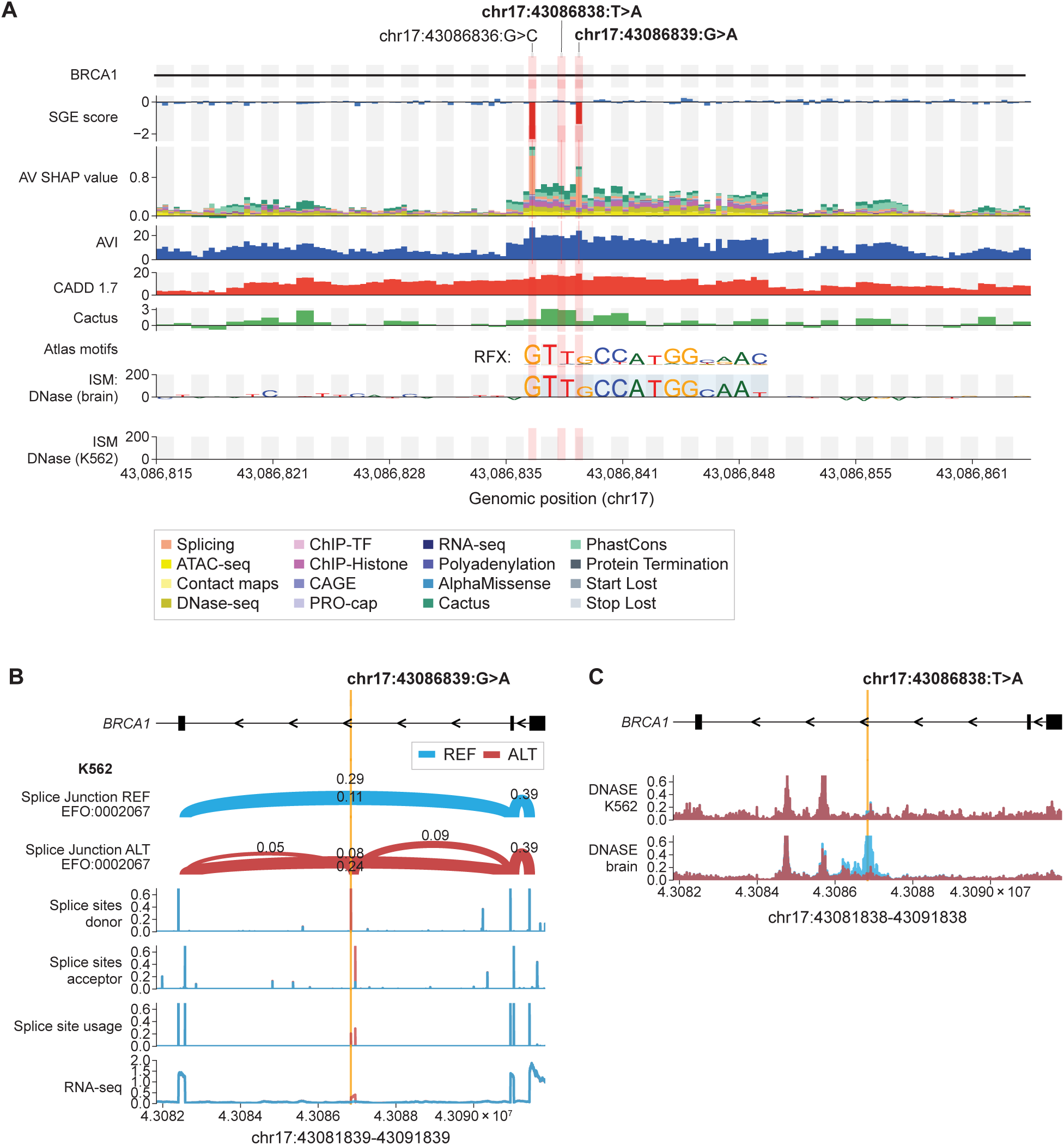
Example of *BRCA1* deep intronic variants / brain-specific enhancer. **(A)** An example region in the intronic region of *BRCA1* that was measured by the SGE experiment (Dace 2025). SGE scores (red for variants classified as functionally abnormal and blue as functionally normal) are shown together with AVI feature attribution, AVI scores, CADD v1.7 scores, Cactus conservation scores, motif instances and AlphaGenome DNAse Active ISM contributions scores in this region for brain and K562. Grey and white stripes mark base-pair resolution. For SGE and AVI scores the 3 data points within each stripe represent the 3 alternative bases at allelic resolution. Alternative variants are ordered alphabetically. **(B)** AlphaGenome splicing predictions for the variant chr17:43086839:G>A. Predictions for the reference sequence shown in blue and for the alternative sequence in red. Splice junction and RNA-seq predictions for K562 are shown. Splice site predictions are not tissue specific. Negative stranded-only scores are shown as BRCA1 is on the negative strand. **(C)** AlphaGenome DNAse predictions in the brain and K562 cell line for the variant chr17:43086838:T>A. Predictions for the reference sequence shown in blue and for the alternative sequence in red.

**fig. S9.**
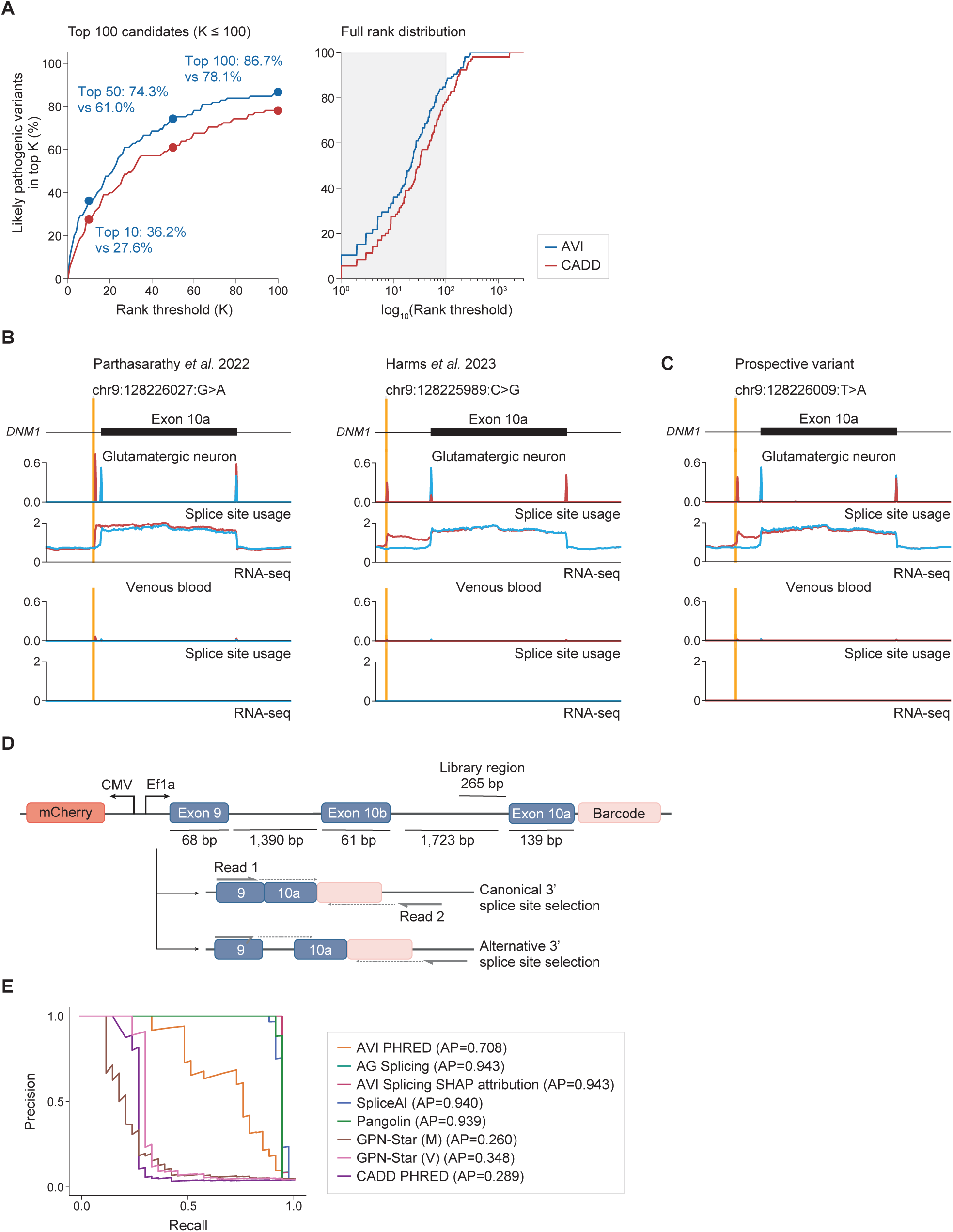
Additional results for GREGoR analyses and *DNM1* intronic variants. **(A)** Retrospective variant prioritization performance in the GREGoR cohort, accounting for allele frequency. AVI and CADD v1.7 were evaluated on 105 likely pathogenic variants from solved cases, ranked against intra-patient background variants filtered by an allele frequency cutoff of 0.001. The causal variant of seven cases in Fig. 3A did not pass the AF filter and were therefore removed. The likely pathogenic variants from solved cases were ranked against background variants in the same patient. Curves plot the cumulative recall (y-axis) of causal variants captured within the top K prioritized candidates (x-axis). The left panel bounds the evaluation at K = 100; the right panel displays the full rank distribution. **(B)** AlphaGenome tracks predictions for the reference and alternative allele of two other known in-frame exon extension variants affecting *DNM1* exon 10a. **(C)** A prospective variant with similar AlphaGenome predictions but previously unknown. **(C)** Experimental reporter design. 3’ alternatively spliced isoforms are quantified through sequencing, with read 1 spanning exon-exon junctions and read 2 capturing the barcode information. **(E)** Precision-recall plot of AVI features and feature attributions as well as other models, evaluated on their ability to separate active from inactive variants as determined by the experimental screen.

**fig. S10.**
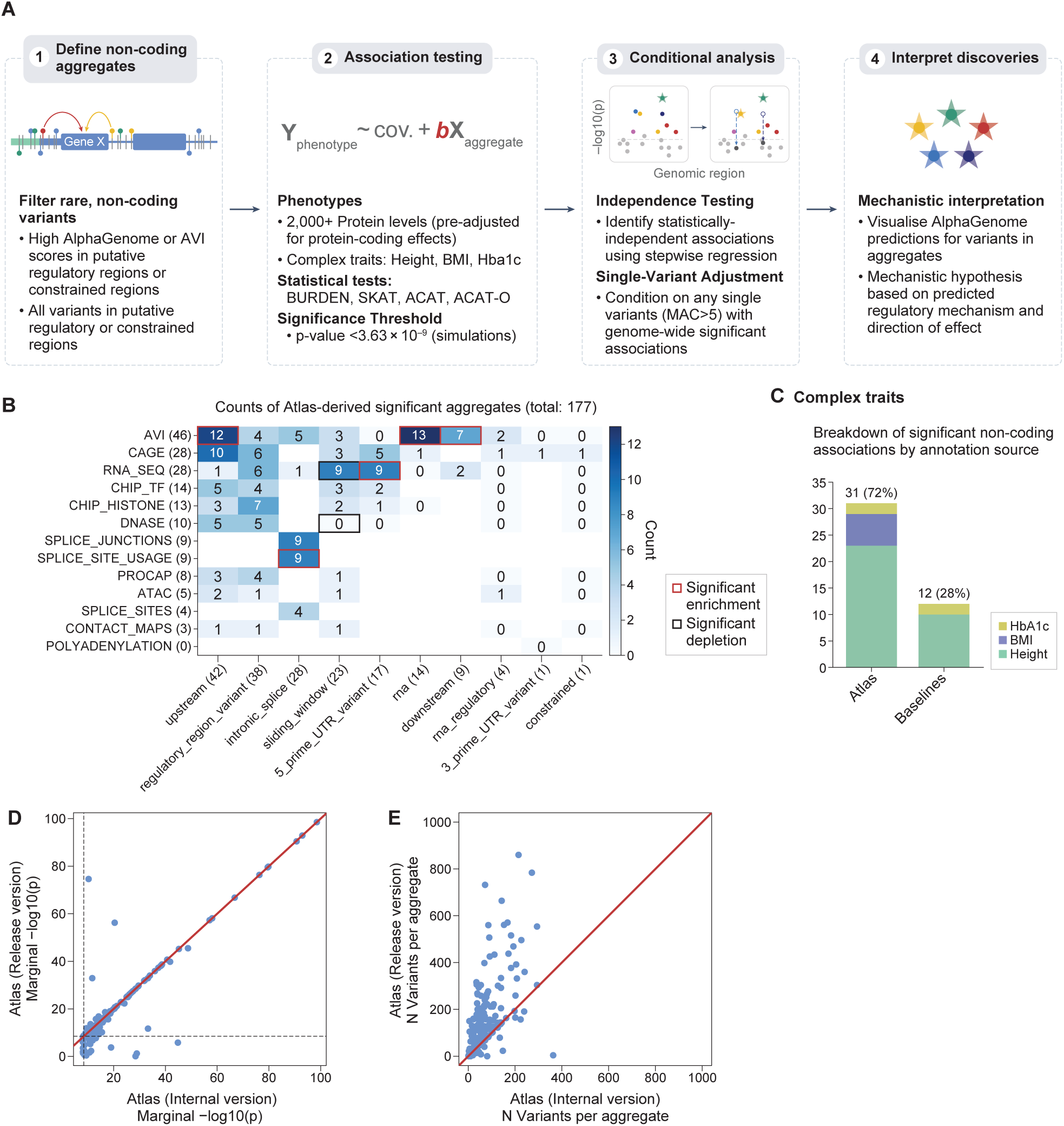
Additional results from rare non-coding burden testing analyses, including complex traits. **(A)** Overview of aggregate association testing pipeline (see Methods for more detail). Unless otherwise stated, results summarised in other figures used all of Steps 1-3 as described here. **(B)** Breakdown of significant, non-coding AG-driven associations for the protein abundance phenotype by annotation source. Marginal totals are shown in parentheses in axis labels. Bold squares indicate a significant enrichment (red), or depletion (black) compared to that expected based on the total number of aggregates tested (two-sided binomial test p-value < 0.000675). See Methods for details of each of the ‘regional’ (x-axis) and AlphaGenome-derived (y-axis) annotations. **(C)** Performance of the AG-Atlas annotations for association testing, after further adjusting for rare single variants associated with each trait, across complex traits: height, BMI, and HbA1c. *Note: as per our analysis of circulating proteins, we will repeat our complex-trait analyses once access to the UKB Research Analysis Platform is restored. In particular, for complex traits we were also unable to measure the impact of including the AVI score as a feature.* **(D)** Comparison between the strength of statistical association, where possible, for each aggregate in the presented data (internal version) and the published version of Atlas (Release version). Dashed lines represent marginal studywide significance (-log10(p) = 8.44). **(E)** Comparison between the number of variants contained within each aggregate in the presented data (internal version) and the published version of Atlas (Release version).

**fig. S11.**
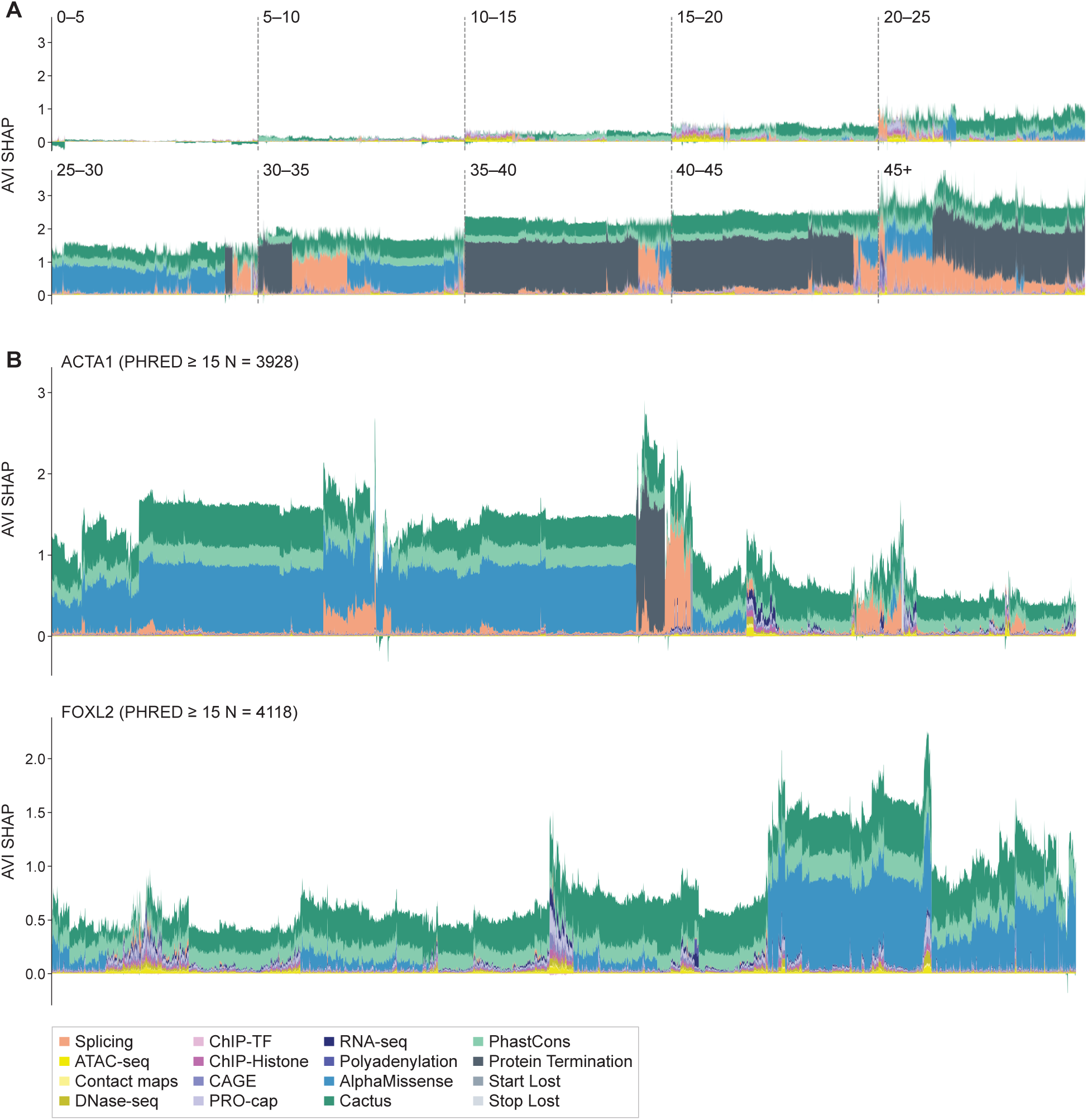
Streamgraphs of variants sampled from different AVI bins and from different genes. **(A)** In each AVI bin (width 5, except for the final open-ended 45+ bin), 10k individual variants were sampled and plotted with their feature attributions in a stacked bar plot. In each bin, variants are clustered along the x axis to highlight similar feature attribution groups. **(B)** Same but showing all variants with AVI PHRED score >= 15 in the gene region (200 bp upstream from the GENCODE GTF V46 Start to GTF End) for *ACTA1* and *FOXL2*. AVI feature attributions for these variants are plotted as a stacked bar plot and clustered along the x-axis to highlight variant groups.

**fig. S12.**
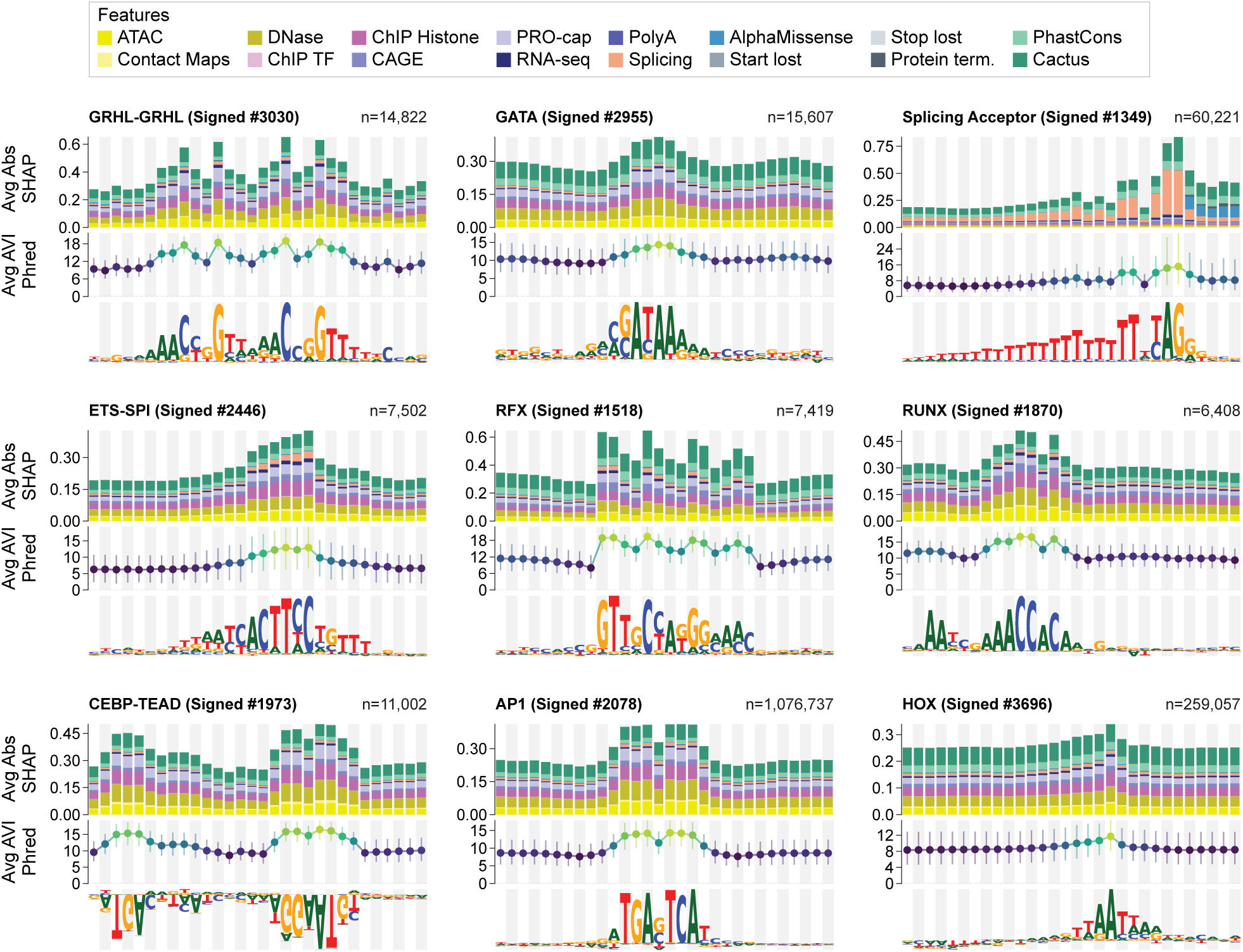
AVI and feature contribution footprint profiles of sample motifs. For each motif, scores are aggregated across all high-confidence genomic instances (total instances n indicated in the top right of each panel). Each motif profile is divided into three aligned tiers: *(Top)* Average Feature Contributions: A stacked bar chart displaying the average absolute SHAP values for 16 distinct AlphaGenome feature modalities, illustrating which specific functional tracks (e.g., chromatin accessibility, transcription factor binding, or evolutionary conservation) drive the model’s predictions at each base position. *(Middle)* Positional AVI Scores: The distribution of AlphaGenome Variant Impact (AVI) Phred scores across the motif window. Points indicate the median Phred score at each nucleotide position, with error bars representing the interquartile range (25th to 75th percentiles). *(Bottom)* Sequence Logo: The contribution weight matrix (CWM) representing the sequence preferences of the motif, using the full untrimmed 30bp width.

**fig. S13.**
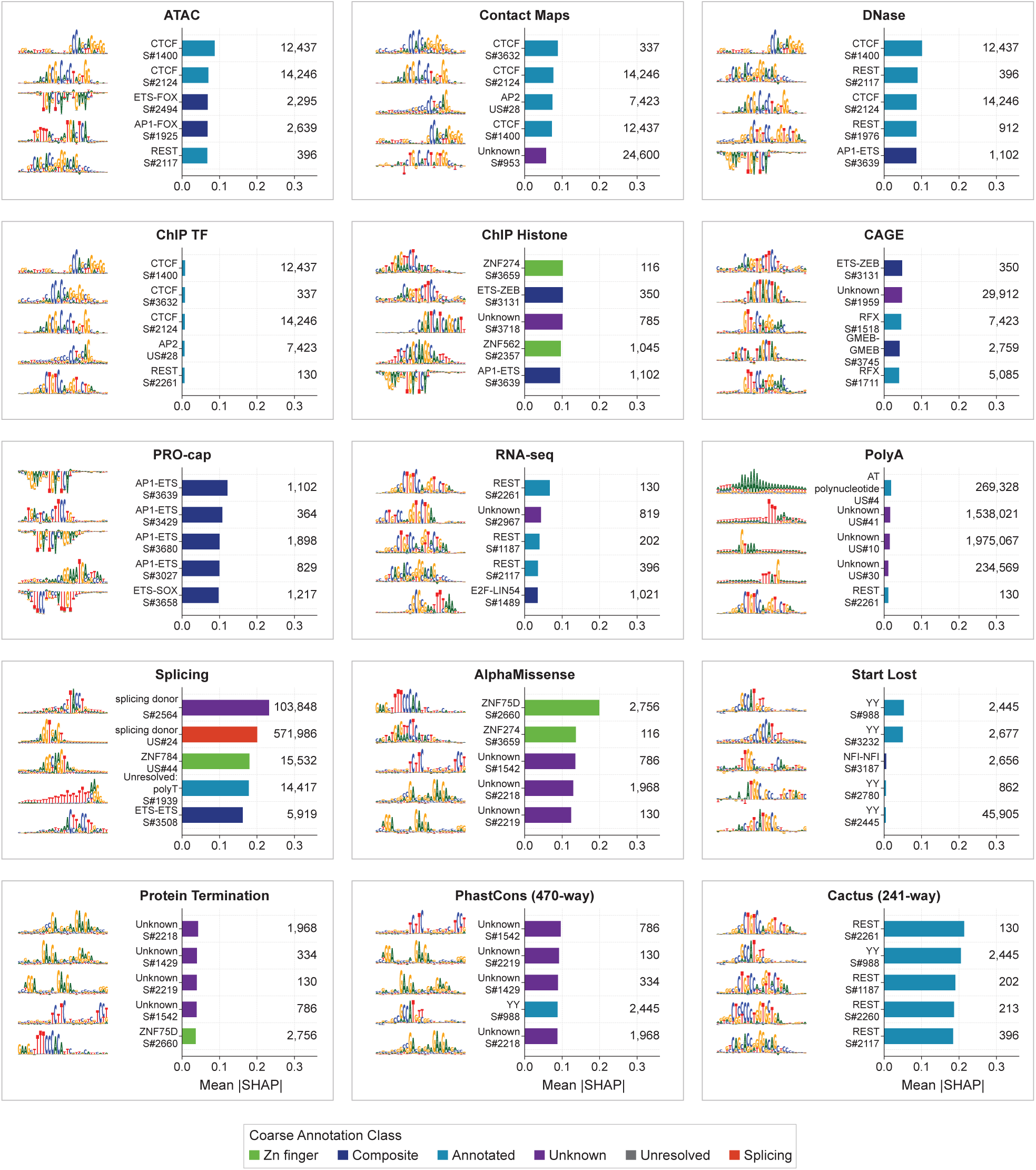
Motifs with the highest average AVI feature attributions. Horizontal bar plots display the top 5 motif clusters with the highest mean attribution for 15 of the 16 individual features comprising the AlphaGenome Variant Impact (AVI) score (the ‘Stop lost’ feature is excluded from this visualization due to very low overall attribution signals). Feature attributions were computed by retrieving high-confidence genomic instances of each motif, extracting their absolute SHAP values per feature, and averaging across all base positions within the mapped core motif instances (evaluating per-nucleotide mean attribution across instance positions to prevent bias by motif length) and across all instances. Bars are color-coded by the motif’s coarse functional annotation class (bottom legend). The total number of genomic instances aggregated is indicated on the right of each panel. Sequence logo insets display the motif cluster’s full 30 bp CWM.

**fig. S14.**
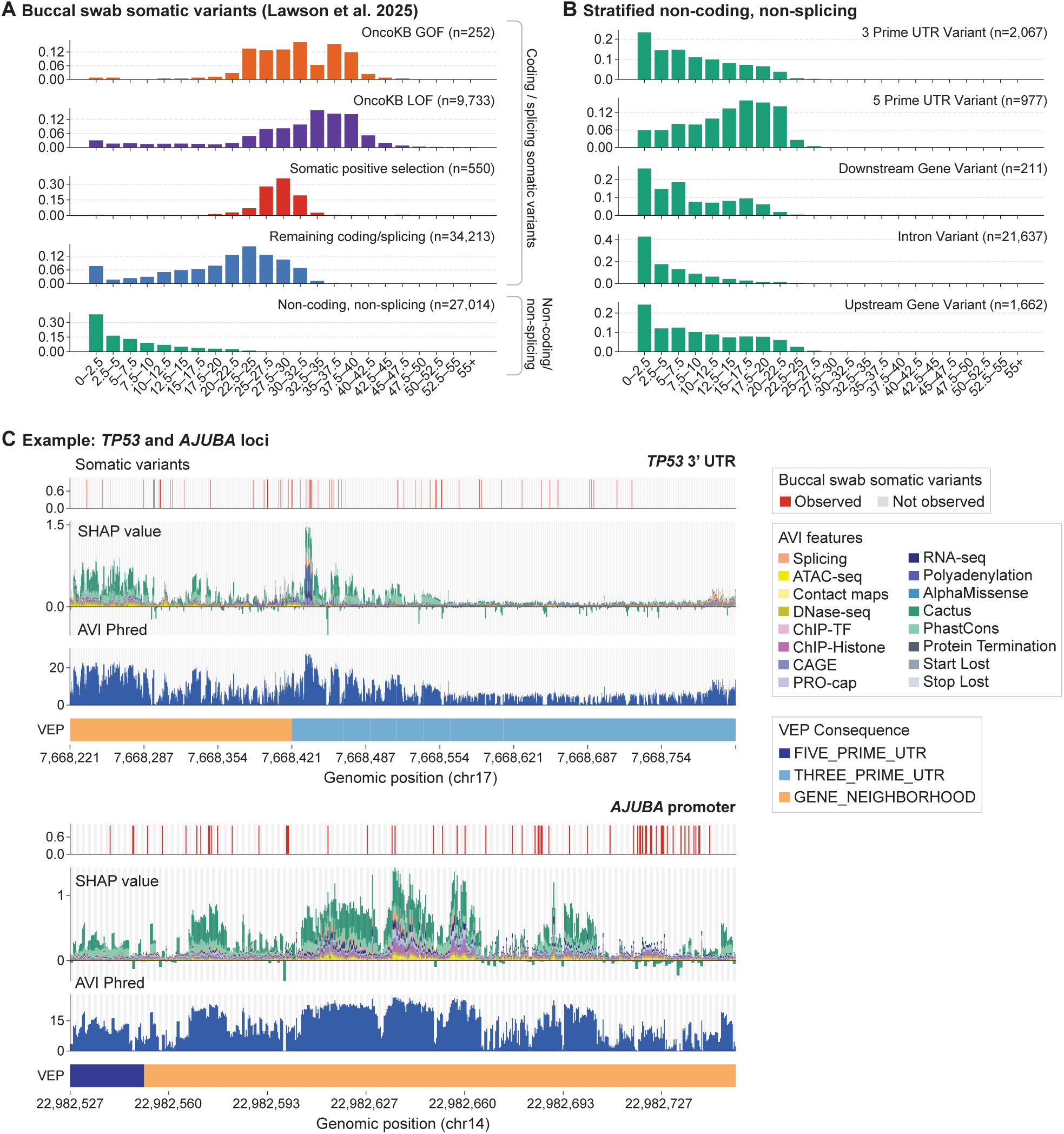
Somatic variant AVI distributions. **(A)** Bar plots grouping buccal swab somatic variants into AVI PHRED bins of width 2.5, stratified by their presence in the OncoKB database under gain of function (GOF) and loss of function (LOF) categories, followed by the somatic positive selection variant annotations. ‘Remaining coding/splicing’ indicates unannotated coding and splicing variants, while ‘non-coding, non-splicing’ indicates all remaining variants. **(B)** Further stratification of the ‘non-coding, non-splicing’ category into non-coding consequences annotated by OncoKB. **(C)** Example loci with cancer somatic variants. Each column indicates an individual variant. Observed somatic variants are indicated in red. *TP53* 3’ UTR region and *AJUBA* promoter region. Grey and white stripes mark base-pair resolution. For somatic variant annotations and AVI SHAP score, the 3 data points within each stripe represent the 3 alternative bases at allelic resolution. Alternative variants are ordered alphabetically.

**fig. S15.**
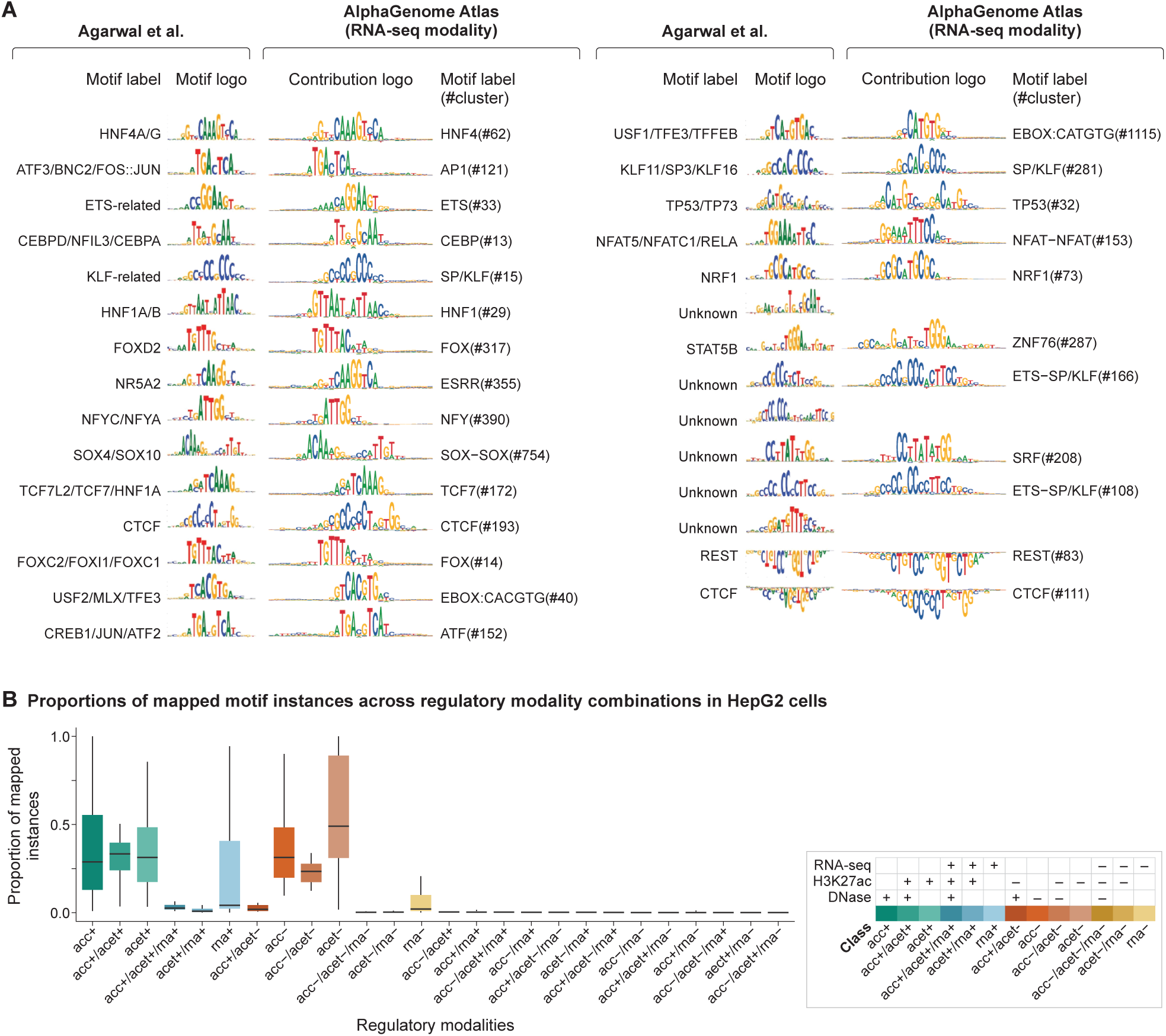
Validation of mapped motifs in HepG2 cells via AlphaGenome Atlas. **(A)** In HepG2 cells, a side-by-side comparison of the motifs returned from MPRALegNet (*60*) and motif clusters returned from the RNA-seq modality of the AlphaGenome Atlas. AlphaGenome Atlas motif clusters are represented as ISM contributions. We note that in two cases, previously unknown motifs are now annotated as composite ETS-SP/KLF motif clusters. **(B)** Proportions of mapped motif instances which are mapped by different combinations of regulatory modalities, where each observation is a different motif instance mapped in HepG2 cells for either the DNase (acc), H3K27ac ChIP-seq (acet), or total RNA-seq (rna) modality for either activating (+) or repressing (-) effects.

**Table S1.** Summary of the 19 recommended variant scoring configurations included in the AlphaGenome Atlas. Please refer to the original AlphaGenome paper. (***9***) **for details on scorer implementations. For stranded scorers, the strands have been merged. Gene annotations were queried from GENCODE GTF V46.**

| Modality Name | Scorer | Scorer Parameters | Unique Biosam-<br>ples | Signed | Total<br>Tracks | AVI | Motifs |
| --- | --- | --- | --- | --- | --- | --- | --- |
| **ATAC** | CenterMaskScorer | width=501, aggregation_type=DIFF_LOG2_SUM | 167 | ✓ | 167 | ✓ | ✓ |
| **ATAC (Active)** | CenterMaskScorer | width=501, aggregation_type=ACTIVE_SUM | 167 |  | 167 |  |  |
| **DNase** | CenterMaskScorer | width=501, aggregation_type=DIFF_LOG2_SUM | 305 | ✓ | 305 | ✓ | ✓ |
| **DNase (Active)** | CenterMaskScorer | width=501, aggregation_type=ACTIVE_SUM | 305 |  | 305 |  |  |
| **ChIP-TF** | CenterMaskScorer | width=501, aggregation_type=DIFF_LOG2_SUM | 163 | ✓ | 1,617 | ✓ | ✓ |
| **ChIP-TF (Active)** | CenterMaskScorer | width=501, aggregation_type=ACTIVE_SUM | 163 |  | 1,617 |  |  |
| **ChIP-Histone** | CenterMaskScorer | width=2001, aggregation_type=DIFF_LOG2_SUM | 219 | ✓ | 1,116 | ✓ | ✓ |
| **ChIP-Histone (Active)** | CenterMaskScorer | width=2001, aggregation_type=ACTIVE_SUM | 219 |  | 1,116 |  |  |
| **CAGE** | CenterMaskScorer | width=501, aggregation_type=DIFF_LOG2_SUM | 264 | ✓ | 546 | ✓ | ✓ |
| **CAGE (Active)** | CenterMaskScorer | width=501, aggregation_type=ACTIVE_SUM | 264 |  | 546 |  |  |
| **PROCAP** | CenterMaskScorer | width=501, aggregation_type=DIFF_LOG2_SUM | 6 | ✓ | 12 | ✓ | ✓ |
| **PROCAP (Active)** | CenterMaskScorer | width=501, aggregation_type=ACTIVE_SUM | 6 |  | 12 |  |  |
| **RNA-seq** | GeneMaskLFCScorer | - | 285 | ✓ | 371 | ✓ | ✓ |
| **RNA-seq (Active)** | GeneMaskActiveScorer | - | 285 |  | 371 |  |  |
| **Polyadenylation** | PolyadenylationScorer | - | 285 |  | 371 | ✓ | ✓ |
| **Splice Sites** | GeneMaskSplicingScorer | requested_output=SPLICE_SITES, width=None | - |  | 2 | ✓ | ✓ |
| **Splice Site Usage** | GeneMaskSplicingScorer | requested_output=SPLICE_SITE_USAGE, width=None | 282 |  | 367 | ✓ | ✓ |
| **Splice Junctions** | SpliceJunctionScorer | - | 282 |  | 367 | ✓ | ✓ |
| **Contact Maps** | ContactMapScorer | - | 12 |  | 28 | ✓ | ✓ |

**Table S2.** Performance across benchmark evaluations with ablations of various feature combinations of the AVI model. Absolute AUPRC values are shown for the AVI baseline model, while subsequent rows display the percentage difference relative to this baseline. The highest performing result for each evaluation is bolded. Rows designated as Features Set to Zero were evaluated by masking the corresponding features at inference without retraining. The ‘+’ symbol indicates that the ‘No Indel Type’ condition was applied to the model from the row immediately above. These rows demonstrate the effect of imputing AlphaMissense features and the reliance on Indel Type. Retrained rows represent ensemble models retrained under identical hyperparameters to the AVI model, with the indicated features masked to zero during training. Indentation indicates subfeatures within the broader feature category above them. Feature group ablations were defined as follows: the Protein group removed AlphaMissense and VEP consequence features, and Conservation removed both PhastCons and Cactus features.

|  | Ablation | ClinVar | ncVar | SGE | TraitGym |  |
| --- | --- | --- | --- | --- | --- | --- |
|  |  |  |  |  | Complex | Mendelian |
| Baseline | AVI Model | **0.978** | 0.936 | 0.731 | 0.283 | 0.764 |
| Features Set to Zero | + No Indel Type | 0.00% | 0.06% | -0.55% | 0.00% | 0.00% |
|  | No AlphaMissense Indel Features | -0.29% | 0.00% | -4.14% | 0.00% | 0.00% |
|  | + No Indel Type | -0.40% | 0.06% | -5.14% | 0.00% | 0.00% |
| Retrained | AlphaGenome | -2.56% | -17.61% | -8.29% | -17.27% | -2.83% |
|  | Splicing Removed Only | -1.98% | -14.97% | -7.46% | -2.61% | **0.77%** |
|  | Protein | -17.27% | 0.10% | -23.76% | **1.13%** | 0.77% |
|  | AlphaMissense | -2.51% | **0.18%** | -0.45% | 0.67% | 0.75% |
|  | Consequence | -13.71% | 0.10% | -27.42% | 0.28% | 0.26% |
|  | Conservation | -0.58% | -4.26% | -2.36% | -26.25% | -37.96% |
|  | Cactus | -0.13% | -0.91% | -1.32% | -6.64% | -16.47% |
|  | PhastCons | -0.09% | 0.04% | -5.20% | -2.15% | -0.93% |
|  | Indel Type | -0.05% | -0.44% | **0.08%** | -2.19% | 0.69% |
|  | All except AlphaGenome | -54.88% | -3.82% | -47.15% | -27.73% | -38.95% |

**Table S3.**
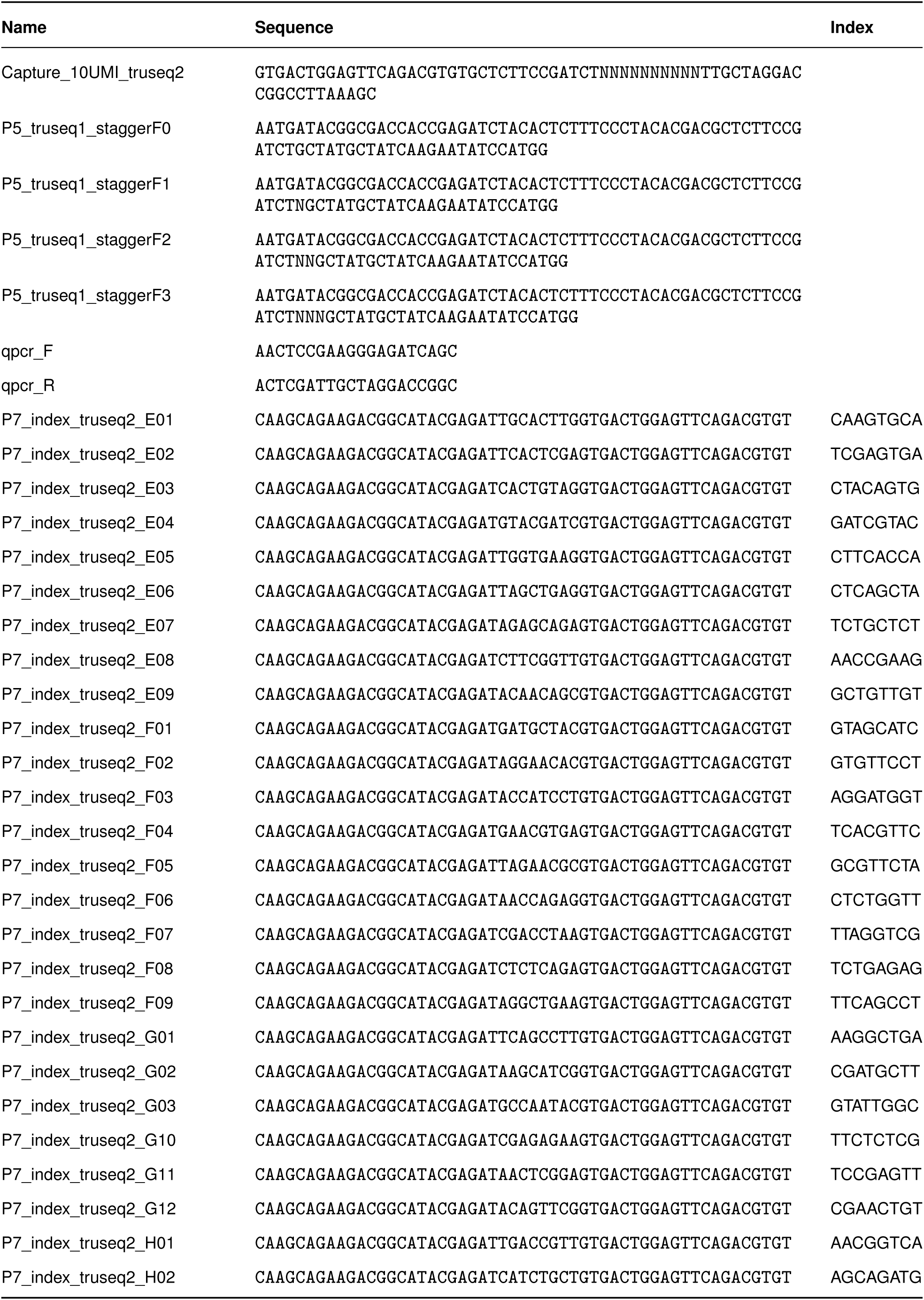
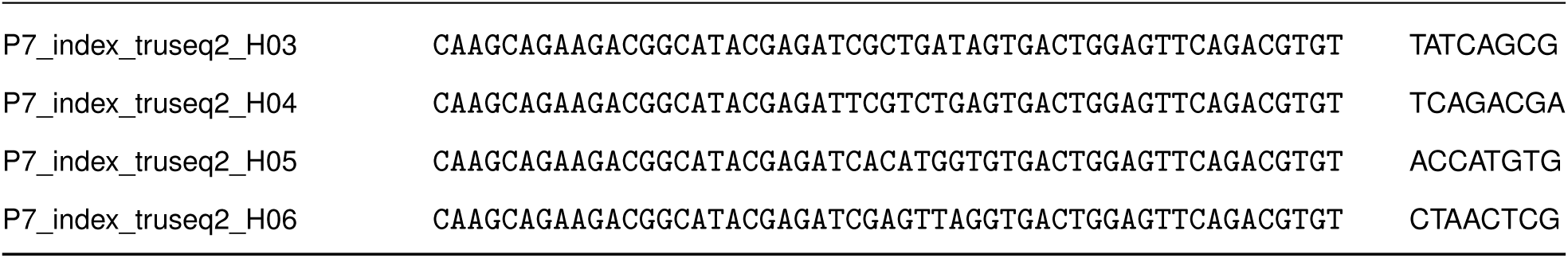
Oligonucleotide sequences used in the high throughput screening assay of *DNM1*.

**Table S4.**
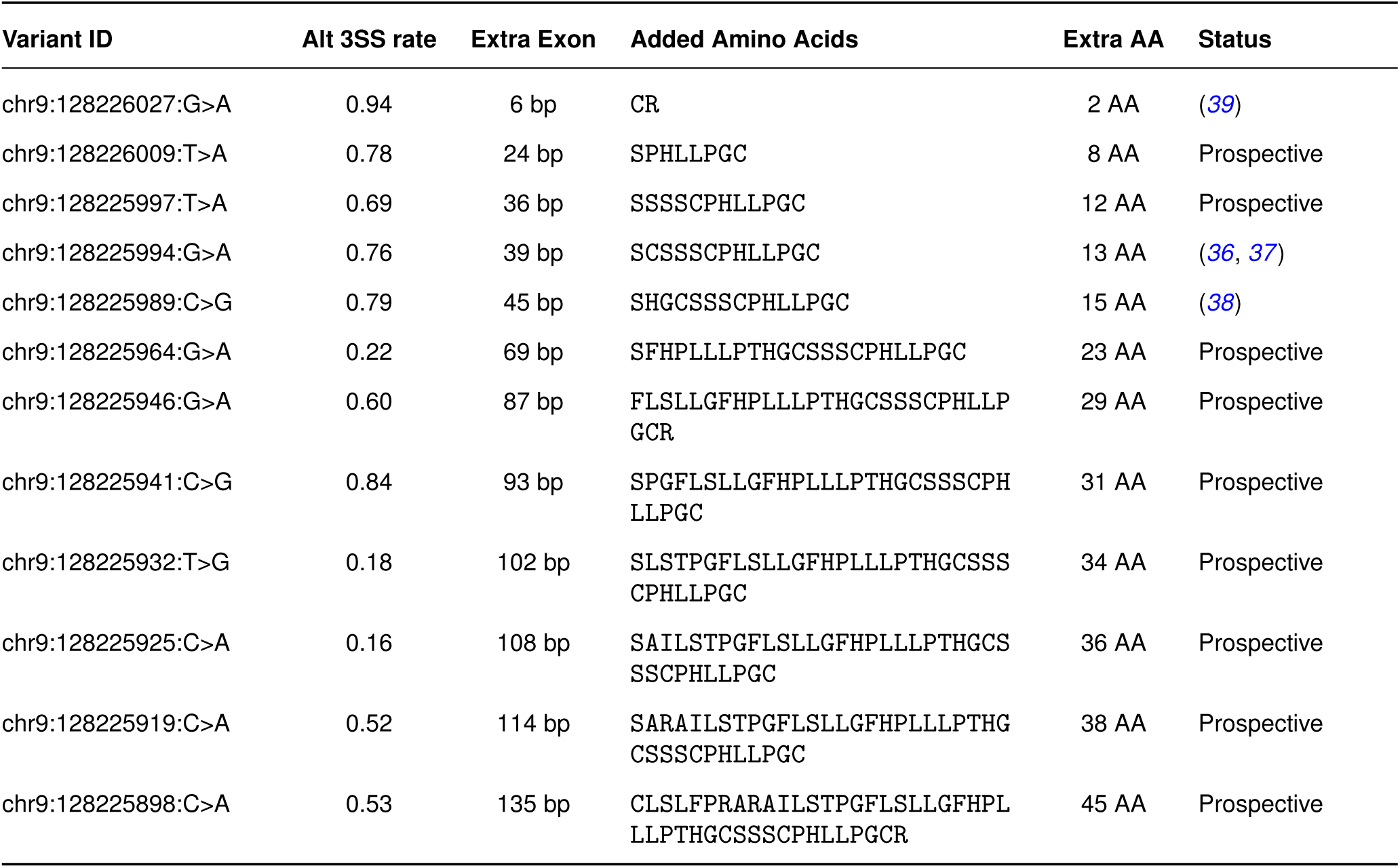
Table of prospective in-frame exon extension variants affecting *DNM1* exon 10a. Notably, the 3’ splice site selection rate is important to consider in evaluating the possibility of dominant negative effect.

**Table S5.** Annotations used in burden testing. Genetic variants included in each grouping. UTR = Untranslated Region, 3’ = variants at the 3’ end of a transcript, 5’ = variants at the 5’ end of a transcript, GERP = Genomic Evolutionary Rate Profiling score (a measure of conservation), Start Gained/Lost = the inclusion or removal of a start codon, Downstream = downstream of a transcript, CADD = Combined Annotation Dependent Deletion score.

| Classification | Mask | Consequences |
| --- | --- | --- |
| Proximal | 3'UTR | 3' UTR |
|  | 3' UTR (GERP>2) | 3' UTR (GERP>2) |
|  | 5' UTR | 5' Start Gained, 5' Start Lost, 5' Start Retained |
|  | 5' Start Gained | 5' Start Gained |
|  | 5' Start Lost | 5' Start Lost |
|  | Conserved and Intronic | Constrained |
|  | Downstream Any | Downstream |
|  | Downstream and conserved | Downstream with GERP>2 |
|  | Downstream and deleterious | Downstream with CADD>25 |
|  | Downstream and constrained | Downstream with JARVIS > 0.99 |
|  | Intron Splice Variant with SpliceAI>0.70 | Intron Splice Acceptor gain/loss with SpliceAI>0.70, Intron Splice Donor gain/loss with SpliceAI>0.70 |
|  | Splice Variant | Splice Region Variant |
|  | Upstream and conserved | Upstream Variant (GERP>2) |
|  | Upstream and deleterious | Upstream Variant (CADD > 25) |
|  | Upstream and constrained | Upstream Variant (JARVIS > 0.99) |
|  | Upstream Variant | Upstream Variant |
|  | RNA | Non-coding exon variant |
| Regulatory | Conserved, Constrained and Intergenic | Constrained and Conserved |
|  | Conserved (GERP >2) Constrained and Intergenic | Constrained and conserved with GERP >2 |
|  | Regulatory Region Variant | Regulatory Region Variant |
|  | Conserved (PhastCons 30) | Top 1% conserved variants in PhastCons 30 window |
|  | Conserved (PhastCons 100) | Top 1% conserved variants in PhastCons 100 window |
|  | PhastCons100&30 and Conserved | Any PhastCons variant (top 1%) for both PhastCons 100 and 30 and conserved (GERP>2) |
|  | PhastCons100 and Conserved | PhastCons100 (top 1%) and conserved (GERP>2) |
|  | PhastCons30 and Conserved | PhastCons30 (top 1%) and conserved (GERP>2) |
|  | PhastCons100 and Conserved at any level | Any PhastCons variant (top 1%) for PhastCons 100 and conserved |
|  | PhastCons30 and Conserved at any level | Any PhastCons variant (top 1%) for PhastCons 30 and conserved |
|  | RNA | Non-coding exon variant |
| Coding | Synonymous | Synonymous |
|  | Missense | Missense |
|  | Missense with CADD>25 | Missense variant (CADD>25) |
|  | LoF | High Confidence Loss of Function |
|  | Splice Region | Splice Region Variant |
|  | Highly Damaging Splice Region | Splice region variant (SpliceAI > 0.7) |

**Table S6.** Region-based annotations tested for each AlphaGenome score.

| Alpha Genome Score | Region Annotations |
| --- | --- |
| contact_maps | upstream, rna_regulatory, constrained, constrained_conserved, conserved_phastcons30way, regulatory_region_variant, sliding_window |
| atac | upstream, rna_regulatory, constrained, constrained_conserved, conserved_phastcons30way, regulatory_region_variant, sliding_window |
| dnase | upstream, 5_prime_UTR_variant, rna_regulatory, constrained, constrained_conserved, conserved_phastcons30way, regulatory_region_variant, sliding_window |
| chip_histone | upstream, 5_prime_UTR_variant, rna, rna_regulatory, constrained, constrained_conserved, conserved_phastcons30way, regulatory_region_variant, sliding_window |
| chip_tf | upstream, 5_prime_UTR_variant, rna_regulatory, intronic_splice, constrained, constrained_conserved, conserved_phastcons30way, regulatory_region_variant, sliding_window |
| cage | upstream, 5_prime_UTR_variant, rna, rna_regulatory, intronic_splice, 3_prime_UTR_variant, constrained, constrained_conserved, conserved_phastcons30way, regulatory_region_variant, sliding_window |
| procap | upstream, rna_regulatory, constrained, constrained_conserved, conserved_phastcons30way, regulatory_region_variant, sliding_window |
| rna_seq | upstream, 5_prime_UTR_variant, rna, rna_regulatory, intronic_splice, downstream, constrained, constrained_conserved, conserved_phastcons30way, regulatory_region_variant, sliding_window |
| splice_junctions | intronic_splice |
| splice_sites | intronic_splice |
| splice_site_usage | intronic_splice |
| polyadenylation | 3_prime_UTR_variant |

**Data S1 to S3**

**Data S1: Experimental splicing mutagenesis screening data of *DNM1* across cell lines.**

**Data S2: Statistical testing results for rare non-coding aggregates for circulating protein levels and complex traits.**

**Data S3. Interactive overview of average contribution effects of all motifs across all cell types for every regulatory modality.**

## Notes

### Competing Interest Statement

J.C., K.R.T., L.N., J.P., C.B., M.P., T.W., R.W.T., N.L., D.H., T.K., L.T., Y.U., C.A.S., A.F., M.N., V.J., R.G., L.H.W., V.D., A.M., A.G., E.A., G.N., P.K., and Z.A. performed their work in the course of employment at Google DeepMind, with no other competing financial interests. B. I. performed their work in the course of employment at Google Research, with no other competing financial interests. E.A., K.R.T, G.N., L.E.N., T.W., Z.A, and J.C. have filed patent applications relating to AlphaGenome (PCT/US2026/017924, PCT/US2026/017862). C.A.L. is a consultant to Cartography Biosciences, unrelated to this work. J.Z. owns a patent on ChIP-nexus (no. 10287628). H.L.R. has received funding for rare disease research from Microsoft, unrelated to this work. A.O'D-L. has received research support from Pacific Biosciences, unrelated to this work. F.C. is an academic founder of Curio Biosciences, Alive Bio, and scientific advisor for Amber Bio. F.C.'s interests were reviewed and managed by the Broad Institute in accordance with their conflict-of-interest policies.

### Author Declarations

The IRB of Massachusetts General Hospital gave ethical approval for work with the GREGoR consortium dataset and DNM1 family RGP_2167 of the Rare Genomes Project at the Broad Institute of MIT and Harvard under protocols #2013P001477 and #2016P001422.

