## Supplementary material for "AlphaGenome Atlas: *in silico* mutagenesis of the entire human genome improves prioritization and interpretation of non-coding variants": Data S3: DataS3_motif_contribution_heatmap.html

AlphaGenome Motif Compendium


Loading AlphaGenome Motif Compendium...

Initializing multi-modal assays, CWM sequence logos & matrix data...

### AlphaGenome Motif Compendium Interactive

Copy Config


SVG


PNG

View:

i

**Fixed:** Standard crisp cell size with vertical and horizontal scrolling.  
  
**Fit Width:** Dynamically scales column widths to fit viewport width (scrollable vertically).  
  
**Fit Screen:** Scales both cell width and height to fit full compendium in viewport without scrolling.

Fixed
Fit Width
Fit Screen

Zoom:

i

Use **Cmd + Scroll** (Mac) or **Ctrl + Scroll** (Linux/Win) over the heatmap to zoom in/out interactively.

−
100%
+
Reset

Inspected Motif

Hover over heatmap

-

Sequence Logo
-

Track & Cell Details

Biosample: -

Lineage: -

Assay Track: -

Quantitation

i

**Scaled [0-1]:** Normalized intensity value clipped at the 99th percentile across TF motifs, used to compute the bivariate Teal/Orange color blend.  
  
**Raw ISM:** In-silico mutagenesis delta prediction (ΔlogFC or Δsignal) averaged across genomic motif instances.  
  
**Fraction (%):** Percentage of unique genomic motif instances having |z| ≥ 2.0 or Phred ≥ 30.

(+) Scaled [0-1]: -

(-) Scaled [0-1]: -

(+) Raw ISM: -

(-) Raw ISM: -

Directionality: -

Plot Display Controls

▶

Motif granularity:

i

**Coarse:** Aggregated TF motif labels (325 rows) with 3-tier lineage staircase clustering.  
  
**Granular:** Individual motif clusters (1,680 rows).

Coarse
Granular

Cluster by:

i

**Coarse:** Expands coarse rows at the same position, hierarchically clustering (+ above -) inside each coarse block.  
  
**Granular:** Global 3-tier lineage staircase clustering across individual motif clusters.

Coarse
Granular

Instance metric:

i

**Importance:** Continuous in-silico mutagenesis effect magnitude.  
  
**Amount:** log10(1 + count) of active motif instances passing significance filter.  
  
**Fraction:** Fraction of genome-wide motif occurrences actively functional.

Importance
Amount
Fraction

Scale:

i

**Log1p:** log(1 + x) transformation compressing high dynamic range outliers.  
**Linear:** Directly proportional linear response.

Log1p
Linear

▶ Assay scale multipliers
Reset


Modality & Track Selection

i

Select active modalities and dynamically filter to common biosample tracks.

▶

Visible Modalities:

↺ Reset
Toggle All

✓ All tracks in common (35)

Showing **35** biosamples
Show all (698)

Lineage aggregation:

i

**None:** Displays all individual biosample tracks.  
**Take first:** Displays 1 representative biosample per lineage.  
**Average:** Averages signal across all biosamples belonging to each lineage.

None
Take first
Average

Custom Selection:
📋 Copy visible (35)

Filter with exact names or wildcards (e.g. CD4\*).

Ordering:

Original
Specified

Motif Selection

i

Filter rows by category, specify custom motif lists, or prune low-signal rows.

▶

All
Main Motifs
Zinc Fingers
Composite

Unknown

Unknown motif clusters are only available when selecting Granular granularity and cluster by: granular.

Custom

Custom motif list:
📋 Copy visible

Edit, filter, or paste custom motif names to include in heatmap.

Ordering:

Original
Specified

Signal Pruning:

Mean Across Visible
Max Across Visible
RNA-seq (Expression)
DNase (Accessibility)
H3K27ac (Active Enh.)
H3K9me3 (Silencing)
H3K4me3 (Promoters)
H3K4me1 (Primed Enh.)
H3K27me3 (Polycomb)
H3K36me3 (Gene Body)

Keep Top:

100%

Showing all motifs

Remove all 0:

i

Drop motifs with zero signal across all currently visible modalities and displayed biosamples.

False
True

Context Actions

📋 Copy Motif Name

📋 Copy Motif ID

📋 Copy Track Name

📋 Copy Biosample

📋 Copy Assay Name
